# Prenatal urban environment and blood pressure trajectories from childhood to early adulthood

**DOI:** 10.1101/2023.03.31.23288002

**Authors:** Ana Gonçalves Soares, Susana Santos, Emie Seyve, Rozenn Nedelec, Soile Puhakka, Aino-Maija Eloranta, Santtu Mikkonen, Wen Lun Yuan, Deborah A Lawlor, Jon Heron, Martine Vrijheid, Johanna Lepeule, Mark Nieuwenhuijsen, Serena Fossati, Vincent W V Jaddoe, Timo Lakka, Sylvain Sebert, Barbara Heude, Janine F Felix, Ahmed Elhakeem, Nicholas J Timpson

## Abstract

**Background:** Prenatal urban environmental exposures have been associated with blood pressure in children. The dynamic of these associations across childhood and later ages is unknown.

**Objectives:** To assess associations of prenatal urban environmental exposures with blood pressure trajectories from childhood to early adulthood.

**Methods:** Repeated measures of systolic (SBP) and diastolic blood pressure (DBP) were collected in up to 7,454 participants from a UK birth cohort. Prenatal urban exposures (n=42) covered measures of noise, air pollution, built environment, natural spaces, traffic, meteorology, and food environment. An exposome-wide association study approach was used. Linear spline mixed-effects models were used to model associations of each exposure with trajectories of blood pressure. Replication was sought in four independent European cohorts (N up to 9,261).

**Results:** In discovery analyses, higher humidity was associated with a faster increase (mean yearly change in SBP for an interquartile range [IQR] increase in humidity: 0.29 mmHg/year, 95%CI 0.20; 0.39) and higher temperature with a slower increase (mean yearly change in SBP per IQR increase in temperature: -0.17 mmHg/year, 95%CI -0.28; -0.07) in SBP in childhood. Higher levels of humidity and air pollution were associated with faster increase in DBP in childhood and slower increase in adolescence. There was little evidence of an association of other exposures with change in SBP or DBP. Results for humidity and temperature, but not for air pollution, were replicated in other cohorts.

**Conclusion:** Replicated findings suggest that higher prenatal humidity and temperature could modulate blood pressure changes across childhood.

## Introduction

Blood pressure is an important modifiable risk factor for cardiovascular disease (CVD)^1,2^ and it tracks from childhood into adulthood.^3^ Elevated blood pressure in childhood or adolescence is associated with several intermediate markers of CVD and with CVD morbidity and mortality in adulthood.^4^ There is a growing body of evidence showing that urban environmental exposures, such as air pollution,^5^ noise,^6^ temperature,^7^ and some characteristics of the built environment^8^ are associated with elevated blood pressure/hypertension in adulthood,^9,10^ and some associations have also been observed in children.^11-14^

Early life, especially the prenatal and early postnatal periods, is a phase of rapid development and particularly vulnerable to environmental factors, during which adverse exposures may lead to higher risk of CVD.^15,16^ Some studies have shown associations of prenatal urban environmental exposures with blood pressure levels in children.^12-14^ Previous studies have mostly measured blood pressure at a single time point, and none have sought replication of the findings.

Studies exploring early-life urban environmental exposures and later blood pressure have predominantly focused on single exposures, or a specific, related group of exposures, particularly air pollution,^13,17,18^ and very few have used a more holistic view of the urban environment.^12,14^ To the best of our knowledge, no study has systematically explored a range of urban environmental exposures and their longitudinal association with blood pressure using repeated measures. Understanding whether associations vary over time is needed to determine the potential impact of the prenatal environment on future CVD.

Our aim was to assess the association of a range of prenatal urban environmental exposures with changes in systolic (SBP) and diastolic blood pressure (DBP) from childhood to early adulthood.

## Methods

Data from the Avon Longitudinal Study of Parents and Children (ALSPAC) were used for the discovery analysis. ALSPAC is a prospective population-based cohort (see www.alspac.bris.ac.uk) that recruited pregnant women living in the former Avon area of the UK who were due to give birth between April 1991 and December 1992.^19-21^ In total, 14,541 pregnancies were enrolled, which resulted in 14,062 live births. Children, mothers and their partners have been followed up repeatedly ever since.^19-21^ The study website contains details of all the data that is available through a fully searchable data dictionary and variable search 4 tool. For full details of the data from the ALSPAC study, see http://www.bristol.ac.uk/alspac/researchers/our-data.

Ethical approval for the study was obtained from the ALSPAC Ethics and Law Committee and the Local Research Ethics Committees. Informed consent for the use of data collected via questionnaires and clinics was obtained from participants following the recommendations of the ALSPAC Ethics and Law Committee at the time. Consent for biological samples has been collected in accordance with the Human Tissue Act (2004). More information on the study ethics is available at http://www.bristol.ac.uk/alspac/researchers/research-ethics/.

### Urban environmental exposures

In total, 46 urban environmental exposures were included in this study (Supplementary Table 1). All exposures were derived as part of the LifeCycle Project^22,23^ and cover noise, air pollution, built environment, natural spaces, traffic, meteorology, and unhealthy food environment. The only exception was particulate matter <10µm (PM_10_), which in ALSPAC was not modelled as part of the LifeCycle.^24^

Briefly, exposures were assigned within Geographic Information System (GIS) tools to the geocoded addresses of the participants at birth. Exposures correspond to average exposure levels during pregnancy. For PM_10_, whole pregnancy and pregnancy trimester-specific measures were available and used in the analysis. Continuous exposures were rescaled for analysis to represent changes by interquartile range (IQR) increase. Further details on the exposures can be found in supplementary material

Pairwise Pearson correlation coefficients across all exposures were estimated; for any r>0.9, only one of the pair was included in subsequent analyses, based on variable properties (e.g., continuous preferred to categorical variables, mean preferred to minimum and maximum).

### Blood pressure

Blood pressure was measured in all ALSPAC participants at clinic assessments at average ages 7, 9, 10, 11, 13, 15, 18 and 24 years, and at ages 3, 4 and 5 years in a 10% sub-sample.^19^ In all clinic assessments, SBP and DBP were measured twice with the individual sitting at rest with the arm supported, using a cuff size appropriate for the child’s upper arm circumference. The mean of the two measures was recorded and used in the analyses. More details on blood pressure measurement are described in supplementary material.

### Confounders

Analyses were adjusted for a set of confounders defined as known or plausible determinants of urban environment and blood pressure (i.e., maternal education, age at delivery, ethnicity, and area deprivation). Sex, although not a confounder, was also included in the adjustment to reduce residual variance. All confounders, except area deprivation, were obtained through questionnaires assessed during pregnancy. Area deprivation was based on data from the UK Ministry of Housing, Communities & Local Government and linked to the geocoded addresses at birth. More details on the confounders are presented in supplementary material.

### Statistical analysis

We included participants who had information on at least one prenatal urban environmental exposure and at least one blood pressure measurement between mean ages 3 and 24 years, and who had complete data on the confounders. This resulted in the inclusion of 5,095 (for noise exposures) to 7,454 (for average greenness exposures) participants with between 28,132 and 41,216 blood pressure observations (Supplementary Figure 1).

An exposome-wide association study (ExWAS) approach^25^ was used to assess the association of each exposure individually with SBP and DBP trajectories from childhood to early adulthood. Associations were examined using linear mixed-effects models with linear splines for age (as a fixed effect) to allow for nonlinear change in blood pressure with age.^26^ Knots were placed at approximate ages 10 and 18 years to summarise linear change in blood pressure across childhood (3-10 years), adolescence (10-18 years), and young adulthood (18-26 years) (more details in supplementary material). Models included random intercept and random linear slope for age to allow for between-individual differences in blood pressure at baseline and in change with age. Age was centred at 3.1 (mean age at first clinic assessment). An interaction term between each exposure and age was included in the models to assess the association of each urban environmental exposure with change in blood pressure. All models were adjusted for the confounders defined above.

To limit false positive results, a Bonferroni correction was used to account for multiple testing; an alpha level of 5% was divided by the number of exposures assessed (42 after removing one of highly correlated pairs, i.e. 0.05/42 = p-value threshold 0.0012). For associations that reached our multiple-testing p-value threshold, average predicted means for SBP and DBP from childhood to early adulthood in the 25^th^ and 75^th^ percentile of the exposure were calculated from the linear mixed-effects models.

Within each exposure-blood pressure trajectory, complete case analyses were undertaken, which gives unbiased results when the chance of being a complete is independent of the outcome after taking the covariates into consideration.^27^ To explore potential impact of missing data, we compared characteristics of those included and not included in the analysis due to missing data on one or more confounders. The analyses were performed in the software Stata 17.0 (Statacorp, College Station, TX, USA) and the software MLwiN 3.05 was accessed from within Stata using *runmlwin*.^28^

### Sensitivity analyses

To explore the sensitivity of our results to including all participants with at least one blood pressure measurement we also repeated analyses with only participants who had at least three repeated measures (N up to 6,128 with 39,358 observations).

We explored possible sex differences in the associations between prenatal urban environmental exposures and blood pressure trajectories in those exposures found to be associated with blood pressure in the main analysis after multiple-testing correction. For that, we included in the models a 3-way interaction term between exposure, age and sex, along with an interaction between age and sex, and assessed the significance of the 3-way interaction (p<0.05). We also performed sex-stratified analyses.

### Replication analysis

To assess robustness in the results found in ALSPAC, we sought replication, for associations that reached our multiple-testing p-value threshold, in four independent European cohorts participating in the LongITools Project.^29^ These were Etude des Déterminants pré et post natals précoces du développement psychomoteur et de la santé de l’ENfant (EDEN),^30^ the Generation R Study (GenR),^31^ the Physical Activity and Nutrition in Children Study (PANIC),^32^ and the Northern Finland Birth Cohort 1986 (NFBC1986).^33^ Identical methods to those used in ALSPAC for deriving exposures were used in EDEN and GenR, with the methods used in NFBC1986 and PANIC described in supplementary material. Blood pressure measurements were available at average ages (years) 2.1, 6.1 and 9.8 in GenR, 3.1 and 5.6 in EDEN, 7.6, 9.8 and 15.8 in PANIC, and 16.0 and 34.1 in NFBC1986 (Supplementary Figure 2). Information on blood pressure measurement in each study is available in supplementary material.

Linear mixed-effects models with random intercept and a random linear slope for age were used to model blood pressure trajectories in each study. Limited numbers of repeated measurements meant we had to use linear mixed-effects models without splines. In each study, age was centred at the mean age at baseline (which differed in each cohort). The set of available confounders may vary across studies hence, adjustment model may differ from the initial analysis in ALSPAC (more details in supplementary material). Results were considered to be replicated if the direction of association was consistent with the ALSPAC results and if p-value <0.05.

## Results

In ALSPAC, up to 7,454 individuals with 41,216 blood pressure observations were included in the analysis. Those included were more likely to have mothers with higher education and older age at delivery, to have White ethnicity and to live in a less deprived area at birth than those not included in the analysis due to missing data on one or more of the confounders (Supplementary Table 2). Up to 11 measures of blood pressure were used (mean 6.7, standard deviation [SD] 2.1), the mean age of the participants ranged from 3.1 (SD 0.02) at baseline to 24.4 (SD 0.79) years at the last follow-up, and the number of individuals with blood pressure measure in each assessment varied from 800 (at age 5 years) to 6,056 (at age 7.5 years) (Supplementary Table 3).

Out of the 46 environmental exposures, four were excluded due to very high correlation (Supplementary Figure 3). The distribution of the environmental exposures (in their original scale) is presented in Supplementary Table 4.

The average trajectories of SBP and DBP from childhood to early adulthood are presented in Supplementary Figure 4 and Supplementary Table 5. The mean SBP at baseline (mean age 3.1y) was 94.3 mmHg (95% confidence interval [95%CI] 89.9, 92.7), and it increased on average 1.56 mmHg/year (95%CI 1.50, 1.62) in childhood (3-10 years) and 2.57 mmHg/year (95%CI 2.53, 2.61) in adolescence (10-18 years), and decreased on average 1.02 mmHg/year (95%CI -1.08, -0.97) in early adulthood (18-26 years). The mean DBP at baseline was 54.9 mmHg (95%CI 53.9, 55.9), and it increased on average 0.17 mmHg/year (95%CI 0.12, 0.21) in childhood, 1.06 mmHg/year (95%CI 1.03, 1.09) in adolescence, and 0.14 mmHg/year (95%CI 0.09, 0.18) in early adulthood.

### Associations of environmental exposures with blood pressure trajectories

The associations of urban environmental exposures (n=42) with change in SBP and DBP from childhood to early adulthood are presented in Figure 1 and Supplementary Tables 6 and 7.

**Figure 1.**
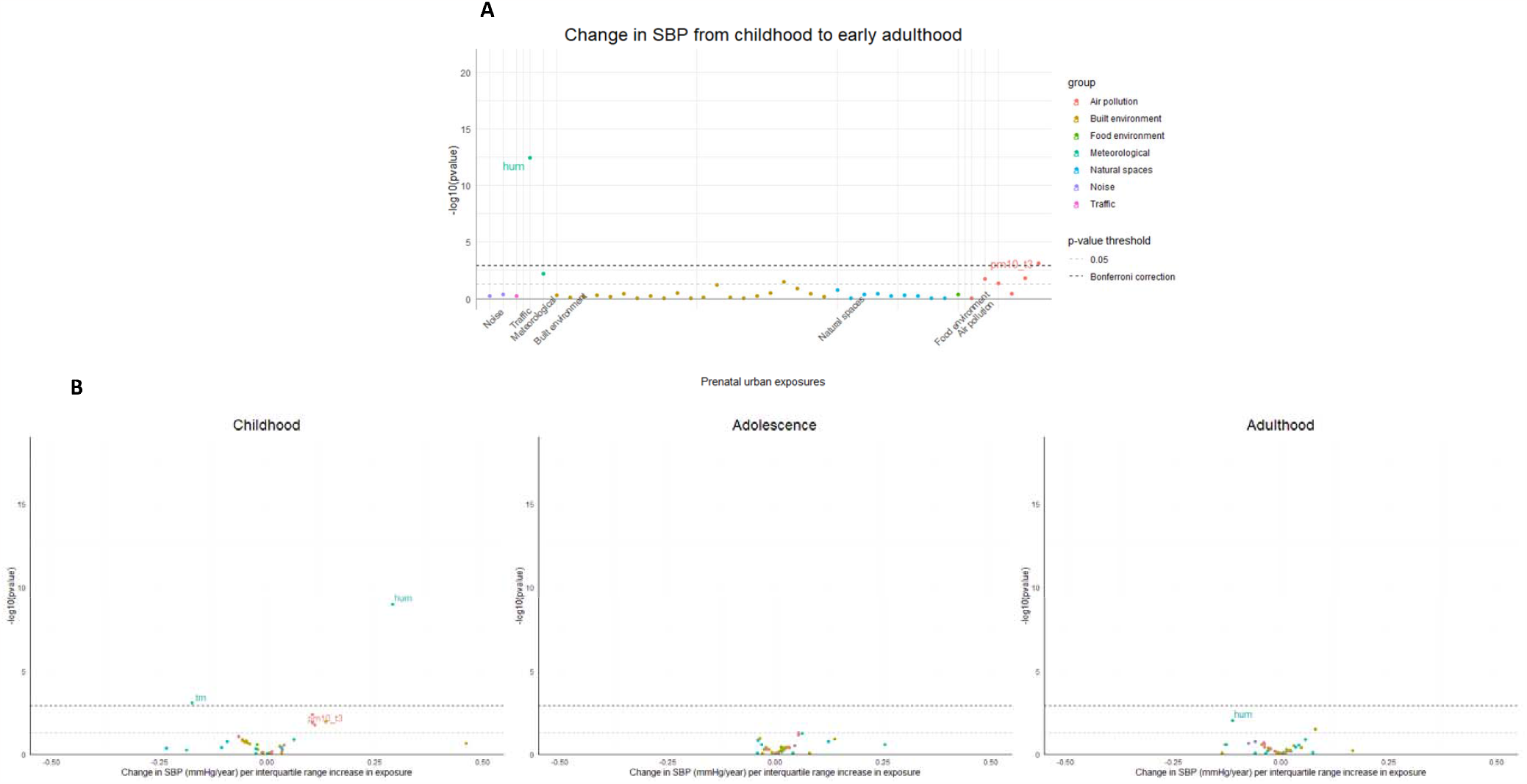
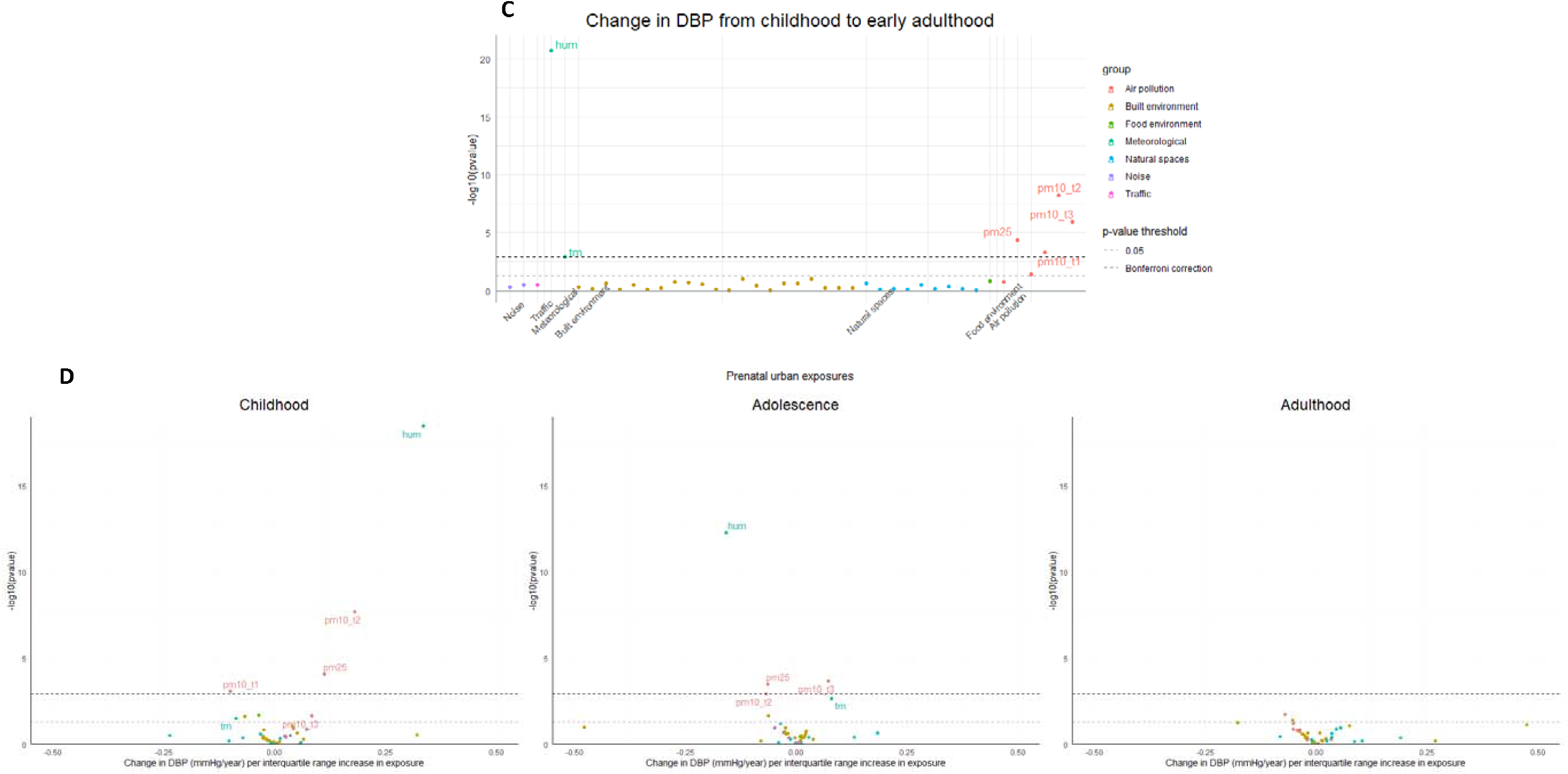
Associations of urban environmental exposures with changes in systolic (SBP) and diastolic blood pressure (DBP) in ALSPAC. Associations of prenatal urban environmental exposures with (A) SBP and (C) DBP from childhood to early adulthood, and volcano plots showing the associations of urban exposures with changes in (B) SBP and (D) DBP in childhood, adolescence and early adulthood. Beta corresponds to mean changes in blood pressure per interquartile range (IQR) increase/change in category in the exposure; the IQR for each exposure is presented in Supplementary Table 4. Models are adjusted for maternal education, age at delivery, ethnicity, area deprivation, and sex.

After Bonferroni correction, prenatal humidity and PM_10_ in the third trimester of pregnancy were associated with change in SBP from childhood to early adulthood (Figure 1A). Higher relative humidity was associated with a faster increase in SBP in childhood (mean change in SBP for an IQR increase in humidity (which corresponds to 2% increase in humidity [Supplementary Table 4]): 0.29 mmHg/year, 95%CI: 0.20, 0.39), and slower decrease in SBP in adulthood (mean change in SBP for an IQR increase in humidity: -0.11 mmHg/year, 95%CI: -0.20, -0.03) (Figure 1B, Supplementary Table 6). Higher PM_10_ in the third trimester of pregnancy was also associated with a faster increase in SBP in childhood (mean change in SBP for an IQR increase in PM_10_: 0.11 mmHg/year, 95%CI: 0.02, 0.20). Higher mean temperature was associated with a slower increase in SBP in childhood (mean change in SBP for an IQR increase in temperature: -0.18 mmHg/year, 95%CI: -0.28, -0.08). There was little evidence for an association between the other urban environmental exposures and change in SBP from childhood to early adulthood.

The average predicted trajectory of SBP from childhood to early adulthood in the 25^th^ and 75^th^ percentiles of humidity, mean temperature and PM_10_ at the third trimester of pregnancy is presented in Figure 2 and Supplementary Table 8. E.g., the average SBP in those in the 25^th^ percentile of humidity (average relative humidity 81.1%) increased from 91.9 mmHg (95%CI 91.1, 92.7%) at baseline to 101.8 mmHg (95%CI 101.1, 102.5) at age 10, whilst in the 75^th^ percentile of humidity (relative humidity 83.1%) the increase was faster, from 89.9 mmHg 95%CI 89.1, 90.7) to 101.8 mmHg (95%CI 101.1, 102.5) in the same period.

**Figure 2.**
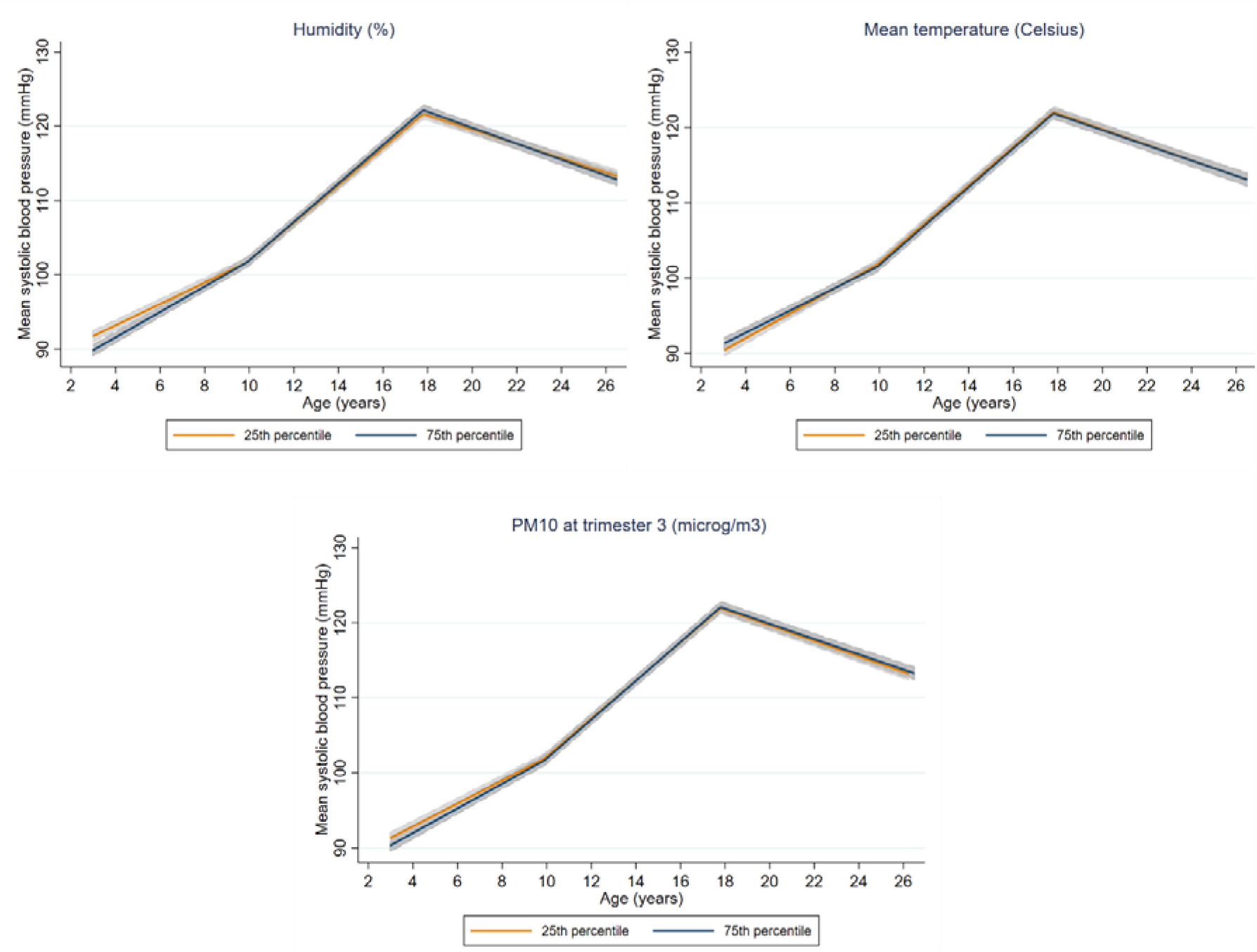
Average predicted population mean for systolic blood pressure from childhood to early adulthood in the 25^th^ and 75^th^ percentiles of relative humidity, mean temperature and PM_10_ in the third trimester of pregnancy. The orange line shows the average predicted systolic blood pressure from childhood to early adulthood in the 25^th^ percentile of exposure, and the navy line in the 75^th^ percentile of exposure. Shaded areas correspond to 95% confidence intervals. All covariates used in the adjustment were set to the mean value or the reference category: maternal education (high), age at delivery (28.9 years), ethnicity (White), area deprivation (least deprived), and sex (male).

Prenatal humidity, temperature and PM_2.5_, and PM_10_ in all trimesters of pregnancy were associated with change in DBP from childhood to early adulthood, after multiple-testing correction (Figure 1C). Higher humidity was associated with faster increase in DBP in childhood and slower increase in adolescence (mean change in DBP for an IQR increase in humidity: 0.34 mmHg/year, 95%CI: 0.26, 0.41, and -0.16 mmHg/year, 95%CI: -0.20, -0.11, respectively) (Figure 1D, Supplementary Table 7). Associations in the opposite direction were observed for temperature (mean change in DBP for an IQR increase in temperature: -

0.09 mmHg/year in childhood, 95%CI -0.17, -0.01, and 0.08 mmHg/year in adolescence, 95%CI 0.03, 0.13). Higher PM_2.5_ was associated with a faster increase in DBP in childhood and a slower increase in adolescence (mean change in DBP for an IQR increase in PM_2.5_:0.11 mmHg/year, 95%CI: 0.06, 0.17, and -0.07 mmHg/year, 95%CI: -0.10, -0.03, respectively). Higher PM_10_ in the first trimester of pregnancy was associated with a slower increase in DBP in childhood (mean change in DBP for an IQR increase in PM_10_: -0.10 mmHg/year, 95%CI: -0.16, -0.04). Higher PM_10_ in the second trimester of pregnancy was associated with a faster increase in DBP in childhood and a slower increase in adolescence (mean change in DBP for an IQR increase in PM_10_: 0.18 mmHg/year, 95%CI: 0.12, 0.24, and -0.07 mmHg/year, 95%CI: -0.11, -0.03, respectively). Higher PM_10_ in the third trimester of pregnancy was associated with a faster increase in DBP in both childhood and adolescence (mean change in DBP for an IQR increase in PM_10_: 0.08mmHg/year, 95%CI 0.01, 0.16, and 0.07 mmHg/year, 95%CI: 0.03, 0.11, respectively). Little evidence for an association between the other urban environmental exposures and change in DBP from childhood to early adulthood was observed. The average predicted trajectory of DBP from childhood to young adulthood in the 25^th^ and 75^th^ percentiles of humidity, temperature, PM_2.5_ and PM_10_ at different trimesters of pregnancy is presented in Figure 3 and Supplementary Table 8.

**Figure 3.**
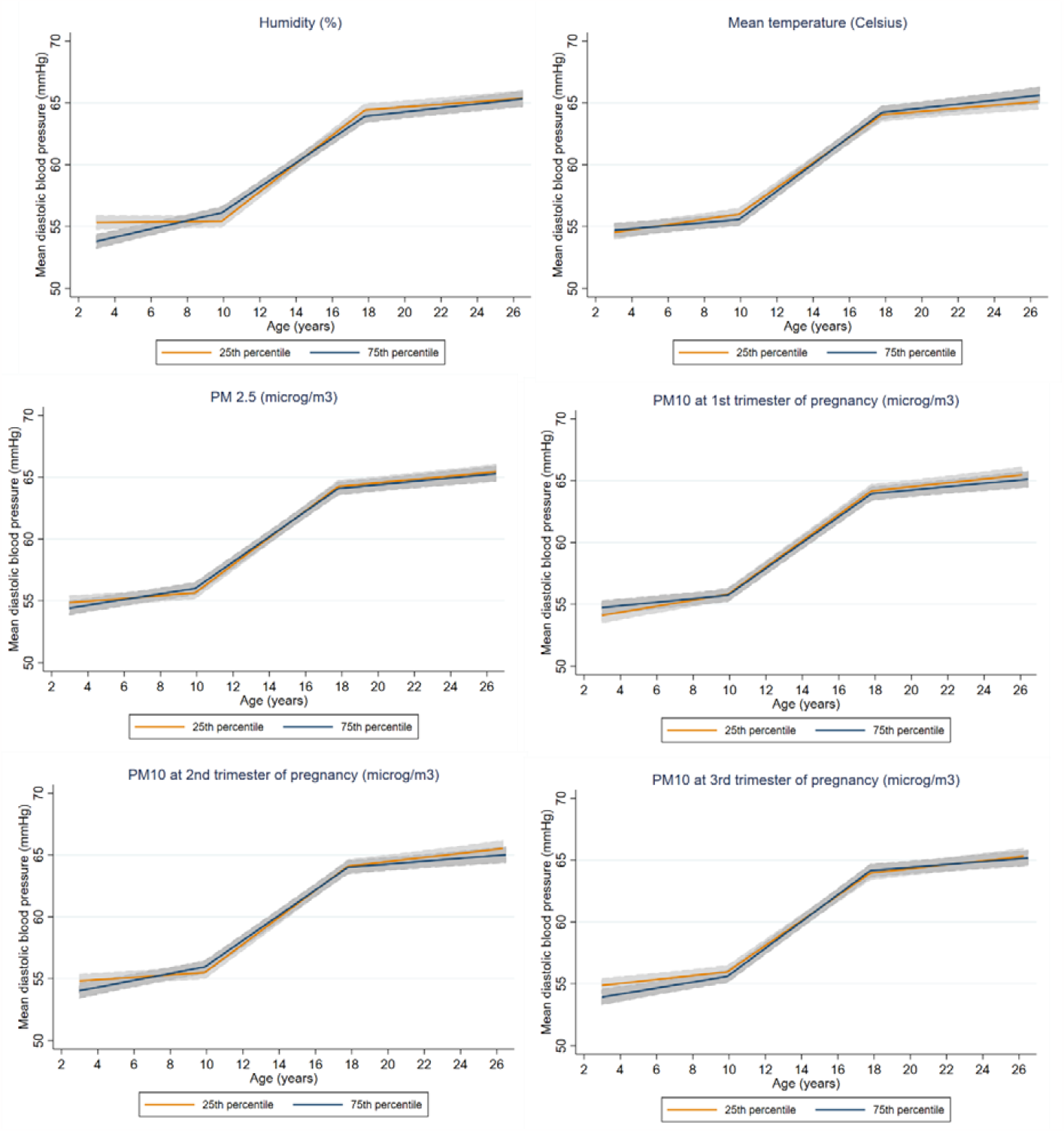
Average predicted population mean for diastolic blood pressure from childhood to early adulthood in the 25^th^ and 75^th^ percentiles of relative humidity, temperature, PM_2.5_ and PM_10_ at different trimesters of pregnancy. The orange line shows the average predicted systolic blood pressure from childhood to early adulthood in the 25^th^ percentile of exposure, and the navy line in the 75^th^ percentile of exposure. Shaded areas correspond to 95% confidence intervals. All covariates used in the adjustment were set to the mean value or the reference category: maternal education (high), age at delivery (28.9 years), ethnicity (White), area deprivation (least deprived), and sex (male).

Results for the associations of the urban environmental exposures with SBP and DBP trajectories remained similar when analyses were restricted to individuals with three or more repeated measures of blood pressure (Supplementary Tables 9 and 10).

Overall associations of humidity, mean temperature, PM_2.5_ and PM_10_ with blood pressure trajectories were similar in males and females, and there was little evidence for sex differences (Supplementary Table 11). The only two exceptions were for the association between PM_10_ in the first trimester of pregnancy and slower increase in SBP in childhood, which was only evident in females, and the association between PM_10_ in the third trimester of pregnancy and faster increase in DBP in childhood, which was stronger in males.

### Replication in other cohorts

The distribution of the baseline characteristics, environmental exposures and outcomes at each follow-up in GenR, EDEN, PANIC and NFBC1986 are presented in Supplementary Table 12. The association of the confounders with the environmental exposures in all the cohorts is presented in Supplementary Table 13. Whilst some associations between the confounders and the environmental exposures were similar across the cohorts (e.g., those living in the most deprived areas had higher levels of PM_2.5_ and PM_10_), some associations differed (e.g., low maternal education was positively associated with PM_2.5_ in ALSPAC and EDEN (Poitiers), whilst no association was evident in EDEN (Nancy) and an inverse association was observed in GenR).

The associations of humidity, mean temperature, PM_2.5_ and PM_10_ with SBP and DBP at baseline in each study are presented in Supplementary Figure 5, and associations with change in SBP and DBP are presented in Figure 4. The association of humidity with faster increase in SBP in childhood replicated in GenR and EDEN (Poitiers), and with faster increase in DBP replicated in EDEN. An association between humidity and change in SBP in adulthood was not observed in NFBC1986. The associations of temperature with slower increase in SBP and DBP in childhood were also observed in GenR and EDEN, but the association of temperature with faster increase in DBP in adolescence did not replicate in PANIC. The association between PM_2.5_ and faster increase in DBP in childhood was also observed in EDEN (Nancy), though CI spanned the null, and in GenR associations between PM_2.5_ and slower increase in both SBP and DBP in childhood were observed. No data were available for PM_2.5_ in adolescence. PM_10_ was associated with faster increase in SBP in childhood in EDEN (Nancy) but with slower increase in SBP and DBP in childhood in GenR. In GenR, the association of PM_10_ in the first trimester of pregnancy with slower increase in DBP in childhood was also observed, but associations of PM_10_ in the second and third trimesters of pregnancy with blood pressure were observed in the opposite direction to those observed in ALSPAC (Supplementary Figure 6).

**Figure 4.**
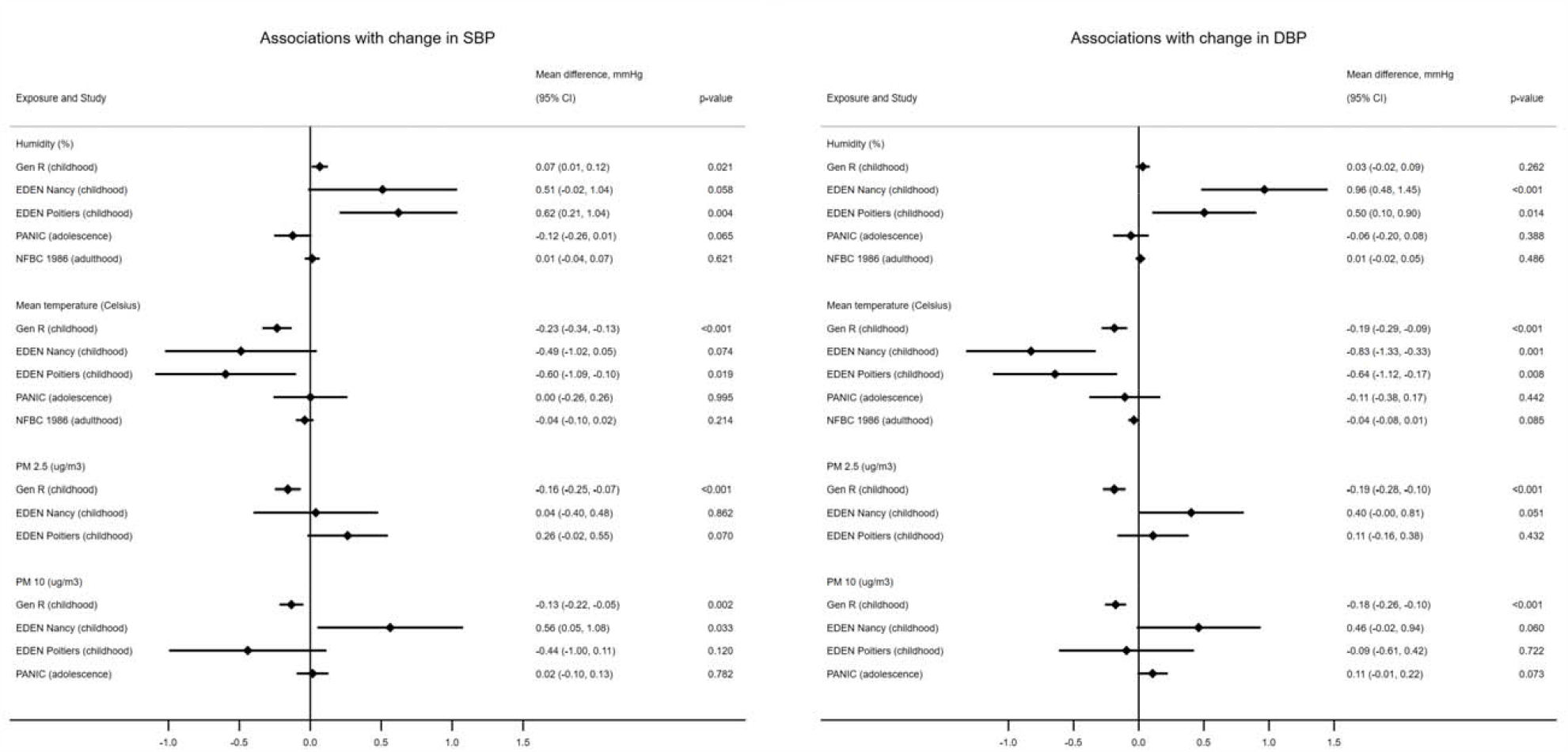
Associations of humidity, mean temperature, PM_2.5_ and PM_10_ with change in SBP and DBP in GenR, EDEN, PANIC and NFBC1986.

## Discussion

We assessed the association of 42 prenatal urban environmental exposures with blood pressure trajectories from childhood to early adulthood in ALSPAC, and sought replication of the findings in four independent European cohorts. After accounting for multiple testing, we found that humidity was associated with faster increase in SBP and DBP in childhood, slower decrease in SBP in adulthood and slower increase in DBP in adolescence. Temperature was associated with slower increase in SBP and DBP in childhood and faster increase in DBP in adolescence. PM_2.5_ and PM_10_ in different trimesters of pregnancy were associated with faster increase in DBP in childhood and slower increase in adolescence. Little evidence for sex differences in these associations was observed. Analyses in independent cohorts replicated results for associations of humidity and temperature with change in blood pressure in childhood. Most of the results for the association of air pollution with blood pressure change did not replicate. The one exception was replication of the association between PM_10_ in the first trimester of pregnancy and slower increase in DBP in childhood.

Many studies have assessed the association of outdoor temperature with blood pressure,^7^ but less evidence is available for humidity. In adults, humidity has been positively 11 associated with blood pressure levels,^34^ and a positive association with DBP has also been observed in children.^14^ We were able to identify only one study which assessed the association of humidity during pregnancy with offspring blood pressure. This study, which included 1,277 European children did not find an association between prenatal humidity and blood pressure in children aged 6-11 years (mean difference per IQR increase in humidity: -0.76 mmHg, 95%CI -3.3, 1.77 for SBP, and -0.44 mmHg, 95%CI -2,79, 1.92 for DBP).^12^

Findings from the same study cited above showed that outdoor temperature during pregnancy was associated with higher SBP and DBP, though CI for the latter spanned the null (mean difference per IQR increase in temperature: 1.6 mmHg, 95%CI: 0.2, 2.9, and 1.2 mmHg, 95%CI: -0.1, 2.4, respectively).^12^ In 4,279 European children aged 4-5 years the association of prenatal temperature with SBP and DBP was more imprecise and CIs included zero (mean difference per IQR increase in temperature: 1.16 mmHg, 95%CI: -0.09, 2.41, and 0.45 mmHg, 95%CI: -0.68, 1.58, respectively).^14^ Our results corroborate these findings and extend research in those studies by showing that higher prenatal temperature was associated with slower increase in SBP and DBP in childhood. This slower change in blood pressure during childhood could suggest that these associations might no longer be evident in the long-term, or that inverse associations between prenatal temperature and blood pressure might be observed in older ages. A study in China with 6,158 individuals (mean age 40 years, SD 9.0) showed an inverse association between temperature during pregnancy and blood pressure in adulthood.^35^ Those born between April and August (and therefore with longer exposure to cold spells during pregnancy) had higher DBP (and also SBP in males only) than those born between September and March.^35^

Outdoor temperature and relative humidity are often correlated (r= -0.50 in our study), and associations with blood pressure were similar when these exposures were mutually adjusted (Supplementary Table 14). The mechanisms underlying the association between outdoor temperature and blood pressure are not fully understood, but it is suggested that lower ambient temperature activates the sympathetic nervous system and increases vasoconstriction leading to increase in heart rate and blood pressure.^7,34^ However, the mechanisms through which outdoor temperature (and humidity) during pregnancy might influence offspring blood pressure require further investigation.

Although overall prenatal air pollution-related factors were associated with faster increase in DBP in childhood, these findings did not replicate in the independent cohorts, except for the association of PM_10_ in the first trimester of pregnancy and slower increase in DBP in childhood. It is possible that these contrasting results are driven by the different 12 levels of exposure and different confounding structure across the cohorts; e.g., air pollution levels are higher in GenR than in the other studies, and the association between socioeconomic factors and air pollution differed across cohorts. Previous studies have shown that prenatal exposure to PM_2.5_ and PM_10_ was associated with higher blood pressure in newborns,^18,36^ particularly exposure in the third trimester of pregnancy.^18^ Prenatal NO_2_ was associated with higher DBP in children aged 4-5 years (mean difference per IQR increase in NO_2_: 0.85 mmHg, 95%CI: 0.38, 1.33), but less evidence was observed for PM_2.5_ and PM_10_ (mean difference per IQR increase in PM_2.5_ and PM_10_: 0.45, 95%CI -0.11, 1.02, and -0.08, 95%CI -0,76, 0.61, respectively).^14^ There are numerous postulated biological mechanisms linking prenatal exposure to air pollution to children’s adverse cardiometabolic health, including direct placental translocation of ultrafine particles, placental and systemic maternal oxidative stress and inflammation elicited by both fine and ultrafine particulate matter, epigenetic changes, and potential endocrine effects.^37^

In our study, most associations with change in blood pressure were observed in childhood. It is possible that any effects of prenatal environmental exposures on blood pressure trajectory are no longer evident at later ages. However, it is important to highlight that contemporaneous exposure to urban environmental factors can affect blood pressure levels,^5,6,9-11,37^ and the long-term exposure to adverse environments might affect blood pressure trajectories across the life course.

Very few studies have explored whether the association between prenatal urban environment and child’s health differs by sex. A study with 822 American children aged 4-6 years showed that associations between prenatal exposure to air pollution and child’s DBP was stronger in females.^17^ In adults, there is suggestion that the association between air pollution and blood pressure is stronger in males.^5^ In our study, we found little evidence of sex differences in the associations between prenatal environmental exposures and blood pressure trajectories.

### Strengths and limitations

We explored the longitudinal association between urban environment in pregnancy and offspring blood pressure using repeated data spanning from childhood to early adulthood. We used an ExWAS approach to assess the association of a range of urban environmental exposures with blood pressure trajectories, taking account of multiple testing and attempting to replicate findings in independent cohorts were possible, in order to minimise chance findings. Our analyses included all participants with at least one blood pressure measure, 13 under the missing at random assumption (i.e. the probability that an outcome value is missing depends on observed values of the outcome, conditional on the covariates in the model), which may have reduced selection bias due to missing outcome data. Violation of this assumption could bias our results, but when we repeated analyses only including those with at least 3 measures we found similar results. We restricted the analyses to those with complete data on the exposures and confounders, which gives unbiased estimates if having complete data is independent of the outcome, after taking the covariates into consideration.^27^ It is not possible to test this assumption in our data, but having complete data is likely to depend on SBP (Supplementary Table 15). Whilst analyses for DBP are likely to be unbiased, analyses for SBP might be overestimated, since those included in the analysis had a higher increase in SBP in childhood. Misclassification is expected to be differential across exposures and different sample sizes might result in different statistical power. The age range of the cohorts included in the replication differed, and although we attempted replication of the results found in ALSPAC, power was limited in some age ranges. The lack of some of our pre-defined confounders in some of the replication cohorts might have influenced the replication results.

### Conclusions

Using an ExWAS approach to systematically assess a range of prenatal urban environmental exposures, this study showed that prenatal outdoor temperature and humidity potentially modulate blood pressure trajectories, particularly in childhood. Our study contributes to the growing body of evidence on the longitudinal associations of prenatal environmental exposures with blood pressure later in life.

## Data Availability

Researchers can apply to use ALSPAC data, including the variables under investigation in this study. Data access information is provided here: http://www.bristol.ac.uk/alspac/researchers/access/

## Abbreviations

ALSPAC: the Avon Longitudinal Study of Parents and Children
CVD: cardiovascular disease
DBP: diastolic blood pressure
EDEN: Etude des Déterminants pré et post natals précoces du développement psychomoteur et de la santé de l’Enfant
ExWAS: exposome-wide association study
IQR: interquartile range
NFBC1986: Northern Finland Birth Cohort 1986
PANIC: the Physical Activity and Nutrition in Children
SBP: systolic blood pressure
SD: standard deviation
UK: United Kingdom

## Acknowledgements

We are extremely grateful to all the families who took part in this study, the midwives for their help in recruiting them, and the whole ALSPAC team, which includes interviewers, computer and laboratory technicians, clerical workers, research scientists, volunteers, managers, receptionists and nurses.

The Generation R Study is conducted by the Erasmus MC, University Medical Center Rotterdam in close collaboration with the School of Law and Faculty of Social Sciences of the Erasmus University Rotterdam, the Municipal Health Service Rotterdam area, Rotterdam, the Rotterdam Homecare Foundation, Rotterdam and the Stichting Trombosedienst & Artsenlaboratorium Rijnmond (STAR-MDC), Rotterdam. We gratefully acknowledge the contribution of children and parents, general practitioners, hospitals, midwives and pharmacies in Rotterdam.

The authors thank the EDEN mother-child cohort study group, whose members are I. Annesi-Maesano, J.Y. Bernard, J. Botton, M.A. Charles, P. Dargent-Molina, B. de Lauzon-Guillain, P. Ducimetière, M. de Agostini, B. Foliguet, A. Forhan, X. Fritel, A. Germa, V. Goua, R. Hankard, B. Heude, M. Kaminski, B. Larroquey, N. Lelong, J. Lepeule, G. Magnin, L. Marchand, C. Nabet, F Pierre, R. Slama, M.J. Saurel-Cubizolles, M. Schweitzer, and O. Thiebaugeorges.

We are grateful to all children and their parents and caregivers who have participated in the PANIC Study. We are also indebted to all members of the PANIC research team for their invaluable contribution in the acquisition of the data throughout the study.

We also thank all NFBC1986 members and researchers who participated in the study.

We also wish to acknowledge the work of the NFBC project centre.

## References

1. Olsen MH, Angell SY, Asma S et al. A call to action and a lifecourse strategy to address the global burden of raised blood pressure on current and future generations: the Lancet Commission on hypertension. Lancet. 2016;388(10060):2665–2712.

2. Zhou B, Perel P, Mensah GA, Ezzati M. Global epidemiology, health burden and effective interventions for elevated blood pressure and hypertension. Nat Rev Cardiol. 2021;18(11):785–802.

3. Chen X, Wang Y. Tracking of blood pressure from childhood to adulthood: a systematic review and meta-regression analysis. Circulation. 2008;117(25):3171–80.

4. Yang L, Magnussen CG, Yang L, Bovet P, Xi B. Elevated blood pressure in childhood or adolescence and cardiovascular outcomes in adulthood: a systematic review. Hypertension. 2020;75(4):948–955.

5. Yang B-Y, Qian Z, Howard SW et al. Global association between ambient air pollution and blood pressure: A systematic review and meta-analysis. Environ Pollut. 2018;235:576–588.

6. van Kempen EE, Kruize H, Boshuizen HC, Ameling CB, Staatsen BAM, de Hollander AEM. The association between noise exposure and blood pressure and ischemic heart disease: a meta-analysis. Environ Health Perspect. 2002;110(3):307–317.

7. Wang Q, Li C, Guo Y et al. Environmental ambient temperature and blood pressure in adults: A systematic review and meta-analysis. Sci Total Environ. 2017;575:276–286.

8. Schulz M, Romppel M, Grande G. Built environment and health: a systematic review of studies in Germany. J Public Health. 2018;40(1):8–15.

9. Bijnens EM, Nawrot TS, Loos RJ et al. Blood pressure in young adulthood and residential greenness in the early-life environment of twins. Environ Health. 2017;16(1):53.

10. Sanders AP, Saland JM, Wright RO, Satlin L. Perinatal and childhood exposure to environmental chemicals and blood pressure in children: a review of literature 2007-2017. Pediatr Res. 2018;84(2):165–180.

11. Dzhambov AM, Dimitrova DD. Children’s blood pressure and its association with road traffic noise exposure - A systematic review with meta-analysis. Environ Res. 2017;152:244–255.

12. Warembourg C, Maitre L, Tamayo-Uria I et al. Early-Life Environmental Exposures and Blood Pressure in Children. J Am Coll Cardiol. 2019;74(10):1317–1328.

13. Zhang M, Mueller NT, Wang H, Hong X, Appel LJ, Wang X. Maternal Exposure to Ambient Particulate Matter _≤_2.5 µm During Pregnancy and the Risk for High Blood Pressure in Childhood. Hypertension. 2018;72:194–201.

14. Warembourg C, Nieuwenhuijsen M, Ballester F et al. Urban environment during early-life and blood pressure in young children. Environ Int. 2021;146:106174.

15. Gluckman PD, Hanson MA, Cooper C, Thornburg KL. Effect of in utero and early-life conditions on adult health and disease. N Engl J Med. 2008;359(1):61–73.

16. Poore KR, Hanson MA, Faustman EM, Neira M. Avoidable early life environmental exposures. Lancet Planet Health. 2017;1(5):e172–e173.

17. Ni Y, Szpiro AA, Young MT et al. Associations of Pre- and Postnatal Air Pollution Exposures with Child Blood Pressure and Modification by Maternal Nutrition: A Prospective Study in the CANDLE Cohort. Environ Health Perspect. 2021;129(4):47004.

18. van Rossem L, Rifas-Shiman SL, Melly SJ et al. Prenatal air pollution exposure and newborn blood pressure. Environ Health Perspect. 2015;123(4):353–9.

19. Boyd A, Golding J, Macleod J et al. Cohort Profile: the ‘children of the 90s’--the index offspring of the Avon Longitudinal Study of Parents and Children. Int J Epidemiol. 2013;42(1):111–27.

20. Fraser A, Macdonald-Wallis C, Tilling K et al. Cohort Profile: the Avon Longitudinal Study of Parents and Children: ALSPAC mothers cohort. Int J Epidemiol. 2013;42(1):97–110.

21. Northstone K, Lewcock M, Groom A et al. The Avon Longitudinal Study of Parents and Children (ALSPAC): an update on the enrolled sample of index children in 2019. Wellcome Open Res. 2019;4:51.

22. Jaddoe VWV, Felix JF, Andersen AN et al. The LifeCycle Project-EU Child Cohort Network: a federated analysis infrastructure and harmonized data of more than 250,000 children and parents. Eur J Epidemiol. 2020;35(7):709–724.

23. Pinot de Moira A, Haakma S, Strandberg-Larsen K et al. The EU Child Cohort Network’s core data: establishing a set of findable, accessible, interoperable and re-usable (FAIR) variables. Eur J Epidemiol. 2021;36(5):565–580.

24. Gulliver J, Elliott P, Henderson J et al. Local- and regional-scale air pollution modelling (PM10) and exposure assessment for pregnancy trimesters, infancy, and childhood to age 15years: Avon Longitudinal Study of Parents And Children (ALSPAC). Environ Int. 2018;113:10–19.

25. Santos S, Maitre L, Warembourg C et al. Applying the exposome concept in birth cohort research: a review of statistical approaches. Eur J Epidemiol. 2020;35(3):193–204.

26. Elhakeem A, Hughes RA, Tilling K et al. Using linear and natural cubic splines, SITAR, and latent trajectory models to characterise nonlinear longitudinal growth trajectories in cohort studies. BMC Med Res Methodol. 2022;22(1):68.

27. Hughes RA, Heron J, Sterne JAC, Tilling K. Accounting for missing data in statistical analyses: multiple imputation is not always the answer. Int J Epidemiol. 2019;48(4):1294–1304.

28. Leckie G, Charlton C. runmlwin: A Program to Run the MLwiN Multilevel Modeling Software from within Stata. J Stat Softw. 2013;52(11):1–40.

29. Ronkainen J, Nedelec R, Atehortua A et al. LongITools: Dynamic longitudinal exposome trajectories in cardiovascular and metabolic noncommunicable diseases. Environmental Epidemiology. 2022;6(1):e184.

30. Heude B, Forhan A, Slama R et al. Cohort Profile: The EDEN mother-child cohort on the prenatal and early postnatal determinants of child health and development. Int J Epidemiol. 2016;45(2):353–63.

31. Kooijman MN, Kruithof CJ, van Duijn CM et al. The Generation R Study: design and cohort update 2017. Eur J Epidemiol. 2016;31(12):1243–1264.

32. Viitasalo A, Eloranta AM, Lintu N et al. The effects of a 2-year individualized and family-based lifestyle intervention on physical activity, sedentary behavior and diet in children. Prev Med. 2016;87:81–88.

33. Miettola S, Hartikainen AL, Vaarasmaki M et al. Offspring’s blood pressure and metabolic phenotype after exposure to gestational hypertension in utero. Eur J Epidemiol. 2013;28(1):87–98.

34. van den Hurk K, de Kort WL, Deinum J, Atsma F. Higher outdoor temperatures are progressively associated with lower blood pressure: a longitudinal study in 100,000 healthy individuals. J Am Soc Hypertens. 2015;9(7):536–43.

35. Li N, Cai L, Heizhati M et al. Maternal exposure to cold spells during pregnancy is associated with higher blood pressure and hypertension in offspring later in life. J Clin Hypertens (Greenwich). 2020;22(10):1884–1891.

36. Madhloum N, Nawrot TS, Gyselaers W et al. Neonatal blood pressure in association with prenatal air pollution exposure, traffic, and land use indicators: An ENVIRONAGE birth cohort study. Environ Int. 2019;130:104853.

37. Johnson NM, Hoffmann AR, Behlen JC et al. Air pollution and children’s health-a review of adverse effects associated with prenatal exposure from fine to ultrafine particulate matter. Environ Health Prev Med. 2021;26(1):72.

