## Supplementary_material for "Prenatal urban environment and blood pressure trajectories from childhood to early adulthood"

**Supplementary Table 9.** Associations between urban environmental exposures and changes in systolic blood pressure (SBP) in ALSPAC, restricted to those with 3 or more blood pressure measurements. 35

**Supplementary methods**

*Environmental exposures*

The environmental exposures were derived as part of the LifeCycle Project. The protocol for the exposures in the urban environment can be found in <https://lifecycle-project.eu/wp-content/uploads/2021/07/Protocol_v4_2021_06_25.pdf>.

A Geographic Information System (GIS) environment for all study areas within the LifeCycle Project was set up at ISGLOBAL, in Barcelona, for centralised processing of the data, in collaboration with each of the cohorts involved. Details on geocoding and linkage to third party physical environment records in the Avon Longitudinal Study of Parents and Children (ALSPAC) have been previously published.[1] The residential geocodes of the participants address history were transferred to a central database held in ISGLOBAL.

Air pollution

For air pollution, exposure estimates for nitrogen dioxide ($NO_2$) and particulate matter < 2.5 µm ($PM_{2.5}$) were based on the land use regression (LUR) modelling approach developed in the ELAPSE Project.[2] Temporal adjustment was conducted using background routine monitoring stations. Temporally adjusted exposure levels to each pollutant were estimated by combining the LUR spatial estimates of pollutants for their geocode with a temporal adjusting factor obtained from the routine monitoring data. Specifically, the ratio of the concentration of the routine monitor of each day of the study period and the annual average during 2010 as the adjustment factor for that day were used. When data on a specific pollutant were not available from the routine network we did a back-extrapolation based on $NO_2$. Data on $NO_2$ was obtained from routine background stations.

For particulate matter < 10 µm ($PM_{10}$), a detailed description is available elsewhere.[3] ADMS-Urban model was used to estimate local traffic and non-traffic daily PM10 within the study area. The NAME-III air pollution model (Numerical Atmospheric-dispersion Modelling Environment) was used to estimate regional/long-range sources (i.e. outside the study area) of daily $PM_{10}$ using meteorological data taken from the ERA-Interim meteorological reanalysis produced by European Centre for Medium-Range Weather Forecasts (ECMWF) and pollution data in the form of daily average concentrations of $PM_{10}$ for 14 receptor locations. A constant was included in the model to account for local non-anthropogenic sources of PM10 (e.g. wind-blown soil and other crustal matter). Exposure estimates were assigned based on home address locations and averaged address-time-weighted $PM_{10}$ exposure estimates in each trimester and the whole pregnancy.[3]

Noise

Noise exposure was based on road traffic noise maps from the UK Department of Environment, Food and Rural Affairs (2006), which were generated under EC Directive 2002/49/EC (Assessment and Management of Environmental Noise) in the framework of the European Noise Directive (END). The primary noise indicators were Lden and Lnight, and these were obtained by doing an intersection between noise map and geocodes. Lden is the long-term average indicator designed to assess annoyance and defined by the END. It refers to an annual average of day, evening and night period of exposure. Lnight is the long-term average indicator designed to assess sleep disturbance and defined by the END. It refers to an annual average of night period of exposure.

Traffic

Traffic assessment was done using local layers of traffic from the UK Department of Environment, Food and Rural Affairs (DEFRA) from 2000. Inverse distance weighted (IDW) interpolation was used to predict values for any unmeasured locations. The inverse distance to nearest road was calculated.

Meteorological

The Long Ashton meteorological station was used to obtain all daily meteorological measurements for ALSPAC; this was part of the European Climate Assessment & Dataset project (ECAD). The meteorological data (temperature and relative humidity) were assigned to participants' geocodes inside a 50km buffer around the station. The average mean relative humidity, average mean temperature, average minimum temperature and average maximum temperature were calculated.

Built environment

Population density was characterised according to the Global Human Settlement Layer[4] from 1990, and corresponds to the number of inhabitants per square kilometre. Values were obtained by doing an intersection between population density grid maps and geocodes.

Building density was created based on the European Settlement Map 2017 and satellite images from 2010 to 2013, and corresponds to the sum of the built area divided by the buffer area around the residential address (both 100 and 300 metres were estimated). The values were generated as an intersection between a buffer geocode and the raster.

Street connectivity corresponds to the street network, which was obtained using the 2012 NAVTEQ. It was measured using the intersection density, which was defined as the number of intersections (that are not dead-ends) inside a buffer of 100 and 300 metres, divided by the area in square kilometres of each buffer. A higher value indicates more intersections and a greater degree of connectivity enabling more direct travel between two points using existing streets and pathways.

Facilities were all points of interest for pedestrians as part of their daily life activities, like restaurants, shops, medical centres, schools, libraries etc, and were obtained using the 2012 NAVTEQ. A total of 100 different subcategories, grouped in 17 categories, were available in the NAVTEQ database. All categories, except Crossings, Auto Services and Parking were included. Facility richness index corresponds to the number of different facility types present divided by the maximum potential number of facility types specified in a buffer of 300 metres, and it ranges from 0 to 1. Facility density index corresponds to the number of facilities present divided by the area of 300 metres buffer (number of facilities/$km^2$). For both facility richness and facility density index, a higher value indicates a higher availability of different facility types.

Land use mix corresponds to the diversity of land use withing a given area, and it was derived from the Shannon's Evenness Index, using Urban Atlas database from 2006. The Shannon's Evenness Index is the degree of mixing of different types of land uses (such as residential, commercial, entertainment, and office development). A higher value indicates a more even distribution of land between the different types of land uses.

Main land use gives a percentage of all types of land use within an area of a buffer of 300 metres from each geocode. The following main land use categories were created, by grouping the land use categories available in the selected databases: "high density residential", "low density residential", "very low density residential", "industrial, commercial, public, military and private units", "transports", "port areas", "airport areas", "other", "urban green", "agricultural green", "natural green", "water".

Accessibility corresponds to public transport network and stops, and were obtained from Open Street Maps in 2019. Public transportation network density was calculated as meters of public transport lines (only bus lines) inside each 300 and 500 metre buffer, divided by the buffer area in square kilometres.

Walkability index was developed to quantify how "walkable" was a buffer of 300 metres around each geocode. The index included four components capturing differences in the physical environment (Land Use Shannon's Evenness Index; facility richness, population density, and connectivity index). Each component was converted to deciles before calculations. A higher value indicates a more walkable environment.

Natural spaces

Normalized Difference Vegetation Index (NDVI) was used to quantify vegetation by measuring the difference between near-infrared (which vegetation strongly reflects) and red light (which vegetation absorbs). NDVI derived from the Landsat 4–5 Thematic Mapper (TM), Landsat 7 Enhanced Thematic Mapper Plus (ETM+), and Landsat 8 Operational Land Imager (OLI)/Thermal Infrared Sensor (TIRS) with 30m x 30m resolution, from 1990, was used to determine the surrounding greenness. The imagery had been selected according to the following criteria: i) cloud cover less than 10 %, ii) Standard Terrain Correction (Level 1T) and iii) greenest period of the year. Two or more images were selected. Surrounding greenness was abstracted as the average of NDVI in buffers of 100, 300 and 500 meters around each geocode. Negative values in the images have been reclassified to null values. Furthermore, an indicator for residential proximity to major green spaces was created, and distance to the nearest green or blue major spaces and the area of this space were calculated. The Europe-wide “Urban Atlas” (prepared by European Environmental Protection Agency) from 2006 was used to extract maps of urban and natural green and blue spaces.

<u>Unhealthy food environment</u>

Unhealthy food environment exposure was generated based on the 2012 NAVTEQ database. The subcategories of facilities related to unhealthy food were selected among the 100 different subcategories in the NAVTEQ database. These were Auto Maintenance, Service, and Petrol (Petrol/Gasoline Station); Entertainment (Bar or Pub); Restaurants (Coffee Shop or Restaurant); Shopping (Convenience Store or Shopping). The unhealthy food environment corresponds to the number of unhealthy facilities present divided by the area of 300 meters buffer (number of facilities/$m^2$). A higher value indicates a higher availability of different unhealthy facilities.

*Environmental exposures in replication cohorts*

Environmental exposure data from GenR and EDEN were generated as part of the LifeCycle Project, and therefore identical methods to those used in ALSPAC were used for these cohorts. Details on the sources of data used can be found in the LifeCycle protocol <https://lifecycle-project.eu/wp-content/uploads/2021/07/Protocol_v4_2021_06_25.pdf>

In PANIC, measures of outdoor temperature, humidity and air pollution were available from the Finnish Meteorological Institute (FMI, https://en.ilmatieteenlaitos.fi/open-data). The participant’s address at baseline was used as proxy of the address at birth. All the participants were linked to the same meteorological station close to the city centre of the city of Kuopio, the University of Eastern Finland and the Kuopio University Hospital.

In NFBC1986, measures of outdoor temperatures and humidity were available from the FMI. Using the Finnish Population Register, the addresses of the participants at birth were retrieved and whenever this was missing, the first address known during the first year of the child was selected. These geographical coordinates were then linked to the closest meteorological station and average temperature and humidity related to the month of birth, or the first month when an address was retrieved, were calculated.

*Blood pressure (BP) measurement*

In ALSPAC, a Dinamap 9300 Vital Signs Monitor was used at the 3-year clinic; a Dinamap 9301 Vital Signs Monitor (Morton Medical, London) was used at the 4-, 5-, 7-, 9-, and 11-year clinics; an Omron MI-5 was used at the 10-year clinic; a Dinamap 8100 Vital Signs Monitor (Morton Medical) was used at the 13-year clinic; and an Omron IntelliSense M6 (Omron Healthcare, Kyoto, Japan) was used at the 15-, and 18-year clinics; and an Omron 705 IT (Omron Electronic Components Europe BV) was used at the 24-year clinic. At age 24, study data were collected and managed using REDCap electronic data capture tools hosted at the University of Bristol.[5] REDCap (Research Electronic Data Capture) is a secure, web-based software platform designed to support data capture for research studies.

In EDEN, blood pressure was measured at 3 and 5 years of age by trained investigators following a standardised measurement protocol. SBP and DBP were measured using the COLIN 8800 oscillometer while the child was lying down after a five-minute rest period, three times spaced two minutes apart; and the last two measurements were averaged. The first measurement was discarded because a white coat effect was observed.

In GenR, child blood pressure was measured at the right brachial artery, four times with one-minute intervals, using a validated automatic sphygmomanometer Datascope Accutor Plus (Paramus, NJ, USA) with an appropriate cuff. We calculated the mean value for systolic (SBP) and diastolic blood pressure (DBP) using the last three measurements of each participant.

In PANIC, blood pressure was measured at baseline (6–8 years of age), at 2-year follow-up (8–10 years of age) and at 8-year follow-up (15–17 years of age). A research nurse measured SBP and DBP from the right arm using the Heine Gamma® G7 aneroid sphygmomanometer (Heine Optotechnik, Herrsching, Germany) to accuracy of 2 mmHg. The measurement protocol included, after a rest of 5 minutes, three measurements in the sitting position at 2-minute intervals. The mean of all three values was used.

In NFBC1986, blood pressure was measured with the participant sitting on a chair after 15 minutes of rest, a cuff of the appropriate size was placed on his/her right upper arm. Blood pressure was measured with an automatic oscillometer blood pressure meter (Omron 705CP). The measure was repeated 2 minutes later; a third measure was done if the second measure failed. If this failed, the nurse used a mercury sphygmomanometer. The mean of first and second measures (or first and third measures, if second failed) were used.

*Confounders*

The confounders used in this study were harmonised as part of the LifeCycle Project. More information on the harmonisation protocol can be found in <https://lifecycle-project.eu/for-scientists/guides-manuals/>.

Maternal level of education was based on the highest ongoing or completed education and classified as low, medium and high according to the International Standard Classification of Education 97/2011 (ISCED-97/2011). High education corresponded to short cycle tertiary, Bachelor, Masters, Doctoral or equivalent (ISCED2011: 5-8, ISCED-97: 5-6); medium education corresponded to upper secondary, or post-secondary non-tertiary (ISCED-2011: 3-4, ISCED-97: 3-4); and low education corresponded to no education; early childhood; pre-primary; primary; lower secondary or second stage of basic education (ISCED-2011: 0-2, ISCED-97: 0-2).

Maternal age at delivery was calculated from the mother's date of birth (obtained at enrolment) and the date of delivery. Ethnicity was self-reported at enrolment and recoded into White and non-White, since 97.5% of women were White.

Area deprivation in ALSPAC was based on data from the Ministry of Housing, Communities & Local Government (UK Government) and combined weighted information from seven domains to produce an overall relative measure of deprivation. The domains and their weights were the following: income deprivation (22.5%), employment deprivation (22.5%), education, skills and training deprivation (13.5%), health deprivation and disability (13.5%), crime (9.3%), barriers to housing and services (9.3%), and living environment deprivation (9.3%). It was categorised in quintiles, where the first corresponds to the least deprived and the last to the most deprived.

*Confounders used in the replication analysis*

We tried to adjust the replication analyses for the same set of covariates used in the main analysis. However, not all cohorts had information for all confounders. In EDEN and GenR, all confounders were available. In PANIC, analyses were adjusted for sex, maternal education, maternal age at birth, and maternal ethnicity. In NFBC1986, analyses were adjusted for sex, maternal education and maternal age at birth.

In EDEN, given the different levels in the exposures and outcomes in both cities where the data were collected (Nancy and Poitiers), the cohort was stratified by city.

*Selection of linear splines knots*

We fitted the best-fitting curve for SBP and DBP trajectories using fractional polynomials to visualise the possible knots for the linear splines. The best-fitting function was selected from a family of flexible polynomial functions. In brief, with simple (single-level) linear regression, the model deviance of each of eight powers (–2, –1, –0.5, 0, 0.5, 1, 2, 3) was used to identify the best-fitting single polynomial. All possible combinations of pairs of these polynomials were then examined, and again the model deviance was used to select the best-fitting model containing two powers (the one with the lowest deviance). For both SBP and DBP models, the best-fitting curve had both powers of 3.

We aimed to fit knots which would represent three developmental periods: childhood, adolescence and emerging adulthood. The location was selected by comparing the Akaike Information Criterion (AIC) and Bayesian Information Criterion (BIC) of different models with knots placed around ages 10 and 18 years. The best model had knots placed at 9.9 and 17.9 years (Supplementary Figure 2).

**References**

1. Boyd A, Thomas R, Hansell AL et al. Data Resource Profile: The ALSPAC birth cohort as a platform to study the relationship of environment and health and social factors. Int J Epidemiol. 2019;48(4):1038-1039k.
2. de Hoogh K, Chen J, Gulliver J et al. Spatial PM(2.5), NO(2), O(3) and BC models for Western Europe - Evaluation of spatiotemporal stability. Environ Int. 2018;120:81-92.
3. Gulliver J, Elliott P, Henderson J et al. Local- and regional-scale air pollution modelling (PM10) and exposure assessment for pregnancy trimesters, infancy, and childhood to age 15years: Avon Longitudinal Study of Parents And Children (ALSPAC). Environ Int. 2018;113:10-19.
4. European Commission, Joint Research Centre; Columbia University, Center for International Earth Science Information Network (2015): GHS population grid, derived from GPW4, multitemporal (1975, 1990, 2000, 2015). European Commission, Joint Research Centre (JRC) [Dataset] PID: http://data.europa.eu/89h/jrc-ghsl-ghs_pop_gpw4_globe_r2015a.
5. Harris PA, Taylor R, Thielke R, Payne J, Gonzalez N, Conde JG. Research electronic data capture (REDCap)--a metadata-driven methodology and workflow process for providing translational research informatics support. J Biomed Inform. 2009;42(2):377-81.

**Supplementary Table 1.** Description of the urban environmental exposures assessed in the Avon Longitudinal Study of Parents and Children (ALSPAC)

| **Type of exposure** | **Variable name** | **Definition (unit)** | **Type of variable** |
|---|---|---|---|
| Noise | lden_c | Average noise exposure of day, evening and night period (Lden, dB) | Categorical |
| | ln_c | Average noise exposure of night period (Lnight, dB) | Categorical |
| Traffic | distinvnear1 | Inverse distance to nearest road (1/m) | Continuous |
| Meteorological | hum | Relative humidity (%) | Continuous |
| | tm | Mean temperature (Celsius) | Continuous |
| | tmax | Maximum temperature (Celsius) | Continuous |
| | tmin | Minimum temperature (Celsius) | Continuous |
| Built environment | bdens100 | Building density within 100m buffer ($m^2$ built/$km^2$) | Continuous |
| | bdens300 | Building density within 300m buffer ($m^2$ built/$km^2$) | Continuous |
| | connind100 | Connectivity within 100m buffer (N intersections/$km^2$) | Continuous |
| | connind300 | Connectivity within 300m buffer (N intersections/$km^2$) | Continuous |
| | fdensity300 | Facility density within 300m buffer (N facilities/$km^2$) | Continuous |
| | frichness300 | Facility richness index within 300m buffer | Continuous |
| | landuseshan300 | Land use within 300m buffer (%) | Continuous |
| | agrgr | Agricultural area within 300m buffer (%) | Continuous |
| | airpt | Airport within 300m buffer (%) | Binary |
| | hdres | Continuous urban fabric within 300m buffer (%) | Continuous |
| | indtr | Industrial, commercial, public, military and private units within 300m buffer (%) | Continuous |
| | ldres | Discontinuous low density urban fabric within 300m buffer (%) | Continuous |

| | | | |
|---|---|---|---|
| | natgr | Forests within 300m buffer (%) | Categorical |
| | other | Mineral extraction and dump sites, constructions within 300m buffer (%) | Categorical |
| | port | Port areas within 300m buffer (%) | Binary |
| | trans | Transport networks and other constructed hard-surfaced areas within 300m buffer (%) | Continuous |
| | urbgr | Green urban areas within 300m buffer (%) | Continuous |
| | vldres | Discontinuous very low density urban fabric within 300m buffer (%) | Binary |
| | water | Water land use within 300m buffer (%) | Binary |
| | popdens | Population density (inhabitants/$km^2$) | Continuous |
| | bus_lines_300 | Length of bus lines in 300m buffer (m/$km^2$) | Continuous |
| | bus_lines_500 | Length of bus lines in 500m buffer (m/$km^2$) | Continuous |
| | walkability_mean | Walkability index | Continuous |
| Natural spaces | blue_dist | Distance to closest blue space (m) | Continuous |
| | blue_size_quint | Area of closest blue space >5,000$m^2$ (quintiles) | Categorical |
| | blueyn300 | Existence of blue space within 300m buffer | Binary |
| | green_dist | Distance to closest green space >5,000$m^2$ (m) | Continuous |
| | green_size | Area of closest green space >5,000$m^2$ ($m^2$) | Continuous |
| | greenyn300 | Existence of green space >5,000$m^2$ within 300m buffer | Binary |
| | ndvi100 | Average greenness within 100m buffer (normalized difference vegetation index [NDVI]) | Categorical |
| | ndvi300 | Average greenness within 300m buffer (NDVI) | Categorical |
| | ndvi500 | Average greenness within 500m buffer (NDVI) | Categorical |

| | | | |
|---|---|---|---|
| Food environment | foodenvdens300 | Food facilities density within 300m buffer (number of facilities related to unhealthy food/$km^2$) | Continuous |
| Air pollution | no2 | Nitrogen dioxide ($NO_2$, $\mu g/m^3$) | Continuous |
| | pm25 | Particulate matter <2.5µm ($PM_{2.5}$, $\mu g/m^3$) | Continuous |
| | pm10 | Particulate matter <10µm ($PM_{10}$, $\mu g/m^3$) | Continuous |
| | pm10_t1 | $PM_{10}$ in the 1st trimester of pregnancy ($\mu g/m^3$) | Continuous |
| | pm10_t2 | $PM_{10}$ in the 2nd trimester of pregnancy ($\mu g/m^3$) | Continuous |
| | pm10_t3 | $PM_{10}$ in the 3rd trimester of pregnancy ($\mu g/m^3$) | Continuous |

**Supplementary Table 2.** Baseline characteristics of the ALSPAC participants included (n=7,454) and not included in the analysis (n=1,780) due to missing data on one or more of the confounders

| Variable | Included | Not included | p-value |
|---|---|---|---|
| | N (%) or mean (SD) | | |
| Sex | | | 0.593 |
| Males | 3,708 (49.7%) | 898 (50.4%) | |
| Females | 3,746 (50.3%) | 882 (49.5%) | |
| Maternal education | | | <0.001 |
| Low | 1,080 (14.5%) | 289 (22.7%) | |
| Medium | 5,237 (70.3%) | 837 (65.9%) | |
| High | 1,137 (15.3%) | 145 (11.4%) | |
| Ethnicity | | | 0.023 |
| White | 7,302 (98.0%) | 1,204 (96.9%) | |
| Non-White | 152 (2.0%) | 38 (3.1%) | |
| Area deprivation | | | <0.001 |
| 1st quintile (least deprived) | 1,434 (19.2%) | 105 (11.4%) | |
| 2nd quintile | 1,391 (18.7%) | 110 (12.0%) | |
| 3rd quintile | 1,677 (22.5%) | 179 (19.5%) | |
| 4th quintile | 1,462 (19.6%) | 207 (22.5%) | |
| 5th quintile (most deprived) | 1,490 (20.0%) | 318 (34.6%) | |
| Age of mother at birth (years), mean (SD) | 28.9 (4.6) | 28.4 (5.1) | 0.005 |

**Supplementary Table 3.** Distribution of age, systolic and diastolic blood pressure in each assessment in the ALSPAC participants included in the analysis (n=7,454)

| Assessment | N | Age (years)<br>Mean (SD) | SBP (mmHg)<br>Mean (SD) | DBP (mmHg)<br>Mean (SD) |
|---|---|---|---|---|
| 1 | 874 | 3.1 (0.02) | 90.2 (8.02) | 56.0 (6.29) |
| 2 | 835 | 4.1 (0.03) | 94.7 (7.83) | 57.4 (6.12) |
| 3 | 800 | 5.2 (0.06) | 98.8 (7.88) | 59.7 (6.45) |
| 4 | 6,056 | 7.5 (0.18) | 98.7 (9.11) | 56.4 (6.61) |
| 5 | 5,811 | 9.9 (0.31) | 102.7 (9.27) | 57.4 (6.37) |
| 6 | 5,522 | 10.6 (0.25) | 104.2 (9.02) | 60.2 (7.91) |
| 7 | 5,387 | 11.7 (0.23) | 105.5 (9.50) | 58.7 (6.36) |
| 8 | 5,185 | 12.8 (0.23) | 111.4 (9.64) | 56.7 (7.85) |
| 9 | 4,093 | 15.4 (0.33) | 122.8 (10.93) | 67.4 (8.77) |
| 10 | 3,585 | 17.7 (0.37) | 116.5 (9.86) | 64.1 (5.86) |
| 11 | 3,070 | 24.4 (0.79) | 115.9 (11.40) | 66.8 (8.01) |

DBP: diastolic blood pressure; SBP: systolic blood pressure; SD: standard deviation

**Supplementary Table 4.** Distribution of the urban environmental exposures in ALSPAC

| Environmental exposure | N (%) or Median (IQR) |
|---|---|
| Lden (dB) | |
| <55 | 1,561 (30.6%) |
| 55-59.9 | 2,397 (47.0%) |
| 60-64.9 | 1,046 (20.5%) |
| 65-69.9 | 75 (1.5%) |
| ≥70 | 16 (0.3%) |
| Lnight (dB) | |
| <55 | 3,463 (68.0%) |
| 55-59.9 | 1,446 (28.4%) |
| 60-64.9 | 161 (3.2%) |
| ≥65 | 25 (0.5%) |
| Inverse distance to nearest road (inverse m), median (IQR) | 0.06 (0.03) |
| Humidity (relative %), median (IQR) | 81.9 (2.0) |
| Mean temperature (Celsius), median (IQR) | 9.5 (2.4) |
| Building density within 100m buffer ($m^2$ built/$km^2$), median (IQR) | 450000 (180000) |
| Building density within 300m buffer ($m^2$ built/$km^2$), median (IQR) | 410000 (180000) |
| Connectivity within 100m buffer (N intersections/$km^2$), median (IQR) | 130 (90) |
| Connectivity within 300m buffer (N intersections/$km^2$), median (IQR) | 120 (70) |
| Facility density within 300m buffer (N facilities/$km^2$), median (IQR) | 18 (32) |
| Land use within 300m buffer (%), median (IQR) | 0.44 (0.13) |
| Agricultural area within 300m buffer (%), median (IQR) | 0 (4) |
| Airport within 300m buffer (%) | |
| 0 | 7,202 (99.4%) |
| >0 | 44 (0.6%) |
| Continuous Urban Fabric within 300m buffer (%) | |
| ≤1 | 6,007 (82.9%) |
| 2-5 | 578 (8.0%) |
| 6-9 | 251 (3.5%) |
| 10-14 | 131 (1.8%) |
| 15+ | 279 (3.9%) |
| Industrial, commercial, public, military and private units within 300m buffer (%), median (IQR) | 6 (10) |
| Discontinuous low density urban fabric within 300m buffer (%), median (IQR) | 64 (24) |
| Forests within 300m buffer (%) | |
| 0 | 6,607 (91.2%) |
| 1-4 | 232 (3.2%) |
| 5-10 | 227 (3.1%) |
| >10 | 180 (2.5%) |
| Mineral extraction and dump sites, constructions within 300m buffer (%) | |
| 0 | 5,934 (81.9%) |
| 1-4 | 927 (12.8%) |
| 5-10 | 385 (5.3%) |
| Port areas within 300m buffer (%) | |
| 0 | 7225 (99.7%) |

| | |
|---|---|
| >0 | 21 (0.3%) |
| Transport networks and other constructed hard-surfaced areas within 300m buffer (%), median (IQR) | 10 (4) |
| Green urban areas within 300m buffer (%), median (IQR) | 6 (12) |
| Discontinuous very low density urban fabric within 300m buffer (%) | |
| 0 | 6,826 (94.2%) |
| >0 | 420 (5.8%) |
| Population density (inhabitants/km$^2$), median (IQR) | 4300 (4300) |
| Bus lines in 300m buffer (m/km$^2$), median (IQR) | 2900 (2000) |
| Bus lines in 500m buffer (m/km$^2$), median (IQR) | 2800 (1900, 3700) |
| Walkability index, median (IQR) | 0.28 (0.08) |
| Distance to closest blue space (m), median (IQR) | 1300 (1200) |
| Area of closest blue space >5,000m$^2$ (m$^2$) | |
| 1st quintile (lowest) | 1,563 (21.6%) |
| 2nd quintile | 1,464 (20.2%) |
| 3rd quintile | 1,754 (24.2%) |
| 4th quintile | 1,411 (19.5%) |
| 5th quintile (highest) | 1,048 (14.5%) |
| Existence of blue space within 300m buffer | |
| No | 6,769 (93.5%) |
| Yes | 474 (6.5%) |
| Distance to closest green space >5,000m$^2$ (m), median (IQR) | 162.17 (215.61) |
| Area of closest green space >5,000m$^2$ (m$^2$), median (IQR) | 40000 (120000) |
| Existence of green space >5,000m$^2$ within 300m buffer | |
| No | 1,761 (24.3%) |
| Yes | 5,482 (75.7%) |
| Average greenness within 100m buffer (NDVI) | |
| 0.2 | 567 (7.6%) |
| 0.3 | 1,808 (24.3%) |
| 0.4 | 3,506 (47.0%) |
| 0.5 | 1,248 (16.7%) |
| 0.6 | 291 (3.9%) |
| 0.7 | 34 (0.5%) |
| Average greenness within 300m buffer (NDVI) | |
| 0.2 | 316 (4.2%) |
| 0.3 | 1,230 (16.5%) |
| 0.4 | 3,724 (50.0%) |
| 0.5 | 1,736 (23.3%) |
| 0.6 | 395 (5.3%) |
| 0.7 | 53 (0.7%) |
| Average greenness within 500m buffer (NDVI) | |
| 0.2 | 227 (3.0%) |
| 0.3 | 973 (13.1%) |
| 0.4 | 3,606 (48.4%) |
| 0.5 | 2,119 (28.4%) |
| 0.6 | 466 (6.3%) |
| 0.7 | 63 (0.8%) |

| | |
|---|---|
| Unhealthy food facilities density within 300m buffer (N facilities/$km^2$), median (IQR) | 4 (7) |
| $NO_2$ (μg/$m^3$), median (IQR) | 27.2 (5.2) |
| $PM_{2.5}$ (μg/$m^3$), median (IQR) | 13.3 (1.1) |
| $PM_{10}$ (μg/$m^3$), median (IQR) | 20.7 (3.8) |
| $PM_{10}$ in the 1st trimester of pregnancy (μg/$m^3$), median (IQR) | 20.8 (8.0) |
| $PM_{10}$ in the 2nd trimester of pregnancy (μg/$m^3$), median (IQR) | 20.0 (7.8) |
| $PM_{10}$ in the 3rd trimester of pregnancy (μg/$m^3$), median (IQR) | 18.4 (6.9) |

**Supplementary Table 5.** Adjusted mean trajectories of systolic and diastolic blood pressure from childhood to early adulthood in ALSPAC

| | **Mean (95%CI)** |
|---|---|
| **Systolic blood pressure (SBP)** | |
| Mean SBP at 3y, mmHg | 94.3 (89.9, 92.7) |
| Change in SBP in childhood (3 to 10y), mmHg/y | 1.56 (1.50, 1.62) |
| Mean SBP at 10y, mmHg | 101.8 (101.1, 102.5) |
| Change in SBP in adolescence (10 to 18y), mmHg/y | 2.57 (2.53, 2.61) |
| Mean SBP at 18y, mmHg | 122.3 (121.6, 123.1) |
| Change in SBP in early adulthood (18 to 24y), mmHg/y | -1.02 (-1.08, -0.97) |
| Ma at 24y, mmHg | 116.2 (115.5, 117.0) |
| **Diastolic blood pressure (DBP)** | |
| DBP at 3y, mmHg | 54.9 (53.9, 55.9) |
| Change in DBP in childhood (3 to 10y), mmHg/y | 0.17 (0.12, 0.21) |
| DBP at 10y, mmHg | 55.8 (55.3, 56.3) |
| Change in DBP in adolescence (10 to 18y), mmHg/y | 1.06 (1.03, 1.09) |
| Blood pressure at 18y, mmHg | 64.3 (63.8, 64.7) |
| Change in DBP in early adulthood (18 to 24y), mmHg/y | 0.14 (0.09, 0.18) |
| Blood pressure at 24y, mmHg | 65.1 (64.6, 65.6) |

Adjusted for maternal education, age at delivery, ethnicity, area deprivation, and sex. All covariates used in the adjustment were set to the mean value or the reference category: maternal education (high), age at delivery (28.9 years), ethnicity (White), area deprivation (least deprived), and sex (male).

**Supplementary Table 6.** Associations between urban environmental exposures and changes in systolic blood pressure (SBP) in ALSPAC

| Exposure | Parameter | N | N obs | beta (95% CI) | p-value |
|---|---|---|---|---|---|
| Lden | mean difference in SBP at age 3y (mmHg) | 5,095 | 28,132 | 0.14 (-0.39, 0.66) | 0.610 |
| | change in SBP in childhood (mmHg/y): 3 to 10y | 5,095 | 28,132 | -0.01 (-0.10, 0.08) | 0.792 |
| | change in SBP in adolescence (mmHg/y): 10 to 18y | 5,095 | 28,132 | 0.02 (-0.04, 0.09) | 0.517 |
| | change in SBP in adulthood (mmHg/y): 18 to 26y | 5,095 | 28,132 | -0.06 (-0.15, 0.03) | 0.170 |
| | change in SBP from childhood to early adulthood | | | | 0.590 |
| Lnight | mean difference in SBP at age 3y (mmHg) | 5,095 | 28,132 | -0.13 (-0.85, 0.59) | 0.715 |
| | change in SBP in childhood (mmHg/y): 3 to 10y | 5,095 | 28,132 | 0.03 (-0.09, 0.16) | 0.576 |
| | change in SBP in adolescence (mmHg/y): 10 to 18y | 5,095 | 28,132 | 0.05 (-0.04, 0.13) | 0.320 |
| | change in SBP in adulthood (mmHg/y): 18 to 26y | 5,095 | 28,132 | -0.08 (-0.20, 0.05) | 0.218 |
| | change in SBP from childhood to early adulthood | | | | 0.429 |
| Inverse distance to nearest road | mean difference in SBP at age 3y (mmHg) | 7,454 | 41,214 | -0.13 (-0.49, 0.22) | 0.470 |
| | change in SBP in childhood (mmHg/y): 3 to 10y | 7,454 | 41,214 | 0.01 (-0.05, 0.07) | 0.718 |
| | change in SBP in adolescence (mmHg/y): 10 to 18y | 7,454 | 41,214 | 0.01 (-0.04, 0.05) | 0.822 |
| | change in SBP in adulthood (mmHg/y): 18 to 26y | 7,454 | 41,214 | -0.04 (-0.10, 0.02) | 0.198 |
| | change in SBP from childhood to early adulthood | | | | 0.599 |
| Humidity | mean difference in SBP at age 3y (mmHg) | 7,402 | 40,925 | -1.97 (-2.53, -1.41) | 4.73E-12 |
| | change in SBP in childhood (mmHg/y): 3 to 10y | 7,402 | 40,925 | 0.29 (0.20, 0.39) | 9.94E-10 |
| | change in SBP in adolescence (mmHg/y): 10 to 18y | 7,402 | 40,925 | 0.06 (0.00, 0.13) | 0.059 |
| | change in SBP in adulthood (mmHg/y): 18 to 26y | 7,402 | 40,925 | -0.11 (-0.20, -0.03) | 0.010 |
| | change in SBP from childhood to early adulthood | | | | 3.51E-13 |
| Mean temperature | mean difference in SBP at age 3y (mmHg) | 7,413 | 40,988 | 0.89 (0.28, 1.50) | 0.004 |
| | change in SBP in childhood (mmHg/y): 3 to 10y | 7,413 | 40,988 | -0.17 (-0.28, -0.07) | 8.55E-04 |
| | change in SBP in adolescence (mmHg/y): 10 to 18y | 7,413 | 40,988 | 0.01 (-0.06, 0.09) | 0.698 |
| | change in SBP in adulthood (mmHg/y): 18 to 26y | 7,413 | 40,988 | 0.02 (-0.08, 0.12) | 0.653 |
| | change in SBP from childhood to early adulthood | | | | 0.007 |
| Building density in 100m buffer | mean difference in SBP at age 3y (mmHg) | 7,447 | 41,172 | -0.06 (-0.49, 0.36) | 0.768 |
| | change in SBP in childhood (mmHg/y): 3 to 10y | 7,447 | 41,172 | -0.05 (-0.12, 0.02) | 0.201 |
| | change in SBP in adolescence (mmHg/y): 10 to 18y | 7,447 | 41,172 | 0.02 (-0.04, 0.07) | 0.559 |

| | | | | | |
|---|---|---|---|---|---|
| | change in SBP in adulthood (mmHg/y): 18 to 26y | 7,447 | 41,172 | 0.02 (-0.05, 0.09) | 0.535 |
| | change in SBP from childhood to early adulthood | | | | 0.511 |
| Building density in 300m buffer | mean difference in SBP at age 3y (mmHg) | 7,450 | 41,191 | -0.13 (-0.55, 0.30) | 0.559 |
| | change in SBP in childhood (mmHg/y): 3 to 10y | 7,450 | 41,191 | -0.02 (-0.09, 0.05) | 0.529 |
| | change in SBP in adolescence (mmHg/y): 10 to 18y | 7,450 | 41,191 | 0.01 (-0.04, 0.06) | 0.743 |
| | change in SBP in adulthood (mmHg/y): 18 to 26y | 7,450 | 41,191 | 0.02 (-0.05, 0.08) | 0.562 |
| | change in SBP from childhood to early adulthood | | | | 0.823 |
| Connectivity in 100m buffer | mean difference in SBP at age 3y (mmHg) | 7,075 | 38,939 | -0.06 (-0.43, 0.30) | 0.730 |
| | change in SBP in childhood (mmHg/y): 3 to 10y | 7,075 | 38,939 | -0.01 (-0.07, 0.05) | 0.765 |
| | change in SBP in adolescence (mmHg/y): 10 to 18y | 7,075 | 38,939 | -0.02 (-0.06, 0.03) | 0.499 |
| | change in SBP in adulthood (mmHg/y): 18 to 26y | 7,075 | 38,939 | 0.03 (-0.03, 0.09) | 0.308 |
| | change in SBP from childhood to early adulthood | | | | 0.694 |
| Connectivity in 300m buffer | mean difference in SBP at age 3y (mmHg) | 7,432 | 41,071 | 0.11 (-0.30, 0.52) | 0.600 |
| | change in SBP in childhood (mmHg/y): 3 to 10y | 7,432 | 41,071 | -0.05 (-0.12, 0.02) | 0.160 |
| | change in SBP in adolescence (mmHg/y): 10 to 18y | 7,432 | 41,071 | 0.02 (-0.03, 0.07) | 0.442 |
| | change in SBP in adulthood (mmHg/y): 18 to 26y | 7,432 | 41,071 | 0.01 (-0.05, 0.08) | 0.698 |
| | change in SBP from childhood to early adulthood | | | | 0.487 |
| Facility density within 300m buffer | mean difference in SBP at age 3y (mmHg) | 7,453 | 41,213 | -0.03 (-0.28, 0.22) | 0.791 |
| | change in SBP in childhood (mmHg/y): 3 to 10y | 7,453 | 41,213 | -0.02 (-0.07, 0.02) | 0.265 |
| | change in SBP in adolescence (mmHg/y): 10 to 18y | 7,453 | 41,213 | 0.01 (-0.02, 0.04) | 0.395 |
| | change in SBP in adulthood (mmHg/y): 18 to 26y | 7,453 | 41,213 | 0.00 (-0.04, 0.03) | 0.812 |
| | change in SBP from childhood to early adulthood | | | | 0.693 |
| Land use within 300m buffer | mean difference in SBP at age 3y (mmHg) | 7,246 | 39,975 | 0.07 (-0.39, 0.54) | 0.758 |
| | change in SBP in childhood (mmHg/y): 3 to 10y | 7,246 | 39,975 | -0.01 (-0.09, 0.07) | 0.812 |
| | change in SBP in adolescence (mmHg/y): 10 to 18y | 7,246 | 39,975 | -0.02 (-0.08, 0.04) | 0.501 |
| | change in SBP in adulthood (mmHg/y): 18 to 26y | 7,246 | 39,975 | -0.04 (-0.12, 0.03) | 0.260 |
| | change in SBP from childhood to early adulthood | | | | 0.390 |
| Agricultural area within 300m buffer | mean difference in SBP at age 3y (mmHg) | 7,246 | 39,975 | -0.02 (-0.12, 0.08) | 0.689 |
| | change in SBP in childhood (mmHg/y): 3 to 10y | 7,246 | 39,975 | 0.00 (-0.02, 0.02) | 0.943 |
| | change in SBP in adolescence (mmHg/y): 10 to 18y | 7,246 | 39,975 | 0.00 (-0.01, 0.01) | 0.838 |
| | change in SBP in adulthood (mmHg/y): 18 to 26y | 7,246 | 39,975 | 0.00 (-0.02, 0.01) | 0.889 |

| | | | | | |
|---|---|---|---|---|---|
| | change in SBP from childhood to early adulthood | | | | 0.995 |
| Airport within 300m buffer | mean difference in SBP at age 3y (mmHg) | 7,246 | 39,975 | -2.09 (-6.58, 2.40) | 0.362 |
| | change in SBP in childhood (mmHg/y): 3 to 10y | 7,246 | 39,975 | 0.46 (-0.29, 1.21) | 0.226 |
| | change in SBP in adolescence (mmHg/y): 10 to 18y | 7,246 | 39,975 | -0.03 (-0.56, 0.50) | 0.911 |
| | change in SBP in adulthood (mmHg/y): 18 to 26y | 7,246 | 39,975 | 0.17 (-0.47, 0.80) | 0.606 |
| | change in SBP from childhood to early adulthood | | | | 0.579 |
| Continuous Urban Fabric within 300m buffer | mean difference in SBP at age 3y (mmHg) | 7,246 | 39,975 | -0.29 (-0.67, 0.09) | 0.136 |
| | change in SBP in childhood (mmHg/y): 3 to 10y | 7,246 | 39,975 | 0.01 (-0.06, 0.07) | 0.855 |
| | change in SBP in adolescence (mmHg/y): 10 to 18y | 7,246 | 39,975 | 0.01 (-0.03, 0.06) | 0.608 |
| | change in SBP in adulthood (mmHg/y): 18 to 26y | 7,246 | 39,975 | 0.00 (-0.06, 0.06) | 0.972 |
| | change in SBP from childhood to early adulthood | | | | 0.928 |
| Industrial, commercial, public, military and private units within 300m buffer | mean difference in SBP at age 3y (mmHg) | 7,246 | 39,975 | -0.05 (-0.41, 0.31) | 0.780 |
| | change in SBP in childhood (mmHg/y): 3 to 10y | 7,246 | 39,975 | 0.03 (-0.03, 0.09) | 0.335 |
| | change in SBP in adolescence (mmHg/y): 10 to 18y | 7,246 | 39,975 | -0.04 (-0.08, 0.01) | 0.115 |
| | change in SBP in adulthood (mmHg/y): 18 to 26y | 7,246 | 39,975 | -0.01 (-0.07, 0.05) | 0.795 |
| | change in SBP from childhood to early adulthood | | | | 0.330 |
| Discontinuous Low Density Urban Fabric within 300m buffer | mean difference in SBP at age 3y (mmHg) | 7,246 | 39,975 | -0.12 (-0.60, 0.36) | 0.622 |
| | change in SBP in childhood (mmHg/y): 3 to 10y | 7,246 | 39,975 | 0.01 (-0.07, 0.09) | 0.822 |
| | change in SBP in adolescence (mmHg/y): 10 to 18y | 7,246 | 39,975 | 0.01 (-0.05, 0.07) | 0.800 |
| | change in SBP in adulthood (mmHg/y): 18 to 26y | 7,246 | 39,975 | 0.01 (-0.06, 0.09) | 0.743 |
| | change in SBP from childhood to early adulthood | | | | 0.937 |
| Forests within 300m buffer | mean difference in SBP at age 3y (mmHg) | 7,246 | 39,975 | -0.24 (-0.80, 0.33) | 0.408 |
| | change in SBP in childhood (mmHg/y): 3 to 10y | 7,246 | 39,975 | 0.01 (-0.09, 0.10) | 0.901 |
| | change in SBP in adolescence (mmHg/y): 10 to 18y | 7,246 | 39,975 | 0.03 (-0.04, 0.10) | 0.386 |
| | change in SBP in adulthood (mmHg/y): 18 to 26y | 7,246 | 39,975 | -0.01 (-0.10, 0.09) | 0.864 |
| | change in SBP from childhood to early adulthood | | | | 0.794 |
| Mineral extraction and dump sites, constructions within 300m buffer | mean difference in SBP at age 3y (mmHg) | 7,246 | 39,975 | -0.56 (-1.18, 0.07) | 0.079 |
| | change in SBP in childhood (mmHg/y): 3 to 10y | 7,246 | 39,975 | 0.14 (0.03, 0.24) | 0.011 |
| | change in SBP in adolescence (mmHg/y): 10 to 18y | 7,246 | 39,975 | -0.03 (-0.10, 0.05) | 0.526 |
| | change in SBP in adulthood (mmHg/y): 18 to 26y | 7,246 | 39,975 | 0.05 (-0.06, 0.15) | 0.398 |
| | change in SBP from childhood to early adulthood | | | | 0.063 |

| | | | | | |
|---|---|---|---|---|---|
| Port areas within 300m buffer | mean difference in SBP at age 3y (mmHg) | 7,246 | 39,975 | 5.33 (-1.9, 12.56) | 0.149 |
| | change in SBP in childhood (mmHg/y): 3 to 10y | 7,246 | 39,975 | -0.65 (-1.87, 0.56) | 0.291 |
| | change in SBP in adolescence (mmHg/y): 10 to 18y | 7,246 | 39,975 | 0.08 (-0.76, 0.91) | 0.852 |
| | change in SBP in adulthood (mmHg/y): 18 to 26y | 7,246 | 39,975 | -0.14 (-1.54, 1.27) | 0.849 |
| | change in SBP from childhood to early adulthood | | | | 0.751 |
| Transport networks and other constructed hard-surfaced areas within 300m buffer | mean difference in SBP at age 3y (mmHg) | 7,246 | 39,975 | 0.04 (-0.38, 0.45) | 0.862 |
| | change in SBP in childhood (mmHg/y): 3 to 10y | 7,246 | 39,975 | -0.02 (-0.09, 0.05) | 0.500 |
| | change in SBP in adolescence (mmHg/y): 10 to 18y | 7,246 | 39,975 | 0.01 (-0.04, 0.06) | 0.566 |
| | change in SBP in adulthood (mmHg/y): 18 to 26y | 7,246 | 39,975 | -0.02 (-0.08, 0.05) | 0.651 |
| | change in SBP from childhood to early adulthood | | | | 0.882 |
| Green urban areas within 300m buffer | mean difference in SBP at age 3y (mmHg) | 7,246 | 39,975 | 0.35 (-0.06, 0.75) | 0.095 |
| | change in SBP in childhood (mmHg/y): 3 to 10y | 7,246 | 39,975 | -0.02 (-0.09, 0.05) | 0.527 |
| | change in SBP in adolescence (mmHg/y): 10 to 18y | 7,246 | 39,975 | -0.02 (-0.07, 0.03) | 0.393 |
| | change in SBP in adulthood (mmHg/y): 18 to 26y | 7,246 | 39,975 | 0.00 (-0.07, 0.07) | 0.984 |
| | change in SBP from childhood to early adulthood | | | | 0.578 |
| Discontinuous very low density urban fabric within 300m buffer | mean difference in SBP at age 3y (mmHg) | 7,246 | 39,975 | -0.61 (-2.15, 0.92) | 0.433 |
| | change in SBP in childhood (mmHg/y): 3 to 10y | 7,246 | 39,975 | 0.03 (-0.22, 0.29) | 0.797 |
| | change in SBP in adolescence (mmHg/y): 10 to 18y | 7,246 | 39,975 | 0.14 (-0.04, 0.31) | 0.123 |
| | change in SBP in adulthood (mmHg/y): 18 to 26y | 7,246 | 39,975 | -0.13 (-0.36, 0.10) | 0.263 |
| | change in SBP from childhood to early adulthood | | | | 0.322 |
| Population density | mean difference in SBP at age 3y (mmHg) | 7,308 | 40,347 | -0.02 (-0.47, 0.43) | 0.930 |
| | change in SBP in childhood (mmHg/y): 3 to 10y | 7,308 | 40,347 | -0.06 (-0.13, 0.02) | 0.140 |
| | change in SBP in adolescence (mmHg/y): 10 to 18y | 7,308 | 40,347 | 0.02 (-0.03, 0.08) | 0.402 |
| | change in SBP in adulthood (mmHg/y): 18 to 26y | 7,308 | 40,347 | 0.08 (0.01, 0.15) | 0.031 |
| | change in SBP from childhood to early adulthood | | | | 0.032 |
| Bus lines in 300m buffer | mean difference in SBP at age 3y (mmHg) | 6,800 | 37,493 | 0.52 (0.08, 0.97) | 0.022 |
| | change in SBP in childhood (mmHg/y): 3 to 10y | 6,800 | 37,493 | -0.06 (-0.13, 0.02) | 0.131 |
| | change in SBP in adolescence (mmHg/y): 10 to 18y | 6,800 | 37,493 | -0.02 (-0.08, 0.04) | 0.479 |
| | change in SBP in adulthood (mmHg/y): 18 to 26y | 6,800 | 37,493 | -0.03 (-0.11, 0.05) | 0.436 |
| | change in SBP from childhood to early adulthood | | | | 0.124 |
| Bus lines in 500m buffer | mean difference in SBP at age 3y (mmHg) | 7,226 | 39,897 | 0.27 (-0.19, 0.73) | 0.252 |

| | | | | | |
|---|---|---|---|---|---|
| | change in SBP in childhood (mmHg/y): 3 to 10y | 7,226 | 39,897 | -0.05 (-0.13, 0.03) | 0.193 |
| | change in SBP in adolescence (mmHg/y): 10 to 18y | 7,226 | 39,897 | -0.01 (-0.07, 0.05) | 0.803 |
| | change in SBP in adulthood (mmHg/y): 18 to 26y | 7,226 | 39,897 | -0.03 (-0.10, 0.05) | 0.499 |
| | change in SBP from childhood to early adulthood | | | | 0.360 |
| Walkability index | mean difference in SBP at age 3y (mmHg) | 7,224 | 39,832 | 0.04 (-0.37, 0.46) | 0.843 |
| | change in SBP in childhood (mmHg/y): 3 to 10y | 7,224 | 39,832 | -0.04 (-0.11, 0.03) | 0.244 |
| | change in SBP in adolescence (mmHg/y): 10 to 18y | 7,224 | 39,832 | 0.01 (-0.04, 0.06) | 0.829 |
| | change in SBP in adulthood (mmHg/y): 18 to 26y | 7,224 | 39,832 | 0.01 (-0.06, 0.07) | 0.859 |
| | change in SBP from childhood to early adulthood | | | | 0.693 |
| Distance to closest blue space | mean difference in SBP at age 3y (mmHg) | 7,243 | 39,952 | 0.27 (-0.16, 0.70) | 0.221 |
| | change in SBP in childhood (mmHg/y): 3 to 10y | 7,243 | 39,952 | -0.03 (-0.1, 0.05) | 0.467 |
| | change in SBP in adolescence (mmHg/y): 10 to 18y | 7,243 | 39,952 | -0.04 (-0.09, 0.01) | 0.146 |
| | change in SBP in adulthood (mmHg/y): 18 to 26y | 7,243 | 39,952 | 0.06 (-0.02, 0.13) | 0.127 |
| | change in SBP from childhood to early adulthood | | | | 0.172 |
| Size of blue space | mean difference in SBP at age 3y (mmHg) | 7,240 | 39,937 | -0.05 (-0.30, 0.20) | 0.718 |
| | change in SBP in childhood (mmHg/y): 3 to 10y | 7,240 | 39,937 | 0.00 (-0.04, 0.04) | 0.955 |
| | change in SBP in adolescence (mmHg/y): 10 to 18y | 7,240 | 39,937 | 0.00 (-0.03, 0.03) | 0.993 |
| | change in SBP in adulthood (mmHg/y): 18 to 26y | 7,240 | 39,937 | 0.01 (-0.04, 0.05) | 0.788 |
| | change in SBP from childhood to early adulthood | | | | 0.993 |
| Existence blue space within 300m buffer | mean difference in SBP at age 3y (mmHg) | 7,243 | 39,952 | 0.77 (-0.63, 2.17) | 0.282 |
| | change in SBP in childhood (mmHg/y): 3 to 10y | 7,243 | 39,952 | -0.11 (-0.34, 0.13) | 0.383 |
| | change in SBP in adolescence (mmHg/y): 10 to 18y | 7,243 | 39,952 | 0.12 (-0.05, 0.30) | 0.164 |
| | change in SBP in adulthood (mmHg/y): 18 to 26y | 7,243 | 39,952 | -0.13 (-0.35, 0.09) | 0.254 |
| | change in SBP from childhood to early adulthood | | | | 0.468 |
| Distance to closest green space | mean difference in SBP at age 3y (mmHg) | 7,243 | 39,952 | -0.46 (-0.93, 0.01) | 0.053 |
| | change in SBP in childhood (mmHg/y): 3 to 10y | 7,243 | 39,952 | 0.06 (-0.02, 0.14) | 0.127 |
| | change in SBP in adolescence (mmHg/y): 10 to 18y | 7,243 | 39,952 | -0.02 (-0.08, 0.04) | 0.474 |
| | change in SBP in adulthood (mmHg/y): 18 to 26y | 7,243 | 39,952 | 0.04 (-0.04, 0.12) | 0.292 |
| | change in SBP from childhood to early adulthood | | | | 0.354 |
| Size of green space | mean difference in SBP at age 3y (mmHg) | 7,243 | 39,952 | -0.05 (-0.51, 0.42) | 0.843 |
| | change in SBP in childhood (mmHg/y): 3 to 10y | 7,243 | 39,952 | 0.03 (-0.04, 0.11) | 0.391 |

| | | | | | |
|---|---|---|---|---|---|
| | change in SBP in adolescence (mmHg/y): 10 to 18y | 7,243 | 39,952 | -0.03 (-0.09, 0.02) | 0.258 |
| | change in SBP in adulthood (mmHg/y): 18 to 26y | 7,243 | 39,952 | 0.03 (-0.04, 0.11) | 0.373 |
| | change in SBP from childhood to early adulthood | | | | 0.614 |
| Existence green space within 300m buffer | mean difference in SBP at age 3y (mmHg) | 7,243 | 39,952 | 0.66 (-0.13, 1.44) | 0.101 |
| | change in SBP in childhood (mmHg/y): 3 to 10y | 7,243 | 39,952 | -0.09 (-0.22, 0.04) | 0.167 |
| | change in SBP in adolescence (mmHg/y): 10 to 18y | 7,243 | 39,952 | 0.01 (-0.09, 0.10) | 0.917 |
| | change in SBP in adulthood (mmHg/y): 18 to 26y | 7,243 | 39,952 | -0.03 (-0.16, 0.10) | 0.635 |
| | change in SBP from childhood to early adulthood | | | | 0.486 |
| Average greenness within 100m buffer | mean difference in SBP at age 3y (mmHg) | 7,454 | 41,214 | 2.94 (-0.63, 6.52) | 0.107 |
| | change in SBP in childhood (mmHg/y): 3 to 10y | 7,454 | 41,214 | -0.23 (-0.83, 0.36) | 0.441 |
| | change in SBP in adolescence (mmHg/y): 10 to 18y | 7,454 | 41,214 | 0.25 (-0.18, 0.69) | 0.250 |
| | change in SBP in adulthood (mmHg/y): 18 to 26y | 7,454 | 41,214 | 0.07 (-0.49, 0.64) | 0.799 |
| | change in SBP from childhood to early adulthood | | | | 0.584 |
| Average greenness within 300m buffer | mean difference in SBP at age 3y (mmHg) | 7,454 | 41,214 | 2.09 (-1.68, 5.85) | 0.277 |
| | change in SBP in childhood (mmHg/y): 3 to 10y | 7,454 | 41,214 | -0.03 (-0.65, 0.60) | 0.935 |
| | change in SBP in adolescence (mmHg/y): 10 to 18y | 7,454 | 41,214 | 0.04 (-0.41, 0.49) | 0.860 |
| | change in SBP in adulthood (mmHg/y): 18 to 26y | 7,454 | 41,214 | -0.04 (-0.63, 0.55) | 0.906 |
| | change in SBP from childhood to early adulthood | | | | 0.998 |
| Average greenness within 500m buffer | mean difference in SBP at age 3y (mmHg) | 7,454 | 41,214 | 3.03 (-0.79, 6.86) | 0.120 |
| | change in SBP in childhood (mmHg/y): 3 to 10y | 7,454 | 41,214 | -0.19 (-0.82, 0.45) | 0.562 |
| | change in SBP in adolescence (mmHg/y): 10 to 18y | 7,454 | 41,214 | -0.04 (-0.50, 0.42) | 0.856 |
| | change in SBP in adulthood (mmHg/y): 18 to 26y | 7,454 | 41,214 | -0.06 (-0.66, 0.54) | 0.845 |
| | change in SBP from childhood to early adulthood | | | | 0.889 |
| Food facilities density within 300m buffer | mean difference in SBP at age 3y (mmHg) | 7,453 | 41,213 | -0.03 (-0.26, 0.21) | 0.829 |
| | change in SBP in childhood (mmHg/y): 3 to 10y | 7,453 | 41,213 | -0.02 (-0.06, 0.02) | 0.257 |
| | change in SBP in adolescence (mmHg/y): 10 to 18y | 7,453 | 41,213 | 0.01 (-0.01, 0.04) | 0.351 |
| | change in SBP in adulthood (mmHg/y): 18 to 26y | 7,453 | 41,213 | 0.01 (-0.02, 0.05) | 0.505 |
| | change in SBP from childhood to early adulthood | | | | 0.451 |
| $NO_2$ | mean difference in SBP at age 3y (mmHg) | 7,412 | 40,984 | -0.24 (-0.67, 0.19) | 0.271 |
| | change in SBP in childhood (mmHg/y): 3 to 10y | 7,412 | 40,984 | 0.01 (-0.06, 0.08) | 0.793 |
| | change in SBP in adolescence (mmHg/y): 10 to 18y | 7,412 | 40,984 | 0.00 (-0.05, 0.05) | 0.940 |

| | | | | | |
|---|---|---|---|---|---|
| | change in SBP in adulthood (mmHg/y): 18 to 26y | 7,412 | 40,984 | -0.02 (-0.09, 0.04) | 0.468 |
| | change in SBP from childhood to early adulthood | | | | 0.888 |
| $PM_{2.5}$ | mean difference in SBP at age 3y (mmHg) | 7,412 | 40,984 | -0.52 (-0.95, -0.09) | 0.017 |
| | change in SBP in childhood (mmHg/y): 3 to 10y | 7,412 | 40,984 | 0.11 (0.03, 0.18) | 0.004 |
| | change in SBP in adolescence (mmHg/y): 10 to 18y | 7,412 | 40,984 | -0.02 (-0.07, 0.03) | 0.451 |
| | change in SBP in adulthood (mmHg/y): 18 to 26y | 7,412 | 40,984 | -0.04 (-0.11, 0.03) | 0.264 |
| | change in SBP from childhood to early adulthood | | | | 0.018 |
| $PM_{10}$ | mean difference in SBP at age 3y (mmHg) | 7,024 | 38,845 | -0.53 (-0.95, -0.11) | 0.013 |
| | change in SBP in childhood (mmHg/y): 3 to 10y | 7,024 | 38,845 | 0.04 (-0.03, 0.11) | 0.271 |
| | change in SBP in adolescence (mmHg/y): 10 to 18y | 7,024 | 38,845 | 0.05 (0.00, 0.11) | 0.052 |
| | change in SBP in adulthood (mmHg/y): 18 to 26y | 7,024 | 38,845 | -0.01 (-0.09, 0.06) | 0.716 |
| | change in SBP from childhood to early adulthood | | | | 0.004 |
| $PM_{10}$ in the 1st trimester of pregnancy | mean difference in SBP at age 3y (mmHg) | 6,860 | 37,953 | 0.23 (-0.22, 0.68) | 0.322 |
| | change in SBP in childhood (mmHg/y): 3 to 10y | 6,860 | 37,953 | -0.07 (-0.14, 0.01) | 0.090 |
| | change in SBP in adolescence (mmHg/y): 10 to 18y | 6,860 | 37,953 | 0.00 (-0.06, 0.07) | 0.916 |
| | change in SBP in adulthood (mmHg/y): 18 to 26y | 6,860 | 37,953 | 0.01 (-0.08, 0.09) | 0.855 |
| | change in SBP from childhood to early adulthood | | | | 0.355 |
| $PM_{10}$ in the 2nd trimester of pregnancy | mean difference in SBP at age 3y (mmHg) | 6,994 | 38,703 | -0.66 (-1.15, -0.17) | 0.008 |
| | change in SBP in childhood (mmHg/y): 3 to 10y | 6,994 | 38,703 | 0.10 (0.02, 0.19) | 0.013 |
| | change in SBP in adolescence (mmHg/y): 10 to 18y | 6,994 | 38,703 | 0.03 (-0.03, 0.09) | 0.359 |
| | change in SBP in adulthood (mmHg/y): 18 to 26y | 6,994 | 38,703 | -0.04 (-0.12, 0.04) | 0.358 |
| | change in SBP from childhood to early adulthood | | | | 0.015 |
| $PM_{10}$ in the 3rd trimester of pregnancy | mean difference in SBP at age 3y (mmHg) | 6,992 | 38,688 | -1.00 (-1.54, -0.46) | 2.97E-04 |
| | change in SBP in childhood (mmHg/y): 3 to 10y | 6,992 | 38,688 | 0.11 (0.02, 0.20) | 0.017 |
| | change in SBP in adolescence (mmHg/y): 10 to 18y | 6,992 | 38,688 | 0.05 (0.00, 0.11) | 0.066 |
| | change in SBP in adulthood (mmHg/y): 18 to 26y | 6,992 | 38,688 | 0.02 (-0.06, 0.09) | 0.655 |
| | change in SBP from childhood to early adulthood | | | | 7.76E-04 |

Adjusted for maternal education, age at delivery, ethnicity, area deprivation, and sex. Betas correspond to mean change in blood pressure by an interquartile range (IQR) increase in the exposure (for continuous exposures), or by change in category of exposure (for categorical exposures); the IQR and categories for each exposure are presented in Supplementary Table 4.

**Supplementary Table 7.** Associations between urban environmental exposures and changes in diastolic blood pressure (DBP) in ALSPAC

| Exposure | Parameter | N | N obs | beta (95% CI) | p-value |
|---|---|---|---|---|---|
| Lden | mean difference in DBP at age 3y (mmHg) | 5,095 | 28,134 | -0.08 (-0.49, 0.32) | 0.688 |
| | change in DBP in childhood (mmHg/y): 3 to 10y | 5,095 | 28,134 | 0.04 (-0.03, 0.10) | 0.303 |
| | change in DBP in adolescence (mmHg/y): 10 to 18y | 5,095 | 28,134 | -0.03 (-0.07, 0.02) | 0.208 |
| | change in DBP in adulthood (mmHg/y): 18 to 26y | 5,095 | 28,134 | 0.00 (-0.07, 0.07) | 0.946 |
| | change in DBP from childhood to early adulthood | | | | 0.530 |
| Lnight | mean difference in DBP at age 3y (mmHg) | 5,095 | 28,134 | -0.27 (-0.83, 0.30) | 0.355 |
| | change in DBP in childhood (mmHg/y): 3 to 10y | 5,095 | 28,134 | 0.07 (-0.02, 0.17) | 0.130 |
| | change in DBP in adolescence (mmHg/y): 10 to 18y | 5,095 | 28,134 | -0.05 (-0.11, 0.01) | 0.112 |
| | change in DBP in adulthood (mmHg/y): 18 to 26y | 5,095 | 28,134 | 0.03 (-0.06, 0.13) | 0.503 |
| | change in DBP from childhood to early adulthood | | | | 0.316 |
| Inverse distance to nearest road | mean difference in DBP at age 3y (mmHg) | 7,454 | 41,216 | -0.14 (-0.42, 0.14) | 0.325 |
| | change in DBP in childhood (mmHg/y): 3 to 10y | 7,454 | 41,216 | 0.02 (-0.02, 0.07) | 0.341 |
| | change in DBP in adolescence (mmHg/y): 10 to 18y | 7,454 | 41,216 | 0.00 (-0.03, 0.04) | 0.780 |
| | change in DBP in adulthood (mmHg/y): 18 to 26y | 7,454 | 41,216 | -0.04 (-0.09, 0.01) | 0.139 |
| | change in DBP from childhood to early adulthood | | | | 0.342 |
| Humidity | mean difference in DBP at age 3y (mmHg) | 7,402 | 40,927 | -1.57 (-2.01, -1.12) | 5.27E-12 |
| | change in DBP in childhood (mmHg/y): 3 to 10y | 7,402 | 40,927 | 0.34 (0.26, 0.41) | 3.27E-19 |
| | change in DBP in adolescence (mmHg/y): 10 to 18y | 7,402 | 40,927 | -0.16 (-0.20, -0.12) | 5.29E-13 |
| | change in DBP in adulthood (mmHg/y): 18 to 26y | 7,402 | 40,927 | 0.06 (-0.01, 0.13) | 0.115 |
| | change in DBP from childhood to early adulthood | | | | 1.80E-21 |
| Mean temperature | mean difference in DBP at age 3y (mmHg) | 7,413 | 40,990 | 0.16 (-0.32, 0.65) | 0.508 |
| | change in DBP in childhood (mmHg/y): 3 to 10y | 7,413 | 40,990 | -0.09 (-0.17, -0.01) | 0.033 |
| | change in DBP in adolescence (mmHg/y): 10 to 18y | 7,413 | 40,990 | 0.08 (0.03, 0.13) | 0.002 |
| | change in DBP in adulthood (mmHg/y): 18 to 26y | 7,413 | 40,990 | 0.03 (-0.04, 0.11) | 0.390 |
| | change in DBP from childhood to early adulthood | | | | 0.001 |
| Building density in 100m buffer | mean difference in DBP at age 3y (mmHg) | 7,447 | 41,174 | 0.14 (-0.19, 0.48) | 0.408 |
| | change in DBP in childhood (mmHg/y): 3 to 10y | 7,447 | 41,174 | -0.03 (-0.08, 0.03) | 0.363 |
| | change in DBP in adolescence (mmHg/y): 10 to 18y | 7,447 | 41,174 | 0.01 (-0.03, 0.04) | 0.638 |

| | | | | | |
|---|---|---|---|---|---|
| | change in DBP in adulthood (mmHg/y): 18 to 26y | 7,447 | 41,174 | -0.03 (-0.09, 0.02) | 0.249 |
| | change in DBP from childhood to early adulthood | | | | 0.543 |
| Building density in 300m buffer | mean difference in DBP at age 3y (mmHg) | 7,450 | 41,193 | 0.09 (-0.25, 0.42) | 0.606 |
| | change in DBP in childhood (mmHg/y): 3 to 10y | 7,450 | 41,193 | -0.02 (-0.08, 0.03) | 0.460 |
| | change in DBP in adolescence (mmHg/y): 10 to 18y | 7,450 | 41,193 | 0.01 (-0.02, 0.05) | 0.432 |
| | change in DBP in adulthood (mmHg/y): 18 to 26y | 7,450 | 41,193 | -0.03 (-0.08, 0.02) | 0.269 |
| | change in DBP from childhood to early adulthood | | | | 0.645 |
| Connectivity in 100m buffer | mean difference in DBP at age 3y (mmHg) | 7,075 | 38,941 | -0.12 (-0.40, 0.17) | 0.423 |
| | change in DBP in childhood (mmHg/y): 3 to 10y | 7,075 | 38,941 | 0.01 (-0.04, 0.05) | 0.817 |
| | change in DBP in adolescence (mmHg/y): 10 to 18y | 7,075 | 38,941 | -0.02 (-0.05, 0.01) | 0.231 |
| | change in DBP in adulthood (mmHg/y): 18 to 26y | 7,075 | 38,941 | -0.02 (-0.07, 0.02) | 0.358 |
| | change in DBP from childhood to early adulthood | | | | 0.250 |
| Connectivity in 300m buffer | mean difference in DBP at age 3y (mmHg) | 7,432 | 41,073 | 0.03 (-0.30, 0.35) | 0.867 |
| | change in DBP in childhood (mmHg/y): 3 to 10y | 7,432 | 41,073 | -0.01 (-0.06, 0.04) | 0.742 |
| | change in DBP in adolescence (mmHg/y): 10 to 18y | 7,432 | 41,073 | 0.00 (-0.03, 0.03) | 0.928 |
| | change in DBP in adulthood (mmHg/y): 18 to 26y | 7,432 | 41,073 | -0.02 (-0.07, 0.03) | 0.381 |
| | change in DBP from childhood to early adulthood | | | | 0.791 |
| Facility density within 300m buffer | mean difference in DBP at age 3y (mmHg) | 7,453 | 41,215 | 0.10 (-0.09, 0.30) | 0.307 |
| | change in DBP in childhood (mmHg/y): 3 to 10y | 7,453 | 41,215 | -0.02 (-0.06, 0.01) | 0.140 |
| | change in DBP in adolescence (mmHg/y): 10 to 18y | 7,453 | 41,215 | 0.01 (-0.01, 0.03) | 0.319 |
| | change in DBP in adulthood (mmHg/y): 18 to 26y | 7,453 | 41,215 | -0.02 (-0.05, 0.01) | 0.223 |
| | change in DBP from childhood to early adulthood | | | | 0.331 |
| Land use within 300m buffer | mean difference in DBP at age 3y (mmHg) | 7,246 | 39,977 | -0.04 (-0.40, 0.33) | 0.840 |
| | change in DBP in childhood (mmHg/y): 3 to 10y | 7,246 | 39,977 | 0.00 (-0.06, 0.06) | 0.916 |
| | change in DBP in adolescence (mmHg/y): 10 to 18y | 7,246 | 39,977 | 0.02 (-0.02, 0.06) | 0.396 |
| | change in DBP in adulthood (mmHg/y): 18 to 26y | 7,246 | 39,977 | -0.02 (-0.09, 0.04) | 0.431 |
| | change in DBP from childhood to early adulthood | | | | 0.779 |
| Agricultural area within 300m buffer | mean difference in DBP at age 3y (mmHg) | 7,246 | 39,977 | -0.01 (-0.09, 0.07) | 0.759 |
| | change in DBP in childhood (mmHg/y): 3 to 10y | 7,246 | 39,977 | 0.00 (-0.02, 0.01) | 0.724 |
| | change in DBP in adolescence (mmHg/y): 10 to 18y | 7,246 | 39,977 | 0.00 (-0.01, 0.00) | 0.419 |
| | change in DBP in adulthood (mmHg/y): 18 to 26y | 7,246 | 39,977 | 0.01 (0.00, 0.02) | 0.219 |

| | | | | | |
|---|---|---|---|---|---|
| | change in DBP from childhood to early adulthood | | | | 0.579 |
| Airport within 300m buffer | mean difference in DBP at age 3y (mmHg) | 7,246 | 39,977 | -0.73 (-4.30, 2.84) | 0.688 |
| | change in DBP in childhood (mmHg/y): 3 to 10y | 7,246 | 39,977 | 0.32 (-0.27, 0.91) | 0.285 |
| | change in DBP in adolescence (mmHg/y): 10 to 18y | 7,246 | 39,977 | -0.08 (-0.45, 0.28) | 0.665 |
| | change in DBP in adulthood (mmHg/y): 18 to 26y | 7,246 | 39,977 | 0.47 (-0.04, 0.99) | 0.071 |
| | change in DBP from childhood to early adulthood | | | | 0.186 |
| Continuous Urban Fabric within 300m buffer | mean difference in DBP at age 3y (mmHg) | 7,246 | 39,977 | 0.01 (-0.29, 0.30) | 0.969 |
| | change in DBP in childhood (mmHg/y): 3 to 10y | 7,246 | 39,977 | -0.01 (-0.06, 0.04) | 0.605 |
| | change in DBP in adolescence (mmHg/y): 10 to 18y | 7,246 | 39,977 | 0.02 (-0.01, 0.05) | 0.169 |
| | change in DBP in adulthood (mmHg/y): 18 to 26y | 7,246 | 39,977 | -0.05 (-0.10, 0.00) | 0.039 |
| | change in DBP from childhood to early adulthood | | | | 0.205 |
| Industrial, commercial, public, military and private units within 300m buffer | mean difference in DBP at age 3y (mmHg) | 7,246 | 39,977 | -0.20 (-0.48, 0.08) | 0.166 |
| | change in DBP in childhood (mmHg/y): 3 to 10y | 7,246 | 39,977 | 0.04 (-0.01, 0.09) | 0.089 |
| | change in DBP in adolescence (mmHg/y): 10 to 18y | 7,246 | 39,977 | -0.02 (-0.05, 0.01) | 0.114 |
| | change in DBP in adulthood (mmHg/y): 18 to 26y | 7,246 | 39,977 | 0.01 (-0.04, 0.06) | 0.606 |
| | change in DBP from childhood to early adulthood | | | | 0.264 |
| Discontinuous Low Density Urban Fabric within 300m buffer | mean difference in DBP at age 3y (mmHg) | 7,246 | 39,977 | 0.15 (-0.23, 0.52) | 0.444 |
| | change in DBP in childhood (mmHg/y): 3 to 10y | 7,246 | 39,977 | -0.02 (-0.08, 0.04) | 0.577 |
| | change in DBP in adolescence (mmHg/y): 10 to 18y | 7,246 | 39,977 | 0.02 (-0.02, 0.06) | 0.387 |
| | change in DBP in adulthood (mmHg/y): 18 to 26y | 7,246 | 39,977 | -0.01 (-0.07, 0.05) | 0.772 |
| | change in DBP from childhood to early adulthood | | | | 0.843 |
| Forests within 300m buffer | mean difference in DBP at age 3y (mmHg) | 7,246 | 39,977 | 0.08 (-0.36, 0.52) | 0.731 |
| | change in DBP in childhood (mmHg/y): 3 to 10y | 7,246 | 39,977 | -0.03 (-0.10, 0.05) | 0.481 |
| | change in DBP in adolescence (mmHg/y): 10 to 18y | 7,246 | 39,977 | 0.00 (-0.05, 0.05) | 0.901 |
| | change in DBP in adulthood (mmHg/y): 18 to 26y | 7,246 | 39,977 | 0.01 (-0.07, 0.08) | 0.898 |
| | change in DBP from childhood to early adulthood | | | | 0.911 |
| Mineral extraction and dump sites, constructions within 300m buffer | mean difference in DBP at age 3y (mmHg) | 7,246 | 39,977 | -0.26 (-0.75, 0.22) | 0.288 |
| | change in DBP in childhood (mmHg/y): 3 to 10y | 7,246 | 39,977 | 0.05 (-0.03, 0.13) | 0.224 |
| | change in DBP in adolescence (mmHg/y): 10 to 18y | 7,246 | 39,977 | -0.06 (-0.12, -0.01) | 0.022 |
| | change in DBP in adulthood (mmHg/y): 18 to 26y | 7,246 | 39,977 | 0.08 (-0.01, 0.16) | 0.087 |
| | change in DBP from childhood to early adulthood | | | | 0.103 |

| | | | | | |
|---|---|---|---|---|---|
| Port areas within 300m buffer | mean difference in DBP at age 3y (mmHg) | 7,246 | 39,977 | 0.85 (-4.91, 6.62) | 0.772 |
| | change in DBP in childhood (mmHg/y): 3 to 10y | 7,246 | 39,977 | 0.06 (-0.90, 1.01) | 0.908 |
| | change in DBP in adolescence (mmHg/y): 10 to 18y | 7,246 | 39,977 | -0.48 (-1.05, 0.09) | 0.099 |
| | change in DBP in adulthood (mmHg/y): 18 to 26y | 7,246 | 39,977 | 0.27 (-0.84, 1.38) | 0.636 |
| | change in DBP from childhood to early adulthood | | | | 0.374 |
| Transport networks and other constructed hard-surfaced areas within 300m buffer | mean difference in DBP at age 3y (mmHg) | 7,246 | 39,977 | 0.07 (-0.25, 0.39) | 0.665 |
| | change in DBP in childhood (mmHg/y): 3 to 10y | 7,246 | 39,977 | -0.01 (-0.07, 0.04) | 0.608 |
| | change in DBP in adolescence (mmHg/y): 10 to 18y | 7,246 | 39,977 | 0.01 (-0.03, 0.04) | 0.698 |
| | change in DBP in adulthood (mmHg/y): 18 to 26y | 7,246 | 39,977 | -0.02 (-0.07, 0.03) | 0.479 |
| | change in DBP from childhood to early adulthood | | | | 0.869 |
| Green urban areas within 300m buffer | mean difference in DBP at age 3y (mmHg) | 7,246 | 39,977 | -0.04 (-0.36, 0.28) | 0.817 |
| | change in DBP in childhood (mmHg/y): 3 to 10y | 7,246 | 39,977 | 0.03 (-0.03, 0.08) | 0.342 |
| | change in DBP in adolescence (mmHg/y): 10 to 18y | 7,246 | 39,977 | 0.02 (-0.01, 0.06) | 0.239 |
| | change in DBP in adulthood (mmHg/y): 18 to 26y | 7,246 | 39,977 | -0.04 (-0.09, 0.02) | 0.180 |
| | change in DBP from childhood to early adulthood | | | | 0.243 |
| Discontinuous very low density urban fabric within 300m buffer | mean difference in DBP at age 3y (mmHg) | 7,246 | 39,977 | -0.68 (-1.89, 0.53) | 0.268 |
| | change in DBP in childhood (mmHg/y): 3 to 10y | 7,246 | 39,977 | 0.06 (-0.14, 0.26) | 0.528 |
| | change in DBP in adolescence (mmHg/y): 10 to 18y | 7,246 | 39,977 | 0.04 (-0.08, 0.16) | 0.528 |
| | change in DBP in adulthood (mmHg/y): 18 to 26y | 7,246 | 39,977 | -0.18 (-0.36, 0.01) | 0.058 |
| | change in DBP from childhood to early adulthood | | | | 0.236 |
| Population density | mean difference in DBP at age 3y (mmHg) | 7,308 | 40,349 | 0.24 (-0.11, 0.59) | 0.186 |
| | change in DBP in childhood (mmHg/y): 3 to 10y | 7,308 | 40,349 | -0.07 (-0.13, -0.01) | 0.025 |
| | change in DBP in adolescence (mmHg/y): 10 to 18y | 7,308 | 40,349 | 0.02 (-0.02, 0.06) | 0.313 |
| | change in DBP in adulthood (mmHg/y): 18 to 26y | 7,308 | 40,349 | 0.02 (-0.04, 0.08) | 0.460 |
| | change in DBP from childhood to early adulthood | | | | 0.095 |
| Bus lines in 300m buffer | mean difference in DBP at age 3y (mmHg) | 6,800 | 37,495 | 0.06 (-0.29, 0.41) | 0.755 |
| | change in DBP in childhood (mmHg/y): 3 to 10y | 6,800 | 37,495 | 0.01 (-0.05, 0.07) | 0.703 |
| | change in DBP in adolescence (mmHg/y): 10 to 18y | 6,800 | 37,495 | -0.03 (-0.06, 0.01) | 0.188 |
| | change in DBP in adulthood (mmHg/y): 18 to 26y | 6,800 | 37,495 | 0.02 (-0.04, 0.08) | 0.505 |
| | change in DBP from childhood to early adulthood | | | | 0.625 |
| Bus lines in 500m buffer | mean difference in DBP at age 3y (mmHg) | 7,226 | 39,899 | 0.03 (-0.34, 0.39) | 0.892 |

| | | | | | |
|---|---|---|---|---|---|
| | change in DBP in childhood (mmHg/y): 3 to 10y | 7,226 | 39,899 | -0.01 (-0.07, 0.05) | 0.855 |
| | change in DBP in adolescence (mmHg/y): 10 to 18y | 7,226 | 39,899 | -0.02 (-0.06, 0.02) | 0.249 |
| | change in DBP in adulthood (mmHg/y): 18 to 26y | 7,226 | 39,899 | 0.01 (-0.05, 0.08) | 0.671 |
| | change in DBP from childhood to early adulthood | | | | 0.623 |
| Walkability index | mean difference in DBP at age 3y (mmHg) | 7,224 | 39,834 | 0.08 (-0.24, 0.41) | 0.620 |
| | change in DBP in childhood (mmHg/y): 3 to 10y | 7,224 | 39,834 | -0.03 (-0.08, 0.03) | 0.334 |
| | change in DBP in adolescence (mmHg/y): 10 to 18y | 7,224 | 39,834 | 0.01 (-0.02, 0.04) | 0.579 |
| | change in DBP in adulthood (mmHg/y): 18 to 26y | 7,224 | 39,834 | -0.03 (-0.08, 0.02) | 0.291 |
| | change in DBP from childhood to early adulthood | | | | 0.582 |
| Distance to closest blue space | mean difference in DBP at age 3y (mmHg) | 7,243 | 39,954 | 0.21 (-0.13, 0.54) | 0.225 |
| | change in DBP in childhood (mmHg/y): 3 to 10y | 7,243 | 39,954 | -0.03 (-0.09, 0.02) | 0.261 |
| | change in DBP in adolescence (mmHg/y): 10 to 18y | 7,243 | 39,954 | -0.01 (-0.05, 0.02) | 0.500 |
| | change in DBP in adulthood (mmHg/y): 18 to 26y | 7,243 | 39,954 | 0.05 (-0.01, 0.10) | 0.128 |
| | change in DBP from childhood to early adulthood | | | | 0.247 |
| Size of blue space | mean difference in DBP at age 3y (mmHg) | 7,240 | 39,939 | -0.09 (-0.28, 0.11) | 0.375 |
| | change in DBP in childhood (mmHg/y): 3 to 10y | 7,240 | 39,939 | 0.01 (-0.02, 0.04) | 0.469 |
| | change in DBP in adolescence (mmHg/y): 10 to 18y | 7,240 | 39,939 | 0.00 (-0.02, 0.02) | 0.840 |
| | change in DBP in adulthood (mmHg/y): 18 to 26y | 7,240 | 39,939 | -0.01 (-0.04, 0.02) | 0.569 |
| | change in DBP from childhood to early adulthood | | | | 0.807 |
| Existence blue space within 300m buffer | mean difference in DBP at age 3y (mmHg) | 7,243 | 39,954 | 0.46 (-0.63, 1.56) | 0.407 |
| | change in DBP in childhood (mmHg/y): 3 to 10y | 7,243 | 39,954 | -0.07 (-0.25, 0.11) | 0.446 |
| | change in DBP in adolescence (mmHg/y): 10 to 18y | 7,243 | 39,954 | 0.01 (-0.11, 0.13) | 0.847 |
| | change in DBP in adulthood (mmHg/y): 18 to 26y | 7,243 | 39,954 | -0.08 (-0.26, 0.09) | 0.361 |
| | change in DBP from childhood to early adulthood | | | | 0.662 |
| Distance to closest green space | mean difference in DBP at age 3y (mmHg) | 7,243 | 39,954 | 0.03 (-0.34, 0.39) | 0.888 |
| | change in DBP in childhood (mmHg/y): 3 to 10y | 7,243 | 39,954 | 0.00 (-0.06, 0.06) | 0.965 |
| | change in DBP in adolescence (mmHg/y): 10 to 18y | 7,243 | 39,954 | -0.02 (-0.06, 0.02) | 0.428 |
| | change in DBP in adulthood (mmHg/y): 18 to 26y | 7,243 | 39,954 | 0.00 (-0.06, 0.06) | 0.970 |
| | change in DBP from childhood to early adulthood | | | | 0.807 |
| Size of green space | mean difference in DBP at age 3y (mmHg) | 7,243 | 39,954 | 0.09 (-0.27, 0.45) | 0.631 |
| | change in DBP in childhood (mmHg/y): 3 to 10y | 7,243 | 39,954 | 0.03 (-0.03, 0.09) | 0.408 |

| | | | | | |
|---|---|---|---|---|---|
| | change in DBP in adolescence (mmHg/y): 10 to 18y | 7,243 | 39,954 | -0.04 (-0.07, 0.00) | 0.064 |
| | change in DBP in adulthood (mmHg/y): 18 to 26y | 7,243 | 39,954 | 0.04 (-0.02, 0.10) | 0.247 |
| | change in DBP from childhood to early adulthood | | | | 0.305 |
| Existence green space within 300m buffer | mean difference in DBP at age 3y (mmHg) | 7,243 | 39,954 | 0.08 (-0.54, 0.69) | 0.801 |
| | change in DBP in childhood (mmHg/y): 3 to 10y | 7,243 | 39,954 | -0.01 (-0.11, 0.09) | 0.860 |
| | change in DBP in adolescence (mmHg/y): 10 to 18y | 7,243 | 39,954 | 0.03 (-0.04, 0.09) | 0.403 |
| | change in DBP in adulthood (mmHg/y): 18 to 26y | 7,243 | 39,954 | 0.02 (-0.08, 0.13) | 0.652 |
| | change in DBP from childhood to early adulthood | | | | 0.665 |
| Average greenness within 100m buffer | mean difference in DBP at age 3y (mmHg) | 7,454 | 41,216 | 0.56 (-2.23, 3.35) | 0.694 |
| | change in DBP in childhood (mmHg/y): 3 to 10y | 7,454 | 41,216 | -0.24 (-0.70, 0.22) | 0.313 |
| | change in DBP in adolescence (mmHg/y): 10 to 18y | 7,454 | 41,216 | 0.18 (-0.11, 0.48) | 0.218 |
| | change in DBP in adulthood (mmHg/y): 18 to 26y | 7,454 | 41,216 | 0.09 (-0.37, 0.54) | 0.709 |
| | change in DBP from childhood to early adulthood | | | | 0.420 |
| Average greenness within 300m buffer | mean difference in DBP at age 3y (mmHg) | 7,454 | 41,216 | 0.30 (-2.64, 3.24) | 0.841 |
| | change in DBP in childhood (mmHg/y): 3 to 10y | 7,454 | 41,216 | -0.10 (-0.58, 0.38) | 0.676 |
| | change in DBP in adolescence (mmHg/y): 10 to 18y | 7,454 | 41,216 | 0.13 (-0.17, 0.43) | 0.398 |
| | change in DBP in adulthood (mmHg/y): 18 to 26y | 7,454 | 41,216 | 0.10 (-0.37, 0.58) | 0.671 |
| | change in DBP from childhood to early adulthood | | | | 0.686 |
| Average greenness within 500m buffer | mean difference in DBP at age 3y (mmHg) | 7,454 | 41,216 | -0.42 (-3.41, 2.56) | 0.781 |
| | change in DBP in childhood (mmHg/y): 3 to 10y | 7,454 | 41,216 | 0.06 (-0.43, 0.55) | 0.812 |
| | change in DBP in adolescence (mmHg/y): 10 to 18y | 7,454 | 41,216 | -0.04 (-0.35, 0.27) | 0.797 |
| | change in DBP in adulthood (mmHg/y): 18 to 26y | 7,454 | 41,216 | 0.19 (-0.29, 0.67) | 0.442 |
| | change in DBP from childhood to early adulthood | | | | 0.885 |
| Food facilities density within 300m buffer | mean difference in DBP at age 3y (mmHg) | 7,453 | 41,215 | 0.17 (-0.02, 0.35) | 0.074 |
| | change in DBP in childhood (mmHg/y): 3 to 10y | 7,453 | 41,215 | -0.04 (-0.07, -0.01) | 0.021 |
| | change in DBP in adolescence (mmHg/y): 10 to 18y | 7,453 | 41,215 | 0.01 (-0.01, 0.03) | 0.373 |
| | change in DBP in adulthood (mmHg/y): 18 to 26y | 7,453 | 41,215 | -0.01 (-0.03, 0.02) | 0.726 |
| | change in DBP from childhood to early adulthood | | | | 0.148 |
| $NO_2$ | mean difference in DBP at age 3y (mmHg) | 7,412 | 40,986 | -0.14 (-0.48, 0.19) | 0.402 |
| | change in DBP in childhood (mmHg/y): 3 to 10y | 7,412 | 40,986 | 0.03 (-0.03, 0.08) | 0.368 |
| | change in DBP in adolescence (mmHg/y): 10 to 18y | 7,412 | 40,986 | 0.00 (-0.03, 0.04) | 0.882 |

| | | | | | |
|---|---|---|---|---|---|
| | change in DBP in adulthood (mmHg/y): 18 to 26y | 7,412 | 40,986 | -0.05 (-0.10, 0.00) | 0.060 |
| | change in DBP from childhood to early adulthood | | | | 0.187 |
| $PM_{2.5}$ | mean difference in DBP at age 3y (mmHg) | 7,412 | 40,986 | -0.41 (-0.75, -0.07) | 0.017 |
| | change in DBP in childhood (mmHg/y): 3 to 10y | 7,412 | 40,986 | 0.11 (0.06, 0.17) | 8.85E-05 |
| | change in DBP in adolescence (mmHg/y): 10 to 18y | 7,412 | 40,986 | -0.07 (-0.10, -0.03) | 3.27E-04 |
| | change in DBP in adulthood (mmHg/y): 18 to 26y | 7,412 | 40,986 | 0.00 (-0.05, 0.06) | 0.982 |
| | change in DBP from childhood to early adulthood | | | | 3.95E-05 |
| $PM_{10}$ | mean difference in DBP at age 3y (mmHg) | 7,024 | 38,846 | -0.35 (-0.67, -0.02) | 0.036 |
| | change in DBP in childhood (mmHg/y): 3 to 10y | 7,024 | 38,846 | 0.04 (-0.01, 0.10) | 0.125 |
| | change in DBP in adolescence (mmHg/y): 10 to 18y | 7,024 | 38,846 | 0.01 (-0.03, 0.04) | 0.742 |
| | change in DBP in adulthood (mmHg/y): 18 to 26y | 7,024 | 38,846 | -0.07 (-0.13, -0.01) | 0.020 |
| | change in DBP from childhood to early adulthood | | | | 0.038 |
| $PM_{10}$ in the 1st trimester of pregnancy | mean difference in DBP at age 3y (mmHg) | 6,860 | 37,954 | 0.61 (0.26, 0.96) | 7.01E-04 |
| | change in DBP in childhood (mmHg/y): 3 to 10y | 6,860 | 37,954 | -0.10 (-0.16, -0.04) | 8.30E-08 |
| | change in DBP in adolescence (mmHg/y): 10 to 18y | 6,860 | 37,954 | -0.02 (-0.06, 0.03) | 0.428 |
| | change in DBP in adulthood (mmHg/y): 18 to 26y | 6,860 | 37,954 | -0.02 (-0.09, 0.05) | 0.569 |
| | change in DBP from childhood to early adulthood | | | | 4.55E-04 |
| $PM_{10}$ in the 2nd trimester of pregnancy | mean difference in DBP at age 3y (mmHg) | 6,994 | 38,704 | -0.77 (-1.15, -0.39) | 6.22E-05 |
| | change in DBP in childhood (mmHg/y): 3 to 10y | 6,994 | 38,704 | 0.18 (0.12, 0.24) | 1.97E-08 |
| | change in DBP in adolescence (mmHg/y): 10 to 18y | 6,994 | 38,704 | -0.07 (-0.11, -0.03) | 0.001 |
| | change in DBP in adulthood (mmHg/y): 18 to 26y | 6,994 | 38,704 | -0.05 (-0.12, 0.02) | 0.130 |
| | change in DBP from childhood to early adulthood | | | | 6.04E-09 |
| $PM_{10}$ in the 3rd trimester of pregnancy | mean difference in DBP at age 3y (mmHg) | 6,992 | 38,689 | -0.94 (-1.38, -0.51) | 2.01E-05 |
| | change in DBP in childhood (mmHg/y): 3 to 10y | 6,992 | 38,689 | 0.08 (0.01, 0.16) | 0.022 |
| | change in DBP in adolescence (mmHg/y): 10 to 18y | 6,992 | 38,689 | 0.07 (0.03, 0.11) | 2.15E-04 |
| | change in DBP in adulthood (mmHg/y): 18 to 26y | 6,992 | 38,689 | -0.04 (-0.10, 0.02) | 0.161 |
| | change in DBP from childhood to early adulthood | | | | 1.20E-06 |

Adjusted for maternal education, age at delivery, ethnicity, area deprivation, and sex. Betas correspond to mean change in blood pressure by an interquartile range (IQR) increase in the exposure (for continuous exposures), or by change in category of exposure (for categorical exposures); the IQR and categories for each exposure are presented in Supplementary Table 4.

**Supplementary Table 8.** Average predicted systolic and diastolic blood pressure according to different levels of urban environmental exposures in ALSPAC

| Exposure | Percentile/ average level of exposure | Age (years) | Predicted blood pressure (mmHg) Mean (95%CI) |
|---|---|---|---|
| **Systolic blood pressure** | | | |
| Humidity | 25th (81.1%) | 3 | 91.9 (91.1, 92.7) |
| | | 10 | 101.8 (101.1, 102.5) |
| | | 18 | 122.1 (121.3, 122.8) |
| | | 24 | 116.3 (115.5, 117.1) |
| | 75th (83.1%) | 3 | 89.9 (89.1, 90.7) |
| | | 10 | 101.8 (101.1, 102.5) |
| | | 18 | 122.6 (121.9, 123.3) |
| | | 24 | 116.1 (115.3, 116.9) |
| Temperature | 25th (8.3 °C) | 3 | 90.5 (89.7, 91.3) |
| | | 10 | 101.9 (101.2, 102.6) |
| | | 18 | 122.4 (121.7, 123.2) |
| | | 24 | 116.2 (115.5, 117.0) |
| | 75th (10.7 °C) | 3 | 91.4 (90.6, 92.2) |
| | | 10 | 101.6 (100.9, 102.3) |
| | | 18 | 122.2 (121.5, 123.0) |
| | | 24 | 116.2 (115.4, 117.0) |
| $PM_{10}$ in the 3rd trimester of pregnancy | 25th (15.7 μg/m$^3$) | 3 | 91.5 (90.7, 92.3) |
| | | 10 | 102 (101.3, 102.8) |
| | | 18 | 122.3 (121.6, 123.1) |
| | | 24 | 116.1 (115.3, 116.9) |
| | 75th (22.6 μg/m$^3$) | 3 | 90.5 (89.7, 91.3) |
| | | 10 | 101.8 (101.1, 102.5) |
| | | 18 | 122.5 (121.7, 123.3) |
| | | 24 | 116.4 (115.6, 117.2) |
| **Diastolic blood pressure** | | | |
| Humidity | 25th (81.1%) | 3 | 55.3 (54.8, 55.9) |
| | | 10 | 55.4 (54.9, 55.9) |
| | | 18 | 64.5 (64.0, 65.0) |
| | | 24 | 65.2 (64.6, 65.7) |
| | 75th (83.1%) | 3 | 53.8 (53.2, 54.3) |
| | | 10 | 56.2 (55.7, 56.7) |
| | | 18 | 64.0 (63.5, 64.5) |
| | | 24 | 65.0 (64.4, 65.6) |
| Temperature | 25th (8.3 °C) | 3 | 54.5 (54.0, 55.1) |
| | | 10 | 56.0 (55.5, 56.5) |
| | | 18 | 64.2 (63.6, 64.7) |
| | | 24 | 64.9 (64.3, 65.5) |
| | 75th (10.7 °C) | 3 | 54.7 (54.1, 55.3) |
| | | 10 | 55.6 (55.1, 56.1) |
| | | 18 | 64.3 (63.8, 64.9) |
| | | 24 | 65.3 (64.7, 65.9) |
| $PM_{10}$ in the 1st trimester of pregnancy | 25th (17.3 μg/m$^3$) | 3 | 54.1 (53.5, 54.7) |
| | | 10 | 55.8 (55.3, 56.3) |
| | | 18 | 64.3 (63.7, 64.8) |

| | | | |
|---|---|---|---|
| | | 24 | 65.2 (64.6, 65.8) |
| | 75th (25.3 μg/m$^3$) | 3 | 54.7 (54.2, 55.3) |
| | | 10 | 55.7 (55.2, 56.3) |
| | | 18 | 64.1 (63.5, 64.6) |
| | | 24 | 64.9 (64.3, 65.5) |
| $PM_{10}$ in the 2nd trimester of pregnancy | 25th (17.0 μg/m$^3$) | 3 | 54.8 (54.2, 55.4) |
| | | 10 | 55.5 (55.0, 56.0) |
| | | 18 | 64.2 (63.7, 64.7) |
| | | 24 | 65.2 (64.6, 65.8) |
| | 75th (24.8 μg/m$^3$) | 3 | 54.0 (53.4, 54.6) |
| | | 10 | 56.0 (55.5, 56.5) |
| | | 18 | 64.1 (63.6, 64.7) |
| | | 24 | 64.8 (64.2, 65.4) |
| $PM_{10}$ in the 3rd trimester of pregnancy | 25th (15.7 μg/m$^3$) | 3 | 54.9 (54.3, 55.4) |
| | | 10 | 56.0 (55.5, 56.5) |
| | | 18 | 64.1 (63.5, 64.6) |
| | | 24 | 65.0 (64.4, 65.6) |
| | 75th (22.6 μg/m$^3$) | 3 | 53.9 (53.3, 54.6) |
| | | 10 | 55.6 (55.1, 56.1) |
| | | 18 | 64.3 (63.8, 64.8) |
| | | 24 | 65.0 (64.4, 65.5) |

Adjusted for maternal education, age at delivery, ethnicity, area deprivation, and sex. All covariates used in the adjustment were set to the mean value or the reference category: maternal education (high), age at delivery (28.9 years), ethnicity (White), area deprivation (least deprived), and sex (male).

**Supplementary Table 9.** Associations between urban environmental exposures and changes in systolic blood pressure (SBP) in ALSPAC, restricted to those with 3 or more blood pressure measurements

| Exposure | Parameter | N | N obs | beta (95% CI) | p-value |
|---|---|---|---|---|---|
| Lden | mean difference in SBP at age 3y (mmHg) | 4,199 | 26,873 | 0.14 (-0.41, 0.68) | 0.622 |
| | change in SBP in childhood (mmHg/y): 3 to 10y | 4,199 | 26,873 | -0.01 (-0.10, 0.08) | 0.832 |
| | change in SBP in adolescence (mmHg/y): 10 to 18y | 4,199 | 26,873 | 0.02 (-0.04, 0.09) | 0.518 |
| | change in SBP in adulthood (mmHg/y): 18 to 26y | 4,199 | 26,873 | -0.05 (-0.14, 0.04) | 0.299 |
| | change in SBP from childhood to early adulthood | | | | 0.762 |
| Lnight | mean difference in SBP at age 3y (mmHg) | 4,199 | 26,873 | -0.18 (-0.94, 0.58) | 0.642 |
| | change in SBP in childhood (mmHg/y): 3 to 10y | 4,199 | 26,873 | 0.04 (-0.09, 0.16) | 0.549 |
| | change in SBP in adolescence (mmHg/y): 10 to 18y | 4,199 | 26,873 | 0.05 (-0.04, 0.14) | 0.292 |
| | change in SBP in adulthood (mmHg/y): 18 to 26y | 4,199 | 26,873 | -0.06 (-0.18, 0.07) | 0.373 |
| | change in SBP from childhood to early adulthood | | | | 0.476 |
| Inverse distance to nearest road | mean difference in SBP at age 3y (mmHg) | 6,129 | 39,356 | -0.21 (-0.58, 0.16) | 0.265 |
| | change in SBP in childhood (mmHg/y): 3 to 10y | 6,129 | 39,356 | 0.02 (-0.04, 0.08) | 0.585 |
| | change in SBP in adolescence (mmHg/y): 10 to 18y | 6,129 | 39,356 | 0.00 (-0.04, 0.05) | 0.899 |
| | change in SBP in adulthood (mmHg/y): 18 to 26y | 6,129 | 39,356 | -0.04 (-0.10, 0.02) | 0.217 |
| | change in SBP from childhood to early adulthood | | | | 0.585 |
| Humidity | mean difference in SBP at age 3y (mmHg) | 6,087 | 39,081 | -1.97 (-2.55, -1.39) | 2.19E-11 |
| | change in SBP in childhood (mmHg/y): 3 to 10y | 6,087 | 39,081 | 0.29 (0.20, 0.39) | 2.01E-09 |
| | change in SBP in adolescence (mmHg/y): 10 to 18y | 6,087 | 39,081 | 0.05 (-0.01, 0.12) | 0.100 |
| | change in SBP in adulthood (mmHg/y): 18 to 26y | 6,087 | 39,081 | -0.09 (-0.18, -0.01) | 0.034 |
| | change in SBP from childhood to early adulthood | | | | 5.06E-12 |
| Mean temperature | mean difference in SBP at age 3y (mmHg) | 6,096 | 39,141 | 0.93 (0.30, 1.56) | 0.004 |
| | change in SBP in childhood (mmHg/y): 3 to 10y | 6,096 | 39,141 | -0.19 (-0.30, -0.09) | 3.34E-04 |
| | change in SBP in adolescence (mmHg/y): 10 to 18y | 6,096 | 39,141 | 0.04 (-0.04, 0.11) | 0.364 |
| | change in SBP in adulthood (mmHg/y): 18 to 26y | 6,096 | 39,141 | 0.01 (-0.09, 0.11) | 0.889 |
| | change in SBP from childhood to early adulthood | | | | 0.004 |
| Building density in 100m buffer | mean difference in SBP at age 3y (mmHg) | 6,123 | 39,316 | -0.09 (-0.53, 0.35) | 0.701 |
| | change in SBP in childhood (mmHg/y): 3 to 10y | 6,123 | 39,316 | -0.05 (-0.12, 0.03) | 0.221 |

| | | | | | |
|---|---|---|---|---|---|
| | change in SBP in adolescence (mmHg/y): 10 to 18y | 6,123 | 39,316 | 0.02 (-0.04, 0.07) | 0.537 |
| | change in SBP in adulthood (mmHg/y): 18 to 26y | 6,123 | 39,316 | 0.01 (-0.06, 0.08) | 0.716 |
| | change in SBP from childhood to early adulthood | | | | 0.608 |
| Building density in 300m buffer | mean difference in SBP at age 3y (mmHg) | 6,126 | 39,335 | -0.17 (-0.60, 0.27) | 0.457 |
| | change in SBP in childhood (mmHg/y): 3 to 10y | 6,126 | 39,335 | -0.02 (-0.09, 0.05) | 0.549 |
| | change in SBP in adolescence (mmHg/y): 10 to 18y | 6,126 | 39,335 | 0.01 (-0.04, 0.06) | 0.707 |
| | change in SBP in adulthood (mmHg/y): 18 to 26y | 6,126 | 39,335 | 0.01 (-0.05, 0.08) | 0.679 |
| | change in SBP from childhood to early adulthood | | | | 0.875 |
| Connectivity in 100m buffer | mean difference in SBP at age 3y (mmHg) | 5,801 | 37,155 | -0.15 (-0.53, 0.22) | 0.424 |
| | change in SBP in childhood (mmHg/y): 3 to 10y | 5,801 | 37,155 | -0.01 (-0.07, 0.05) | 0.811 |
| | change in SBP in adolescence (mmHg/y): 10 to 18y | 5,801 | 37,155 | -0.01 (-0.06, 0.03) | 0.643 |
| | change in SBP in adulthood (mmHg/y): 18 to 26y | 5,801 | 37,155 | 0.03 (-0.02, 0.09) | 0.260 |
| | change in SBP from childhood to early adulthood | | | | 0.701 |
| Connectivity in 300m buffer | mean difference in SBP at age 3y (mmHg) | 6,108 | 39,215 | 0.01 (-0.42, 0.44) | 0.962 |
| | change in SBP in childhood (mmHg/y): 3 to 10y | 6,108 | 39,215 | -0.04 (-0.11, 0.03) | 0.221 |
| | change in SBP in adolescence (mmHg/y): 10 to 18y | 6,108 | 39,215 | 0.02 (-0.03, 0.07) | 0.396 |
| | change in SBP in adulthood (mmHg/y): 18 to 26y | 6,108 | 39,215 | 0.01 (-0.05, 0.08) | 0.742 |
| | change in SBP from childhood to early adulthood | | | | 0.563 |
| Facility density within 300m buffer | mean difference in SBP at age 3y (mmHg) | 6,129 | 39,356 | 0.00 (-0.26, 0.25) | 0.988 |
| | change in SBP in childhood (mmHg/y): 3 to 10y | 6,129 | 39,356 | -0.03 (-0.07, 0.01) | 0.204 |
| | change in SBP in adolescence (mmHg/y): 10 to 18y | 6,129 | 39,356 | 0.01 (-0.02, 0.04) | 0.507 |
| | change in SBP in adulthood (mmHg/y): 18 to 26y | 6,129 | 39,356 | -0.01 (-0.05, 0.03) | 0.614 |
| | change in SBP from childhood to early adulthood | | | | 0.617 |
| Land use within 300m buffer | mean difference in SBP at age 3y (mmHg) | 5,949 | 38,155 | 0.19 (-0.29, 0.68) | 0.433 |
| | change in SBP in childhood (mmHg/y): 3 to 10y | 5,949 | 38,155 | -0.01 (-0.09, 0.07) | 0.767 |
| | change in SBP in adolescence (mmHg/y): 10 to 18y | 5,949 | 38,155 | -0.03 (-0.08, 0.03) | 0.401 |
| | change in SBP in adulthood (mmHg/y): 18 to 26y | 5,949 | 38,155 | -0.05 (-0.12, 0.03) | 0.250 |
| | change in SBP from childhood to early adulthood | | | | 0.298 |
| Agricultural area within 300m buffer | mean difference in SBP at age 3y (mmHg) | 5,949 | 38,155 | 0.00 (-0.11, 0.10) | 0.941 |
| | change in SBP in childhood (mmHg/y): 3 to 10y | 5,949 | 38,155 | 0.00 (-0.02, 0.02) | 0.892 |
| | change in SBP in adolescence (mmHg/y): 10 to 18y | 5,949 | 38,155 | 0.00 (-0.01, 0.01) | 0.790 |

| | | | | | |
|---|---|---|---|---|---|
| | change in SBP in adulthood (mmHg/y): 18 to 26y | 5,949 | 38,155 | 0.00 (-0.02, 0.01) | 0.881 |
| | change in SBP from childhood to early adulthood | | | | 0.995 |
| Airport within 300m buffer | mean difference in SBP at age 3y (mmHg) | 5,949 | 38,155 | -2.38 (-6.89, 2.13) | 0.301 |
| | change in SBP in childhood (mmHg/y): 3 to 10y | 5,949 | 38,155 | 0.49 (-0.26, 1.24) | 0.199 |
| | change in SBP in adolescence (mmHg/y): 10 to 18y | 5,949 | 38,155 | 0.00 (-0.53, 0.54) | 0.990 |
| | change in SBP in adulthood (mmHg/y): 18 to 26y | 5,949 | 38,155 | 0.15 (-0.49, 0.78) | 0.647 |
| | change in SBP from childhood to early adulthood | | | | 0.513 |
| Continuous Urban Fabric within 300m buffer | mean difference in SBP at age 3y (mmHg) | 5,949 | 38,155 | -0.36 (-0.74, 0.03) | 0.072 |
| | change in SBP in childhood (mmHg/y): 3 to 10y | 5,949 | 38,155 | 0.02 (-0.04, 0.08) | 0.565 |
| | change in SBP in adolescence (mmHg/y): 10 to 18y | 5,949 | 38,155 | 0.00 (-0.04, 0.05) | 0.888 |
| | change in SBP in adulthood (mmHg/y): 18 to 26y | 5,949 | 38,155 | -0.01 (-0.07, 0.06) | 0.769 |
| | change in SBP from childhood to early adulthood | | | | 0.914 |
| Industrial, commercial, public, military and private units within 300m buffer | mean difference in SBP at age 3y (mmHg) | 5,949 | 38,155 | -0.02 (-0.39, 0.35) | 0.904 |
| | change in SBP in childhood (mmHg/y): 3 to 10y | 5,949 | 38,155 | 0.03 (-0.04, 0.09) | 0.418 |
| | change in SBP in adolescence (mmHg/y): 10 to 18y | 5,949 | 38,155 | -0.03 (-0.08, 0.01) | 0.144 |
| | change in SBP in adulthood (mmHg/y): 18 to 26y | 5,949 | 38,155 | -0.01 (-0.07, 0.05) | 0.692 |
| | change in SBP from childhood to early adulthood | | | | 0.366 |
| Discontinuous low density urban fabric within 300m buffer | mean difference in SBP at age 3y (mmHg) | 5,949 | 38,155 | -0.21 (-0.71, 0.28) | 0.398 |
| | change in SBP in childhood (mmHg/y): 3 to 10y | 5,949 | 38,155 | 0.01 (-0.07, 0.09) | 0.797 |
| | change in SBP in adolescence (mmHg/y): 10 to 18y | 5,949 | 38,155 | 0.01 (-0.05, 0.07) | 0.716 |
| | change in SBP in adulthood (mmHg/y): 18 to 26y | 5,949 | 38,155 | 0.02 (-0.06, 0.09) | 0.701 |
| | change in SBP from childhood to early adulthood | | | | 0.885 |
| Forests within 300m buffer | mean difference in SBP at age 3y (mmHg) | 5,949 | 38,155 | -0.29 (-0.88, 0.30) | 0.336 |
| | change in SBP in childhood (mmHg/y): 3 to 10y | 5,949 | 38,155 | 0.01 (-0.08, 0.11) | 0.774 |
| | change in SBP in adolescence (mmHg/y): 10 to 18y | 5,949 | 38,155 | 0.03 (-0.05, 0.10) | 0.492 |
| | change in SBP in adulthood (mmHg/y): 18 to 26y | 5,949 | 38,155 | -0.01 (-0.11, 0.08) | 0.767 |
| | change in SBP from childhood to early adulthood | | | | 0.845 |
| Mineral extraction and dump sites, constructions within 300m buffer | mean difference in SBP at age 3y (mmHg) | 5,949 | 38,155 | -0.47 (-1.11, 0.18) | 0.155 |
| | change in SBP in childhood (mmHg/y): 3 to 10y | 5,949 | 38,155 | 0.15 (0.04, 0.25) | 0.008 |
| | change in SBP in adolescence (mmHg/y): 10 to 18y | 5,949 | 38,155 | -0.04 (-0.12, 0.04) | 0.369 |
| | change in SBP in adulthood (mmHg/y): 18 to 26y | 5,949 | 38,155 | 0.04 (-0.07, 0.15) | 0.429 |

| | | | | | |
|---|---|---|---|---|---|
| | change in SBP from childhood to early adulthood | | | | 0.054 |
| Port areas within 300m buffer | mean difference in SBP at age 3y (mmHg) | 5,949 | 38,155 | 7.15 (-0.66, 14.96) | 0.073 |
| | change in SBP in childhood (mmHg/y): 3 to 10y | 5,949 | 38,155 | -0.90 (-2.18, 0.39) | 0.171 |
| | change in SBP in adolescence (mmHg/y): 10 to 18y | 5,949 | 38,155 | 0.23 (-0.61, 1.08) | 0.590 |
| | change in SBP in adulthood (mmHg/y): 18 to 26y | 5,949 | 38,155 | -0.25 (-1.71, 1.22) | 0.740 |
| | change in SBP from childhood to early adulthood | | | | 0.590 |
| Transport networks and other constructed hard-surfaced areas within 300m buffer | mean difference in SBP at age 3y (mmHg) | 5,949 | 38,155 | -0.08 (-0.51, 0.34) | 0.696 |
| | change in SBP in childhood (mmHg/y): 3 to 10y | 5,949 | 38,155 | -0.01 (-0.08, 0.06) | 0.755 |
| | change in SBP in adolescence (mmHg/y): 10 to 18y | 5,949 | 38,155 | 0.01 (-0.04, 0.06) | 0.637 |
| | change in SBP in adulthood (mmHg/y): 18 to 26y | 5,949 | 38,155 | -0.02 (-0.09, 0.05) | 0.566 |
| | change in SBP from childhood to early adulthood | | | | 0.931 |
| Green urban areas within 300m buffer | mean difference in SBP at age 3y (mmHg) | 5,949 | 38,155 | 0.42 (0.00, 0.84) | 0.050 |
| | change in SBP in childhood (mmHg/y): 3 to 10y | 5,949 | 38,155 | -0.02 (-0.09, 0.05) | 0.509 |
| | change in SBP in adolescence (mmHg/y): 10 to 18y | 5,949 | 38,155 | -0.02 (-0.08, 0.03) | 0.370 |
| | change in SBP in adulthood (mmHg/y): 18 to 26y | 5,949 | 38,155 | 0.00 (-0.07, 0.08) | 0.897 |
| | change in SBP from childhood to early adulthood | | | | 0.567 |
| Discontinuous very low density urban fabric within 300m buffer | mean difference in SBP at age 3y (mmHg) | 5,949 | 38,155 | -0.59 (-2.16, 0.97) | 0.458 |
| | change in SBP in childhood (mmHg/y): 3 to 10y | 5,949 | 38,155 | 0.05 (-0.21, 0.31) | 0.689 |
| | change in SBP in adolescence (mmHg/y): 10 to 18y | 5,949 | 38,155 | 0.13 (-0.05, 0.30) | 0.150 |
| | change in SBP in adulthood (mmHg/y): 18 to 26y | 5,949 | 38,155 | -0.13 (-0.36, 0.10) | 0.274 |
| | change in SBP from childhood to early adulthood | | | | 0.334 |
| Population density | mean difference in SBP at age 3y (mmHg) | 6,002 | 38,515 | -0.02 (-0.49, 0.45) | 0.927 |
| | change in SBP in childhood (mmHg/y): 3 to 10y | 6,002 | 38,515 | -0.06 (-0.14, 0.02) | 0.135 |
| | change in SBP in adolescence (mmHg/y): 10 to 18y | 6,002 | 38,515 | 0.02 (-0.04, 0.07) | 0.584 |
| | change in SBP in adulthood (mmHg/y): 18 to 26y | 6,002 | 38,515 | 0.07 (0.00, 0.14) | 0.067 |
| | change in SBP from childhood to early adulthood | | | | 0.088 |
| Bus lines in 300m buffer | mean difference in SBP at age 3y (mmHg) | 5,590 | 35,800 | 0.46 (-0.01, 0.93) | 0.053 |
| | change in SBP in childhood (mmHg/y): 3 to 10y | 5,590 | 35,800 | -0.05 (-0.13, 0.03) | 0.199 |
| | change in SBP in adolescence (mmHg/y): 10 to 18y | 5,590 | 35,800 | -0.03 (-0.08, 0.03) | 0.372 |
| | change in SBP in adulthood (mmHg/y): 18 to 26y | 5,590 | 35,800 | -0.02 (-0.10, 0.05) | 0.529 |
| | change in SBP from childhood to early adulthood | | | | 0.165 |

| | | | | | |
|---|---|---|---|---|---|
| Bus lines in 500m buffer | mean difference in SBP at age 3y (mmHg) | 5,936 | 38,093 | 0.26 (-0.22, 0.75) | 0.284 |
| | change in SBP in childhood (mmHg/y): 3 to 10y | 5,936 | 38,093 | -0.05 (-0.13, 0.03) | 0.193 |
| | change in SBP in adolescence (mmHg/y): 10 to 18y | 5,936 | 38,093 | -0.01 (-0.07, 0.05) | 0.829 |
| | change in SBP in adulthood (mmHg/y): 18 to 26y | 5,936 | 38,093 | -0.02 (-0.10, 0.06) | 0.663 |
| | change in SBP from childhood to early adulthood | | | | 0.444 |
| Walkability index | mean difference in SBP at age 3y (mmHg) | 5,928 | 38,014 | 0.04 (-0.39, 0.47) | 0.858 |
| | change in SBP in childhood (mmHg/y): 3 to 10y | 5,928 | 38,014 | -0.04 (-0.11, 0.03) | 0.279 |
| | change in SBP in adolescence (mmHg/y): 10 to 18y | 5,928 | 38,014 | 0.00 (-0.05, 0.05) | 0.949 |
| | change in SBP in adulthood (mmHg/y): 18 to 26y | 5,928 | 38,014 | 0.00 (-0.07, 0.06) | 0.941 |
| | change in SBP from childhood to early adulthood | | | | 0.691 |
| Distance to closest blue space | mean difference in SBP at age 3y (mmHg) | 5,946 | 38,132 | 0.34 (-0.10, 0.79) | 0.129 |
| | change in SBP in childhood (mmHg/y): 3 to 10y | 5,946 | 38,132 | -0.04 (-0.11, 0.04) | 0.345 |
| | change in SBP in adolescence (mmHg/y): 10 to 18y | 5,946 | 38,132 | -0.05 (-0.10, 0.01) | 0.103 |
| | change in SBP in adulthood (mmHg/y): 18 to 26y | 5,946 | 38,132 | 0.05 (-0.02, 0.13) | 0.160 |
| | change in SBP from childhood to early adulthood | | | | 0.108 |
| Size of blue space | mean difference in SBP at age 3y (mmHg) | 5,944 | 38,118 | 0.06 (-0.20, 0.32) | 0.668 |
| | change in SBP in childhood (mmHg/y): 3 to 10y | 5,944 | 38,118 | -0.01 (-0.05, 0.04) | 0.796 |
| | change in SBP in adolescence (mmHg/y): 10 to 18y | 5,944 | 38,118 | 0.00 (-0.03, 0.03) | 0.971 |
| | change in SBP in adulthood (mmHg/y): 18 to 26y | 5,944 | 38,118 | 0.01 (-0.03, 0.05) | 0.678 |
| | change in SBP from childhood to early adulthood | | | | 0.968 |
| Existence blue space within 300m buffer | mean difference in SBP at age 3y (mmHg) | 5,946 | 38,132 | 0.69 (-0.77, 2.14) | 0.355 |
| | change in SBP in childhood (mmHg/y): 3 to 10y | 5,946 | 38,132 | -0.08 (-0.32, 0.16) | 0.519 |
| | change in SBP in adolescence (mmHg/y): 10 to 18y | 5,946 | 38,132 | 0.14 (-0.04, 0.32) | 0.120 |
| | change in SBP in adulthood (mmHg/y): 18 to 26y | 5,946 | 38,132 | -0.11 (-0.33, 0.11) | 0.325 |
| | change in SBP from childhood to early adulthood | | | | 0.454 |
| Distance to closest green space | mean difference in SBP at age 3y (mmHg) | 5,946 | 38,132 | -0.52 (-1.00, -0.03) | 0.037 |
| | change in SBP in childhood (mmHg/y): 3 to 10y | 5,946 | 38,132 | 0.06 (-0.02, 0.15) | 0.116 |
| | change in SBP in adolescence (mmHg/y): 10 to 18y | 5,946 | 38,132 | -0.02 (-0.08, 0.03) | 0.420 |
| | change in SBP in adulthood (mmHg/y): 18 to 26y | 5,946 | 38,132 | 0.04 (-0.04, 0.12) | 0.297 |
| | change in SBP from childhood to early adulthood | | | | 0.342 |
| Size of green space | mean difference in SBP at age 3y (mmHg) | 5,946 | 38,132 | -0.06 (-0.53, 0.42) | 0.822 |

| | | | | | |
|---|---|---|---|---|---|
| | change in SBP in childhood (mmHg/y): 3 to 10y | 5,946 | 38,132 | 0.03 (-0.04, 0.11) | 0.396 |
| | change in SBP in adolescence (mmHg/y): 10 to 18y | 5,946 | 38,132 | -0.02 (-0.08, 0.03) | 0.430 |
| | change in SBP in adulthood (mmHg/y): 18 to 26y | 5,946 | 38,132 | 0.03 (-0.05, 0.11) | 0.439 |
| | change in SBP from childhood to early adulthood | | | | 0.727 |
| Existence green space within 300m buffer | mean difference in SBP at age 3y (mmHg) | 5,946 | 38,132 | 0.73 (-0.08, 1.54) | 0.079 |
| | change in SBP in childhood (mmHg/y): 3 to 10y | 5,946 | 38,132 | -0.10 (-0.23, 0.03) | 0.144 |
| | change in SBP in adolescence (mmHg/y): 10 to 18y | 5,946 | 38,132 | 0.01 (-0.09, 0.11) | 0.791 |
| | change in SBP in adulthood (mmHg/y): 18 to 26y | 5,946 | 38,132 | -0.03 (-0.16, 0.10) | 0.634 |
| | change in SBP from childhood to early adulthood | | | | 0.478 |
| Average greenness within 100m buffer | mean difference in SBP at age 3y (mmHg) | 6,129 | 39,356 | 3.28 (-0.40, 6.95) | 0.081 |
| | change in SBP in childhood (mmHg/y): 3 to 10y | 6,129 | 39,356 | -0.25 (-0.85, 0.35) | 0.418 |
| | change in SBP in adolescence (mmHg/y): 10 to 18y | 6,129 | 39,356 | 0.31 (-0.13, 0.74) | 0.172 |
| | change in SBP in adulthood (mmHg/y): 18 to 26y | 6,129 | 39,356 | 0.15 (-0.43, 0.73) | 0.609 |
| | change in SBP from childhood to early adulthood | | | | 0.381 |
| Average greenness within 300m buffer | mean difference in SBP at age 3y (mmHg) | 6,129 | 39,356 | 2.17 (-1.70, 6.04) | 0.272 |
| | change in SBP in childhood (mmHg/y): 3 to 10y | 6,129 | 39,356 | 0.01 (-0.62, 0.64) | 0.976 |
| | change in SBP in adolescence (mmHg/y): 10 to 18y | 6,129 | 39,356 | 0.05 (-0.41, 0.50) | 0.834 |
| | change in SBP in adulthood (mmHg/y): 18 to 26y | 6,129 | 39,356 | 0.03 (-0.57, 0.63) | 0.928 |
| | change in SBP from childhood to early adulthood | | | | 0.993 |
| Average greenness within 500m buffer | mean difference in SBP at age 3y (mmHg) | 6,129 | 39,356 | 3.24 (-0.71, 7.19) | 0.108 |
| | change in SBP in childhood (mmHg/y): 3 to 10y | 6,129 | 39,356 | -0.16 (-0.80, 0.49) | 0.634 |
| | change in SBP in adolescence (mmHg/y): 10 to 18y | 6,129 | 39,356 | -0.04 (-0.51, 0.42) | 0.858 |
| | change in SBP in adulthood (mmHg/y): 18 to 26y | 6,129 | 39,356 | 0.00 (-0.61, 0.61) | 0.998 |
| | change in SBP from childhood to early adulthood | | | | 0.943 |
| Food facilities density within 300m buffer | mean difference in SBP at age 3y (mmHg) | 6,129 | 39,356 | -0.02 (-0.26, 0.23) | 0.899 |
| | change in SBP in childhood (mmHg/y): 3 to 10y | 6,129 | 39,356 | -0.03 (-0.07, 0.01) | 0.201 |
| | change in SBP in adolescence (mmHg/y): 10 to 18y | 6,129 | 39,356 | 0.01 (-0.02, 0.04) | 0.400 |
| | change in SBP in adulthood (mmHg/y): 18 to 26y | 6,129 | 39,356 | 0.01 (-0.03, 0.04) | 0.740 |
| | change in SBP from childhood to early adulthood | | | | 0.541 |
| $NO_2$ | mean difference in SBP at age 3y (mmHg) | 6,095 | 39,137 | -0.23 (-0.68, 0.22) | 0.311 |
| | change in SBP in childhood (mmHg/y): 3 to 10y | 6,095 | 39,137 | 0.01 (-0.06, 0.09) | 0.717 |

| | | | | | |
|---|---|---|---|---|---|
| | change in SBP in adolescence (mmHg/y): 10 to 18y | 6,095 | 39,137 | 0.00 (-0.05, 0.05) | 0.954 |
| | change in SBP in adulthood (mmHg/y): 18 to 26y | 6,095 | 39,137 | -0.02 (-0.09, 0.05) | 0.525 |
| | change in SBP from childhood to early adulthood | | | | 0.897 |
| $PM_{2.5}$ | mean difference in SBP at age 3y (mmHg) | 6,095 | 39,137 | -0.54 (-0.99, -0.10) | 0.017 |
| | change in SBP in childhood (mmHg/y): 3 to 10y | 6,095 | 39,137 | 0.10 (0.02, 0.17) | 0.009 |
| | change in SBP in adolescence (mmHg/y): 10 to 18y | 6,095 | 39,137 | -0.02 (-0.07, 0.04) | 0.529 |
| | change in SBP in adulthood (mmHg/y): 18 to 26y | 6,095 | 39,137 | -0.03 (-0.10, 0.04) | 0.364 |
| | change in SBP from childhood to early adulthood | | | | 0.046 |
| $PM_{10}$ | mean difference in SBP at age 3y (mmHg) | 5,777 | 37,099 | -0.54 (-0.97, -0.10) | 0.015 |
| | change in SBP in childhood (mmHg/y): 3 to 10y | 5,777 | 37,099 | 0.05 (-0.02, 0.12) | 0.194 |
| | change in SBP in adolescence (mmHg/y): 10 to 18y | 5,777 | 37,099 | 0.05 (-0.01, 0.10) | 0.091 |
| | change in SBP in adulthood (mmHg/y): 18 to 26y | 5,777 | 37,099 | 0.00 (-0.08, 0.07) | 0.902 |
| | change in SBP from childhood to early adulthood | | | | 0.052 |
| $PM_{10}$ in the 1$^{st}$ trimester of pregnancy | mean difference in SBP at age 3y (mmHg) | 5,640 | 36,243 | 0.27 (-0.20, 0.73) | 0.267 |
| | change in SBP in childhood (mmHg/y): 3 to 10y | 5,640 | 36,243 | -0.05 (-0.13, 0.03) | 0.223 |
| | change in SBP in adolescence (mmHg/y): 10 to 18y | 5,640 | 36,243 | -0.02 (-0.08, 0.05) | 0.648 |
| | change in SBP in adulthood (mmHg/y): 18 to 26y | 5,640 | 36,243 | 0.01 (-0.07, 0.10) | 0.765 |
| | change in SBP from childhood to early adulthood | | | | 0.475 |
| $PM_{10}$ in the 2$^{nd}$ trimester of pregnancy | mean difference in SBP at age 3y (mmHg) | 5,753 | 36,967 | -0.71 (-1.21, -0.21) | 0.006 |
| | change in SBP in childhood (mmHg/y): 3 to 10y | 5,753 | 36,967 | 0.11 (0.03, 0.19) | 0.010 |
| | change in SBP in adolescence (mmHg/y): 10 to 18y | 5,753 | 36,967 | 0.02 (-0.04, 0.08) | 0.521 |
| | change in SBP in adulthood (mmHg/y): 18 to 26y | 5,753 | 36,967 | -0.03 (-0.11, 0.06) | 0.516 |
| | change in SBP from childhood to early adulthood | | | | 0.020 |
| $PM_{10}$ in the 3$^{rd}$ trimester of pregnancy | mean difference in SBP at age 3y (mmHg) | 5,750 | 36,950 | -1.00 (-1.56, -0.43) | 5.11E-04 |
| | change in SBP in childhood (mmHg/y): 3 to 10y | 5,750 | 36,950 | 0.10 (0.01, 0.19) | 0.032 |
| | change in SBP in adolescence (mmHg/y): 10 to 18y | 5,750 | 36,950 | 0.07 (0.01, 0.12) | 0.024 |
| | change in SBP in adulthood (mmHg/y): 18 to 26y | 5,750 | 36,950 | 0.02 (-0.06, 0.09) | 0.674 |
| | change in SBP from childhood to early adulthood | | | | 4.05E-04 |

Adjusted for maternal education, age at delivery, ethnicity, area deprivation, and sex. Betas correspond to mean change in blood pressure by an interquartile range (IQR) increase in the exposure (for continuous exposures), or by change in category of exposure (for categorical exposures); the IQR and categories for each exposure are presented in Supplementary Table 4.

**Supplementary Table 10.** Associations between urban environmental exposures and changes in diastolic blood pressure (DBP) in ALSPAC, restricted to those with 3 or more blood pressure measurements

| Exposure | Parameter | N | N obs | beta (95% CI) | p-value |
|---|---|---|---|---|---|
| Lden | mean difference in DBP at age 3y (mmHg) | 4,199 | 26,875 | -0.15 (-0.57, 0.27) | 0.487 |
| | change in DBP in childhood (mmHg/y): 3 to 10y | 4,199 | 26,875 | 0.05 (-0.02, 0.12) | 0.185 |
| | change in DBP in adolescence (mmHg/y): 10 to 18y | 4,199 | 26,875 | -0.03 (-0.07, 0.02) | 0.217 |
| | change in DBP in adulthood (mmHg/y): 18 to 26y | 4,199 | 26,875 | 0.01 (-0.06, 0.08) | 0.759 |
| | change in DBP from childhood to early adulthood | | | | 0.483 |
| Lnight | mean difference in DBP at age 3y (mmHg) | 4,199 | 26,875 | -0.33 (-0.92, 0.26) | 0.272 |
| | change in DBP in childhood (mmHg/y): 3 to 10y | 4,199 | 26,875 | 0.08 (-0.02, 0.18) | 0.102 |
| | change in DBP in adolescence (mmHg/y): 10 to 18y | 4,199 | 26,875 | -0.04 (-0.11, 0.02) | 0.168 |
| | change in DBP in adulthood (mmHg/y): 18 to 26y | 4,199 | 26,875 | 0.04 (-0.06, 0.14) | 0.390 |
| | change in DBP from childhood to early adulthood | | | | 0.313 |
| Inverse distance to nearest road | mean difference in DBP at age 3y (mmHg) | 6,129 | 39,358 | -0.15 (-0.44, 0.14) | 0.307 |
| | change in DBP in childhood (mmHg/y): 3 to 10y | 6,129 | 39,358 | 0.02 (-0.03, 0.07) | 0.382 |
| | change in DBP in adolescence (mmHg/y): 10 to 18y | 6,129 | 39,358 | 0.00 (-0.03, 0.04) | 0.768 |
| | change in DBP in adulthood (mmHg/y): 18 to 26y | 6,129 | 39,358 | -0.04 (-0.09, 0.01) | 0.102 |
| | change in DBP from childhood to early adulthood | | | | 0.296 |
| Humidity | mean difference in DBP at age 3y (mmHg) | 6,087 | 39,083 | -1.54 (-2.00, -1.08) | 4.64E-11 |
| | change in DBP in childhood (mmHg/y): 3 to 10y | 6,087 | 39,083 | 0.33 (0.26, 0.41) | 3.36E-18 |
| | change in DBP in adolescence (mmHg/y): 10 to 18y | 6,087 | 39,083 | -0.16 (-0.20, -0.12) | 8.29E-13 |
| | change in DBP in adulthood (mmHg/y): 18 to 26y | 6,087 | 39,083 | 0.06 (-0.01, 0.13) | 0.076 |
| | change in DBP from childhood to early adulthood | | | | 1.59E-20 |
| Mean temperature | mean difference in DBP at age 3y (mmHg) | 6,096 | 39,143 | 0.20 (-0.29, 0.70) | 0.420 |
| | change in DBP in childhood (mmHg/y): 3 to 10y | 6,096 | 39,143 | -0.10 (-0.18, -0.02) | 0.020 |
| | change in DBP in adolescence (mmHg/y): 10 to 18y | 6,096 | 39,143 | 0.08 (0.03, 0.13) | 0.001 |
| | change in DBP in adulthood (mmHg/y): 18 to 26y | 6,096 | 39,143 | 0.03 (-0.05, 0.11) | 0.508 |
| | change in DBP from childhood to early adulthood | | | | 9.30E-04 |
| Building density in 100m buffer | mean difference in DBP at age 3y (mmHg) | 6,123 | 39,318 | 0.16 (-0.18, 0.50) | 0.353 |
| | change in DBP in childhood (mmHg/y): 3 to 10y | 6,123 | 39,318 | -0.03 (-0.08, 0.03) | 0.316 |

| | | | | | |
|---|---|---|---|---|---|
| | change in DBP in adolescence (mmHg/y): 10 to 18y | 6,123 | 39,318 | 0.01 (-0.03, 0.04) | 0.774 |
| | change in DBP in adulthood (mmHg/y): 18 to 26y | 6,123 | 39,318 | -0.03 (-0.08, 0.03) | 0.363 |
| | change in DBP from childhood to early adulthood | | | | 0.583 |
| Building density in 300m buffer | mean difference in DBP at age 3y (mmHg) | 6,126 | 39,337 | 0.12 (-0.22, 0.47) | 0.477 |
| | change in DBP in childhood (mmHg/y): 3 to 10y | 6,126 | 39,337 | -0.03 (-0.08, 0.03) | 0.366 |
| | change in DBP in adolescence (mmHg/y): 10 to 18y | 6,126 | 39,337 | 0.01 (-0.02, 0.05) | 0.509 |
| | change in DBP in adulthood (mmHg/y): 18 to 26y | 6,126 | 39,337 | -0.03 (-0.08, 0.03) | 0.351 |
| | change in DBP from childhood to early adulthood | | | | 0.666 |
| Connectivity in 100m buffer | mean difference in DBP at age 3y (mmHg) | 5,801 | 37,157 | -0.18 (-0.47, 0.12) | 0.236 |
| | change in DBP in childhood (mmHg/y): 3 to 10y | 5,801 | 37,157 | 0.01 (-0.04, 0.06) | 0.658 |
| | change in DBP in adolescence (mmHg/y): 10 to 18y | 5,801 | 37,157 | -0.02 (-0.05, 0.01) | 0.246 |
| | change in DBP in adulthood (mmHg/y): 18 to 26y | 5,801 | 37,157 | -0.02 (-0.07, 0.03) | 0.418 |
| | change in DBP from childhood to early adulthood | | | | 0.326 |
| Connectivity in 300m buffer | mean difference in DBP at age 3y (mmHg) | 6,108 | 39,217 | 0.05 (-0.28, 0.39) | 0.748 |
| | change in DBP in childhood (mmHg/y): 3 to 10y | 6,108 | 39,217 | -0.01 (-0.06, 0.04) | 0.708 |
| | change in DBP in adolescence (mmHg/y): 10 to 18y | 6,108 | 39,217 | 0.00 (-0.03, 0.03) | 0.986 |
| | change in DBP in adulthood (mmHg/y): 18 to 26y | 6,108 | 39,217 | -0.02 (-0.07, 0.03) | 0.484 |
| | change in DBP from childhood to early adulthood | | | | 0.847 |
| Facility density within 300m buffer | mean difference in DBP at age 3y (mmHg) | 6,129 | 39,358 | 0.12 (-0.08, 0.32) | 0.230 |
| | change in DBP in childhood (mmHg/y): 3 to 10y | 6,129 | 39,358 | -0.03 (-0.06, 0.00) | 0.086 |
| | change in DBP in adolescence (mmHg/y): 10 to 18y | 6,129 | 39,358 | 0.01 (-0.01, 0.03) | 0.358 |
| | change in DBP in adulthood (mmHg/y): 18 to 26y | 6,129 | 39,358 | -0.02 (-0.05, 0.01) | 0.267 |
| | change in DBP from childhood to early adulthood | | | | 0.268 |
| Land use within 300m buffer | mean difference in DBP at age 3y (mmHg) | 5,949 | 38,157 | 0.02 (-0.36, 0.40) | 0.927 |
| | change in DBP in childhood (mmHg/y): 3 to 10y | 5,949 | 38,157 | 0.00 (-0.06, 0.06) | 0.972 |
| | change in DBP in adolescence (mmHg/y): 10 to 18y | 5,949 | 38,157 | 0.02 (-0.02, 0.06) | 0.390 |
| | change in DBP in adulthood (mmHg/y): 18 to 26y | 5,949 | 38,157 | -0.02 (-0.08, 0.04) | 0.537 |
| | change in DBP from childhood to early adulthood | | | | 0.829 |
| Agricultural area within 300m buffer | mean difference in DBP at age 3y (mmHg) | 5,949 | 38,157 | 0.00 (-0.09, 0.08) | 0.932 |
| | change in DBP in childhood (mmHg/y): 3 to 10y | 5,949 | 38,157 | -0.01 (-0.02, 0.01) | 0.473 |
| | change in DBP in adolescence (mmHg/y): 10 to 18y | 5,949 | 38,157 | 0.00 (-0.01, 0.01) | 0.577 |

| | | | | | |
|---|---|---|---|---|---|
| | change in DBP in adulthood (mmHg/y): 18 to 26y | 5,949 | 38,157 | 0.01 (-0.01, 0.02) | 0.332 |
| | change in DBP from childhood to early adulthood | | | | 0.614 |
| Airport within 300m buffer | mean difference in DBP at age 3y (mmHg) | 5,949 | 38,157 | -0.73 (-4.31, 2.85) | 0.690 |
| | change in DBP in childhood (mmHg/y): 3 to 10y | 5,949 | 38,157 | 0.33 (-0.26, 0.91) | 0.278 |
| | change in DBP in adolescence (mmHg/y): 10 to 18y | 5,949 | 38,157 | -0.05 (-0.42, 0.32) | 0.782 |
| | change in DBP in adulthood (mmHg/y): 18 to 26y | 5,949 | 38,157 | 0.45 (-0.07, 0.96) | 0.089 |
| | change in DBP from childhood to early adulthood | | | | 0.187 |
| Continuous Urban Fabric within 300m buffer | mean difference in DBP at age 3y (mmHg) | 5,949 | 38,157 | 0.01 (-0.30, 0.31) | 0.962 |
| | change in DBP in childhood (mmHg/y): 3 to 10y | 5,949 | 38,157 | -0.01 (-0.06, 0.04) | 0.612 |
| | change in DBP in adolescence (mmHg/y): 10 to 18y | 5,949 | 38,157 | 0.02 (-0.01, 0.05) | 0.193 |
| | change in DBP in adulthood (mmHg/y): 18 to 26y | 5,949 | 38,157 | -0.06 (-0.11, 0.00) | 0.038 |
| | change in DBP from childhood to early adulthood | | | | 0.207 |
| Industrial, commercial, public, military and private units within 300m buffer | mean difference in DBP at age 3y (mmHg) | 5,949 | 38,157 | -0.19 (-0.48, 0.10) | 0.192 |
| | change in DBP in childhood (mmHg/y): 3 to 10y | 5,949 | 38,157 | 0.04 (-0.01, 0.09) | 0.101 |
| | change in DBP in adolescence (mmHg/y): 10 to 18y | 5,949 | 38,157 | -0.02 (-0.06, 0.01) | 0.114 |
| | change in DBP in adulthood (mmHg/y): 18 to 26y | 5,949 | 38,157 | 0.01 (-0.04, 0.06) | 0.615 |
| | change in DBP from childhood to early adulthood | | | | 0.279 |
| Discontinuous Low Density Urban Fabric within 300m buffer | mean difference in DBP at age 3y (mmHg) | 5,949 | 38,157 | 0.10 (-0.29, 0.49) | 0.605 |
| | change in DBP in childhood (mmHg/y): 3 to 10y | 5,949 | 38,157 | -0.01 (-0.08, 0.05) | 0.701 |
| | change in DBP in adolescence (mmHg/y): 10 to 18y | 5,949 | 38,157 | 0.02 (-0.02, 0.06) | 0.438 |
| | change in DBP in adulthood (mmHg/y): 18 to 26y | 5,949 | 38,157 | 0.00 (-0.07, 0.06) | 0.934 |
| | change in DBP from childhood to early adulthood | | | | 0.875 |
| Forests within 300m buffer | mean difference in DBP at age 3y (mmHg) | 5,949 | 38,157 | 0.09 (-0.36, 0.55) | 0.685 |
| | change in DBP in childhood (mmHg/y): 3 to 10y | 5,949 | 38,157 | -0.03 (-0.10, 0.05) | 0.492 |
| | change in DBP in adolescence (mmHg/y): 10 to 18y | 5,949 | 38,157 | 0.00 (-0.05, 0.05) | 0.897 |
| | change in DBP in adulthood (mmHg/y): 18 to 26y | 5,949 | 38,157 | 0.01 (-0.07, 0.09) | 0.804 |
| | change in DBP from childhood to early adulthood | | | | 0.904 |
| Mineral extraction and dump sites, constructions within 300m buffer | mean difference in DBP at age 3y (mmHg) | 5,949 | 38,157 | -0.19 (-0.69, 0.31) | 0.460 |
| | change in DBP in childhood (mmHg/y): 3 to 10y | 5,949 | 38,157 | 0.06 (-0.03, 0.14) | 0.188 |
| | change in DBP in adolescence (mmHg/y): 10 to 18y | 5,949 | 38,157 | -0.07 (-0.12, -0.01) | 0.013 |
| | change in DBP in adulthood (mmHg/y): 18 to 26y | 5,949 | 38,157 | 0.06 (-0.02, 0.15) | 0.154 |

| | | | | | |
|---|---|---|---|---|---|
| | change in DBP from childhood to early adulthood | | | | 0.087 |
| Port areas within 300m buffer | mean difference in DBP at age 3y (mmHg) | 5,949 | 38,157 | 2.17 (-4.06, 8.40) | 0.495 |
| | change in DBP in childhood (mmHg/y): 3 to 10y | 5,949 | 38,157 | -0.15 (-1.16, 0.87) | 0.775 |
| | change in DBP in adolescence (mmHg/y): 10 to 18y | 5,949 | 38,157 | -0.47 (-1.04, 0.11) | 0.114 |
| | change in DBP in adulthood (mmHg/y): 18 to 26y | 5,949 | 38,157 | 0.01 (-1.16, 1.17) | 0.993 |
| | change in DBP from childhood to early adulthood | | | | 0.262 |
| Transport networks and other constructed hard-surfaced areas within 300m buffer | mean difference in DBP at age 3y (mmHg) | 5,949 | 38,157 | 0.06 (-0.27, 0.40) | 0.717 |
| | change in DBP in childhood (mmHg/y): 3 to 10y | 5,949 | 38,157 | -0.01 (-0.06, 0.05) | 0.778 |
| | change in DBP in adolescence (mmHg/y): 10 to 18y | 5,949 | 38,157 | 0.00 (-0.03, 0.04) | 0.894 |
| | change in DBP in adulthood (mmHg/y): 18 to 26y | 5,949 | 38,157 | -0.02 (-0.07, 0.04) | 0.580 |
| | change in DBP from childhood to early adulthood | | | | 0.938 |
| Green urban areas within 300m buffer | mean difference in DBP at age 3y (mmHg) | 5,949 | 38,157 | -0.03 (-0.36, 0.30) | 0.848 |
| | change in DBP in childhood (mmHg/y): 3 to 10y | 5,949 | 38,157 | 0.03 (-0.02, 0.08) | 0.280 |
| | change in DBP in adolescence (mmHg/y): 10 to 18y | 5,949 | 38,157 | 0.02 (-0.01, 0.06) | 0.246 |
| | change in DBP in adulthood (mmHg/y): 18 to 26y | 5,949 | 38,157 | -0.04 (-0.09, 0.02) | 0.217 |
| | change in DBP from childhood to early adulthood | | | | 0.226 |
| Discontinuous Very Low Density Urban Fabric within 300m buffer | mean difference in DBP at age 3y (mmHg) | 5,949 | 38,157 | -0.64 (-1.87, 0.59) | 0.309 |
| | change in DBP in childhood (mmHg/y): 3 to 10y | 5,949 | 38,157 | 0.06 (-0.14, 0.26) | 0.549 |
| | change in DBP in adolescence (mmHg/y): 10 to 18y | 5,949 | 38,157 | 0.04 (-0.08, 0.16) | 0.518 |
| | change in DBP in adulthood (mmHg/y): 18 to 26y | 5,949 | 38,157 | -0.17 (-0.36, 0.01) | 0.067 |
| | change in DBP from childhood to early adulthood | | | | 0.268 |
| Population density | mean difference in DBP at age 3y (mmHg) | 6,002 | 38,517 | 0.29 (-0.08, 0.65) | 0.120 |
| | change in DBP in childhood (mmHg/y): 3 to 10y | 6,002 | 38,517 | -0.07 (-0.13, -0.01) | 0.023 |
| | change in DBP in adolescence (mmHg/y): 10 to 18y | 6,002 | 38,517 | 0.01 (-0.03, 0.05) | 0.608 |
| | change in DBP in adulthood (mmHg/y): 18 to 26y | 6,002 | 38,517 | 0.02 (-0.03, 0.08) | 0.408 |
| | change in DBP from childhood to early adulthood | | | | 0.103 |
| Bus lines in 300m buffer | mean difference in DBP at age 3y (mmHg) | 5,590 | 35,802 | 0.03 (-0.34, 0.39) | 0.893 |
| | change in DBP in childhood (mmHg/y): 3 to 10y | 5,590 | 35,802 | 0.02 (-0.04, 0.08) | 0.461 |
| | change in DBP in adolescence (mmHg/y): 10 to 18y | 5,590 | 35,802 | -0.03 (-0.07, 0.01) | 0.093 |
| | change in DBP in adulthood (mmHg/y): 18 to 26y | 5,590 | 35,802 | 0.03 (-0.03, 0.09) | 0.353 |
| | change in DBP from childhood to early adulthood | | | | 0.406 |

| | | | | | |
|---|---|---|---|---|---|
| Bus lines in 500m buffer | mean difference in DBP at age 3y (mmHg) | 5,936 | 38,095 | 0.09 (-0.29, 0.47) | 0.639 |
| | change in DBP in childhood (mmHg/y): 3 to 10y | 5,936 | 38,095 | -0.01 (-0.07, 0.06) | 0.850 |
| | change in DBP in adolescence (mmHg/y): 10 to 18y | 5,936 | 38,095 | -0.03 (-0.07, 0.01) | 0.172 |
| | change in DBP in adulthood (mmHg/y): 18 to 26y | 5,936 | 38,095 | 0.02 (-0.04, 0.09) | 0.462 |
| | change in DBP from childhood to early adulthood | | | | 0.495 |
| Walkability index | mean difference in DBP at age 3y (mmHg) | 5,928 | 38,016 | 0.12 (-0.21, 0.45) | 0.479 |
| | change in DBP in childhood (mmHg/y): 3 to 10y | 5,928 | 38,016 | -0.03 (-0.08, 0.03) | 0.313 |
| | change in DBP in adolescence (mmHg/y): 10 to 18y | 5,928 | 38,016 | 0.01 (-0.03, 0.04) | 0.714 |
| | change in DBP in adulthood (mmHg/y): 18 to 26y | 5,928 | 38,016 | -0.02 (-0.08, 0.03) | 0.398 |
| | change in DBP from childhood to early adulthood | | | | 0.627 |
| Distance to closest blue space | mean difference in DBP at age 3y (mmHg) | 5,946 | 38,134 | 0.15 (-0.20, 0.50) | 0.395 |
| | change in DBP in childhood (mmHg/y): 3 to 10y | 5,946 | 38,134 | -0.02 (-0.08, 0.03) | 0.405 |
| | change in DBP in adolescence (mmHg/y): 10 to 18y | 5,946 | 38,134 | -0.02 (-0.05, 0.02) | 0.400 |
| | change in DBP in adulthood (mmHg/y): 18 to 26y | 5,946 | 38,134 | 0.05 (-0.01, 0.11) | 0.106 |
| | change in DBP from childhood to early adulthood | | | | 0.279 |
| Size of blue space | mean difference in DBP at age 3y (mmHg) | 5,944 | 38,120 | -0.04 (-0.24, 0.16) | 0.715 |
| | change in DBP in childhood (mmHg/y): 3 to 10y | 5,944 | 38,120 | 0.01 (-0.02, 0.04) | 0.576 |
| | change in DBP in adolescence (mmHg/y): 10 to 18y | 5,944 | 38,120 | 0.00 (-0.02, 0.02) | 0.875 |
| | change in DBP in adulthood (mmHg/y): 18 to 26y | 5,944 | 38,120 | -0.01 (-0.04, 0.02) | 0.565 |
| | change in DBP from childhood to early adulthood | | | | 0.859 |
| Existence blue space within 300m buffer | mean difference in DBP at age 3y (mmHg) | 5,946 | 38,134 | 0.29 (-0.85, 1.43) | 0.619 |
| | change in DBP in childhood (mmHg/y): 3 to 10y | 5,946 | 38,134 | -0.05 (-0.23, 0.14) | 0.631 |
| | change in DBP in adolescence (mmHg/y): 10 to 18y | 5,946 | 38,134 | 0.01 (-0.10, 0.13) | 0.810 |
| | change in DBP in adulthood (mmHg/y): 18 to 26y | 5,946 | 38,134 | -0.10 (-0.28, 0.08) | 0.258 |
| | change in DBP from childhood to early adulthood | | | | 0.649 |
| Distance to closest green space | mean difference in DBP at age 3y (mmHg) | 5,946 | 38,134 | 0.08 (-0.30, 0.46) | 0.686 |
| | change in DBP in childhood (mmHg/y): 3 to 10y | 5,946 | 38,134 | 0.00 (-0.07, 0.06) | 0.888 |
| | change in DBP in adolescence (mmHg/y): 10 to 18y | 5,946 | 38,134 | -0.02 (-0.06, 0.02) | 0.354 |
| | change in DBP in adulthood (mmHg/y): 18 to 26y | 5,946 | 38,134 | 0.00 (-0.06, 0.07) | 0.891 |
| | change in DBP from childhood to early adulthood | | | | 0.744 |
| Size of green space | mean difference in DBP at age 3y (mmHg) | 5,946 | 38,134 | 0.08 (-0.29, 0.46) | 0.661 |

| | | | | | |
|---|---|---|---|---|---|
| | change in DBP in childhood (mmHg/y): 3 to 10y | 5,946 | 38,134 | 0.02 (-0.04, 0.08) | 0.531 |
| | change in DBP in adolescence (mmHg/y): 10 to 18y | 5,946 | 38,134 | -0.03 (-0.07, 0.01) | 0.174 |
| | change in DBP in adulthood (mmHg/y): 18 to 26y | 5,946 | 38,134 | 0.02 (-0.04, 0.08) | 0.522 |
| | change in DBP from childhood to early adulthood | | | | 0.598 |
| Existence green space within 300m buffer | mean difference in DBP at age 3y (mmHg) | 5,946 | 38,134 | -0.06 (-0.69, 0.57) | 0.850 |
| | change in DBP in childhood (mmHg/y): 3 to 10y | 5,946 | 38,134 | 0.00 (-0.10, 0.11) | 0.947 |
| | change in DBP in adolescence (mmHg/y): 10 to 18y | 5,946 | 38,134 | 0.03 (-0.03, 0.10) | 0.312 |
| | change in DBP in adulthood (mmHg/y): 18 to 26y | 5,946 | 38,134 | 0.02 (-0.09, 0.12) | 0.754 |
| | change in DBP from childhood to early adulthood | | | | 0.571 |
| Average greenness within 100m buffer | mean difference in DBP at age 3y (mmHg) | 6,129 | 39,358 | 0.71 (-2.15, 3.57) | 0.626 |
| | change in DBP in childhood (mmHg/y): 3 to 10y | 6,129 | 39,358 | -0.25 (-0.72, 0.21) | 0.286 |
| | change in DBP in adolescence (mmHg/y): 10 to 18y | 6,129 | 39,358 | 0.21 (-0.08, 0.51) | 0.157 |
| | change in DBP in adulthood (mmHg/y): 18 to 26y | 6,129 | 39,358 | 0.06 (-0.41, 0.52) | 0.808 |
| | change in DBP from childhood to early adulthood | | | | 0.363 |
| Average greenness within 300m buffer | mean difference in DBP at age 3y (mmHg) | 6,129 | 39,358 | 0.31 (-2.71, 3.32) | 0.842 |
| | change in DBP in childhood (mmHg/y): 3 to 10y | 6,129 | 39,358 | -0.10 (-0.59, 0.39) | 0.678 |
| | change in DBP in adolescence (mmHg/y): 10 to 18y | 6,129 | 39,358 | 0.16 (-0.14, 0.47) | 0.295 |
| | change in DBP in adulthood (mmHg/y): 18 to 26y | 6,129 | 39,358 | 0.08 (-0.41, 0.56) | 0.762 |
| | change in DBP from childhood to early adulthood | | | | 0.617 |
| Average greenness within 500m buffer | mean difference in DBP at age 3y (mmHg) | 6,129 | 39,358 | -0.83 (-3.90, 2.24) | 0.597 |
| | change in DBP in childhood (mmHg/y): 3 to 10y | 6,129 | 39,358 | 0.12 (-0.38, 0.61) | 0.647 |
| | change in DBP in adolescence (mmHg/y): 10 to 18y | 6,129 | 39,358 | -0.02 (-0.33, 0.29) | 0.904 |
| | change in DBP in adulthood (mmHg/y): 18 to 26y | 6,129 | 39,358 | 0.22 (-0.27, 0.72) | 0.381 |
| | change in DBP from childhood to early adulthood | | | | 0.770 |
| Food facilities density within 300m buffer | mean difference in DBP at age 3y (mmHg) | 6,129 | 39,358 | 0.18 (-0.01, 0.37) | 0.063 |
| | change in DBP in childhood (mmHg/y): 3 to 10y | 6,129 | 39,358 | -0.04 (-0.07, -0.01) | 0.011 |
| | change in DBP in adolescence (mmHg/y): 10 to 18y | 6,129 | 39,358 | 0.01 (-0.01, 0.03) | 0.434 |
| | change in DBP in adulthood (mmHg/y): 18 to 26y | 6,129 | 39,358 | 0.00 (-0.03, 0.03) | 0.845 |
| | change in DBP from childhood to early adulthood | | | | 0.093 |
| $NO_2$ | mean difference in DBP at age 3y (mmHg) | 6,095 | 39,139 | -0.10 (-0.45, 0.24) | 0.558 |
| | change in DBP in childhood (mmHg/y): 3 to 10y | 6,095 | 39,139 | 0.03 (-0.03, 0.08) | 0.374 |

| | | | | | |
|---|---|---|---|---|---|
| | change in DBP in adolescence (mmHg/y): 10 to 18y | 6,095 | 39,139 | 0.00 (-0.04, 0.03) | 0.896 |
| | change in DBP in adulthood (mmHg/y): 18 to 26y | 6,095 | 39,139 | -0.04 (-0.10, 0.01) | 0.110 |
| | change in DBP from childhood to early adulthood | | | | 0.268 |
| $PM_{2.5}$ | mean difference in DBP at age 3y (mmHg) | 6,095 | 39,139 | -0.38 (-0.73, -0.03) | 0.034 |
| | change in DBP in childhood (mmHg/y): 3 to 10y | 6,095 | 39,139 | 0.10 (0.04, 0.16) | 4.77E-04 |
| | change in DBP in adolescence (mmHg/y): 10 to 18y | 6,095 | 39,139 | -0.06 (-0.10, -0.03) | 7.03E-04 |
| | change in DBP in adulthood (mmHg/y): 18 to 26y | 6,095 | 39,139 | 0.01 (-0.05, 0.06) | 0.797 |
| | change in DBP from childhood to early adulthood | | | | 2.97E-04 |
| $PM_{10}$ | mean difference in DBP at age 3y (mmHg) | 5,777 | 37,100 | -0.31 (-0.64, 0.03) | 0.075 |
| | change in DBP in childhood (mmHg/y): 3 to 10y | 5,777 | 37,100 | 0.04 (-0.02, 0.09) | 0.162 |
| | change in DBP in adolescence (mmHg/y): 10 to 18y | 5,777 | 37,100 | 0.00 (-0.03, 0.04) | 0.847 |
| | change in DBP in adulthood (mmHg/y): 18 to 26y | 5,777 | 37,100 | -0.06 (-0.12, 0.00) | 0.048 |
| | change in DBP from childhood to early adulthood | | | | 0.095 |
| $PM_{10}$ in the 1$^{st}$ trimester of pregnancy | mean difference in DBP at age 3y (mmHg) | 5,640 | 36,244 | 0.64 (0.28, 1.01) | 5.63E-04 |
| | change in DBP in childhood (mmHg/y): 3 to 10y | 5,640 | 36,244 | -0.09 (-0.15, -0.03) | 0.002 |
| | change in DBP in adolescence (mmHg/y): 10 to 18y | 5,640 | 36,244 | -0.03 (-0.07, 0.02) | 0.248 |
| | change in DBP in adulthood (mmHg/y): 18 to 26y | 5,640 | 36,244 | -0.01 (-0.09, 0.06) | 0.702 |
| | change in DBP from childhood to early adulthood | | | | 6.89E-04 |
| $PM_{10}$ in the 2$^{nd}$ trimester of pregnancy | mean difference in DBP at age 3y (mmHg) | 5,753 | 36,968 | -0.73 (-1.12, -0.34) | 2.61E-04 |
| | change in DBP in childhood (mmHg/y): 3 to 10y | 5,753 | 36,968 | 0.17 (0.11, 0.24) | 1.59E-07 |
| | change in DBP in adolescence (mmHg/y): 10 to 18y | 5,753 | 36,968 | -0.07 (-0.11, -0.03) | 9.98E-04 |
| | change in DBP in adulthood (mmHg/y): 18 to 26y | 5,753 | 36,968 | -0.05 (-0.11, 0.02) | 0.178 |
| | change in DBP from childhood to early adulthood | | | | 4.52E-08 |
| $PM_{10}$ in the 3$^{rd}$ trimester of pregnancy | mean difference in DBP at age 3y (mmHg) | 5,750 | 36,951 | -0.93 (-1.38, -0.48) | 5.29E-05 |
| | change in DBP in childhood (mmHg/y): 3 to 10y | 5,750 | 36,951 | 0.08 (0.00, 0.15) | 0.039 |
| | change in DBP in adolescence (mmHg/y): 10 to 18y | 5,750 | 36,951 | 0.08 (0.04, 0.12) | 8.87-E05 |
| | change in DBP in adulthood (mmHg/y): 18 to 26y | 5,750 | 36,951 | -0.04 (-0.10, 0.02) | 0.210 |
| | change in DBP from childhood to early adulthood | | | | 9.42E-07 |

Adjusted for maternal education, age at delivery, ethnicity, area deprivation, and sex. Betas correspond to mean change in blood pressure by an interquartile range (IQR) increase in the exposure (for continuous exposures), or by change in category of exposure (for categorical exposures); the IQR and categories for each exposure are presented in Supplementary Table 4.

**Supplementary Table 11.** Associations of urban environmental exposures with systolic (SBP) and diastolic blood pressure (DBP) in males and females in ALSPAC.

| Exposure | Parameter | Males beta (95% CI) | p-value | Females beta (95% CI) | p-value | p-value for sex interaction |
|---|---|---|---|---|---|---|
| **Systolic blood pressure** | | | | | | |
| Humidity | mean difference in SBP at age 3y (mmHg) | -2.06 (-2.83, -1.30) | 1.19E-07 | -1.95 (-2.77, -1.13) | 3.24E-06 | 0.852 |
| | change in SBP in childhood (mmHg/y) | 0.32 (0.19, 0.45) | 1.11E-06 | 0.30 (0.17, 0.44) | 1.33E-05 | 0.872 |
| | change in SBP in adolescence (mmHg/y) | 0.02 (-0.06, 0.11) | 0.572 | 0.01 (-0.07, 0.09) | 0.847 | 0.755 |
| | change in SBP in adulthood (mmHg/y) | -0.18 (-0.31, -0.04) | 0.009 | -0.08 (-0.18, 0.02) | 0.127 | 0.219 |
| | change from childhood to early adulthood | | 1.16E-07 | | 2.20E-05 | 0.658 |
| Mean temperature | mean difference in SBP at age 3y (mmHg) | 1.00 (0.17, 1.82) | 0.018 | 0.84 (-0.07, 1.74) | 0.069 | 0.803 |
| | change in SBP in childhood (mmHg/y) | -0.25 (-0.39, -0.11) | 4.29E-04 | -0.13 (-0.28, 0.02) | 0.099 | 0.249 |
| | change in SBP in adolescence (mmHg/y) | 0.10 (0.00, 0.20) | 0.042 | -0.03 (-0.12, 0.07) | 0.587 | 0.091 |
| | change in SBP in adulthood (mmHg/y) | 0.01 (-0.15, 0.16) | 0.933 | 0.08 (-0.03, 0.20) | 0.158 | 0.508 |
| | change from childhood to early adulthood | | 0.004 | | 0.137 | 0.366 |
| $PM_{10}$ in the first trimester of pregnancy | mean difference in SBP at age 3y (mmHg) | -0.04 (-0.67, 0.60) | 0.912 | 0.46 (-0.18, 1.10) | 0.160 | 0.246 |
| | change in SBP in childhood (mmHg/y) | 0.02 (-0.09, 0.13) | 0.719 | -0.15 (-0.25, -0.04) | 0.008 | 0.031 |
| | change in SBP in adolescence (mmHg/y) | -0.02 (-0.10, 0.06) | 0.645 | 0.05 (-0.03, 0.12) | 0.254 | 0.299 |
| | change in SBP in adulthood (mmHg/y) | 0.03 (-0.11, 0.17) | 0.705 | -0.05 (-0.15, 0.05) | 0.344 | 0.383 |
| | change from childhood to early adulthood | | 0.958 | | 0.052 | 0.162 |
| **Diastolic blood pressure** | | | | | | |
| Humidity | mean difference in DBP at age 3y (mmHg) | -1.58 (-2.18, -0.97) | 3.80E-07 | -1.58 (-2.23, -0.92) | 2.35E-06 | 0.974 |
| | change in DBP in childhood (mmHg/y) | 0.33 (0.23, 0.43) | 9.51E-11 | 0.35 (0.24, 0.45) | 3.46E-10 | 0.892 |
| | change in DBP in adolescence (mmHg/y) | -0.13 (-0.20, -0.07) | 5.04E-05 | -0.18 (-0.24, -0.13) | 2.75E-10 | 0.256 |
| | change in DBP in adulthood (mmHg/y) | 0.03 (-0.09, 0.14) | 0.664 | 0.07 (-0.02, 0.15) | 0.114 | 0.535 |
| | change from childhood to early adulthood | | 4.15E-10 | | 1.27E-12 | 0.708 |
| Mean temperature | mean difference in DBP at age 3y (mmHg) | 0.16 (-0.49, 0.82) | 0.628 | 0.17 (-0.55, 0.89) | 0.648 | 0.897 |
| | change in DBP in childhood (mmHg/y) | -0.12 (-0.22, -0.01) | 0.037 | -0.06 (-0.18, 0.06) | 0.305 | 0.491 |
| | change in DBP in adolescence (mmHg/y) | 0.11 (0.04, 0.19) | 0.004 | 0.05 (-0.02, 0.12) | 0.147 | 0.384 |

| | | | | | | |
|---|---|---|---|---|---|---|
| | change in DBP in adulthood (mmHg/y) | 0.03 (-0.10, 0.16) | 0.702 | 0.05 (-0.05, 0.14) | 0.343 | 0.879 |
| | change from childhood to early adulthood | | 0.005 | | 0.152 | 0.810 |
| $PM_{2.5}$ | mean difference in DBP at age 3y (mmHg) | -0.30 (-0.75, 0.16) | 0.197 | -0.56 (-1.07, -0.05) | 0.031 | 0.566 |
| | change in DBP in childhood (mmHg/y) | 0.09 (0.02, 0.17) | 0.015 | 0.14 (0.05, 0.22) | 0.001 | 0.529 |
| | change in DBP in adolescence (mmHg/y) | -0.06 (-0.11, 0.00) | 0.041 | -0.07 (-0.12, -0.03) | 0.002 | 0.548 |
| | change in DBP in adulthood (mmHg/y) | -0.03 (-0.12, 0.07) | 0.588 | 0.02 (-0.05, 0.08) | 0.638 | 0.475 |
| | change from childhood to early adulthood | | 0.021 | | 0.002 | 0.836 |
| $PM_{10}$ in the first trimester of pregnancy | mean difference in DBP at age 3y (mmHg) | 0.52 (0.02, 1.02) | 0.042 | 0.69 (0.20, 1.19) | 0.006 | 0.578 |
| | change in DBP in childhood (mmHg/y) | -0.06 (-0.15, 0.02) | 0.142 | -0.14 (-0.22, -0.05) | 0.001 | 0.228 |
| | change in DBP in adolescence (mmHg/y) | -0.03 (-0.10, 0.04) | 0.386 | 0.00 (-0.06, 0.06) | 0.966 | 0.777 |
| | change in DBP in adulthood (mmHg/y) | 0.00 (-0.11, 0.12) | 0.946 | -0.04 (-0.13, 0.04) | 0.317 | 0.605 |
| | change from childhood to early adulthood | | 0.204 | | 0.002 | 0.615 |
| $PM_{10}$ in the second trimester of pregnancy | mean difference in DBP at age 3y (mmHg) | -0.56 (-1.07, -0.05) | 0.032 | -1.03 (-1.60, -0.47) | 3.36E-04 | 0.304 |
| | change in DBP in childhood (mmHg/y) | 0.16 (0.08, 0.25) | 1.68E-04 | 0.21 (0.11, 0.30) | 1.74E-05 | 0.593 |
| | change in DBP in adolescence (mmHg/y) | -0.06 (-0.13, 0.00) | 0.043 | -0.07 (-0.13, -0.02) | 0.011 | 0.809 |
| | change in DBP in adulthood (mmHg/y) | -0.09 (-0.20, 0.02) | 0.109 | -0.04 (-0.12, 0.04) | 0.377 | 0.482 |
| | change from childhood to early adulthood | | 1.10E-04 | | 4.36E-05 | 0.849 |
| $PM_{10}$ in the third trimester of pregnancy | mean difference in DBP at age 3y (mmHg) | -1.36 (-1.96, -0.75) | 1.11E-05 | -0.49 (-1.11, 0.14) | 0.128 | 0.058 |
| | change in DBP in childhood (mmHg/y) | 0.15 (0.05, 0.25) | 0.003 | 0.01 (-0.10, 0.11) | 0.888 | 0.048 |
| | change in DBP in adolescence (mmHg/y) | 0.10 (0.04, 0.16) | 5.94E-04 | 0.05 (0.00, 0.10) | 0.058 | 0.282 |
| | change in DBP in adulthood (mmHg/y) | -0.04 (-0.14, 0.06) | 0.409 | -0.04 (-0.11, 0.03) | 0.301 | 0.910 |
| | change from childhood to early adulthood | | 1.31E-07 | | 0.213 | 0.034 |

Adjusted for maternal education, age at delivery, ethnicity, and area deprivation. Betas correspond to mean change in blood pressure by an interquartile range (IQR) increase in the exposure (for continuous exposures), or by change in category of exposure (for categorical exposures); the IQR and categories for each exposure are presented in Supplementary Table 4.

**Supplementary Table 12.** Distribution of baseline characteristics, environmental exposures and outcomes in the participants included in the analysis in GenR, EDEN, PANIC and NFBC1986

| Variable | GenR (N= 6,331) | EDEN Nancy (N= 628) | EDEN Poitiers (N= 713) | PANIC (N= 369) | NFBC1986 (N= 1,220) |
|---|---|---|---|---|---|
| | N (%), mean (SD) or median (IQR) | | | | |
| Sex | | | | | |
| Males | 3,157 (49.9) | 309 (49.2) | 397 (55.7) | 186 (50.4) | 475 (38.0) |
| Females | 3,174 (50.1) | 319 (50.8) | 316 (44.3) | 183 (49.6) | 745 (61.1) |
| Maternal education | | | | | |
| Low | 582 (9.2) | 21 (3.3) | 42 (5.9) | 107 (29.0) | 457 (37.5) |
| Medium | 2,761 (43.6) | 190 (30.3) | 304 (42.6) | 166 (45.0) | 435 (35.6) |
| High | 2,988 (47.2) | 417 (66.4) | 367 (51.5) | 96 (26.0) | 328 (26.9) |
| Ethnicity | | | | | NA |
| White | - | - | - | 368 (99.7) | - |
| Non-White | - | - | - | 1 (0.3) | - |
| Western | 3,847 (60.8) | 537 (99.6) | 635 (99.4) | - | - |
| Non-Western | 1,860 (29.4) | - | 1 (0.2) | - | - |
| Mixed | 624 (9.9) | 2 (0.4) | 3 (0.5) | - | - |
| Area socioeconomic status | | | | NA | NA |
| 1st quintile (less deprived) | 671 (10.6) | 142 (22.6) | 230 (32.3) | - | - |
| 2nd quintile | 691 (10.9) | 115 (18.3) | 181 (25.4) | - | - |
| 3rd quintile | 743 (11.7) | 94 (15.0) | 120 (16.8) | - | - |
| 4th quintile | 694 (11.0) | 128 (20.4) | 116 (16.3) | - | - |
| 5th quintile (more deprived) | 3,532 (55.8) | 149 (23.7) | 66 (9.3) | - | - |
| Age of mother at birth (years), mean (SD) | 31.0 (5.0) | 30.2 (4.7) | 29.8 (4.8) | 30.0 (5.2) | 28.5 (5.5) |
| Humidity (%), median (IQR) | 81.2 (1.7) | 77.4 (3.7) | 76.9 (4.6) | 84.5 (5.6) | 77.5 (9.9) |
| Mean temperature (Celsius), median (IQR) | 10.8 (2.1) | 9.8 (3.2) | 12.1 (3.3) | 6.7 (5.6) | 1.4 (11.1) |
| $PM_{2.5}$ (μg/m$^3$), median (IQR) | 19.7 (3.9) | 17.8 (2.5) | 16.4 (2.3) | - | - |
| $PM_{10}$ (μg/m$^3$), median (IQR) | 31.4 (6.2) | 23.2 (3.1) | 15.7 (2.2) | 12.8 (8.4) | - |

| | | | | | |
|---|---|---|---|---|---|
| $PM_{10}$ in the 1st trimester of pregnancy (μg/m$^3$), median (IQR) | 31.7 (6.3) | - | - | - | - |
| $PM_{10}$ in the 2nd trimester of pregnancy (μg/m$^3$), median (IQR) | 31.5 (6.3) | - | - | - | - |
| $PM_{10}$ in the 3rd trimester of pregnancy (μg/m$^3$), median (IQR) | 31.3 (7.4) | - | - | - | - |
| Age at 1st assessment (years), mean (SD) | 2.1 (0.10) | 3.1 (0.08) | 3.1 (0.07) | 7.6 (0.39) | 16.0 (0.38) |
| Age at 2nd assessment (years), mean (SD) | 6.1 (0.42) | 5.6 (0.18) | 5.6 (0.12) | 9.8 (0.43) | 34.1 (0.59) |
| Age at 3rd assessment (years), mean (SD) | 9.8 (0.38) | - | - | 15.8 (0.44) | - |
| SBP at 1st assessment (mmHg), mean (SD) | 101.6 (11.4) | 97.8 (7.3) | 88.2 (6.3) | 100.3 (7.5) | 114.4 (12.4) |
| SBP at 2nd assessment (mmHg), mean (SD) | 103.1 (8.0) | 102.1 (7.4) | 101.9 (8.6) | 100.2 (7.8) | 112.0 (12.6) |
| SBP at 3rd assessment (mmHg), mean (SD) | 103.8 (7.7) | - | - | 113.2 (10.0) | - |
| DBP at 1st assessment (mmHg), mean (SD) | 62.6 (11.6) | 53.8 (7.1) | 45.7 (6.2) | 61.5 (7.04) | 67.3 (7.4) |
| DBP at 2nd assessment (mmHg), mean (SD) | 61.3 (6.7) | 59.2 (6.2) | 49.8 (7.4) | 61.4 (7.7) | 74.5 (8.8) |
| DBP at 3rd assessment (mmHg), mean (SD) | 59.4 (6.2) | - | - | 67.4 (9.52) | - |

DBP: diastolic blood pressure; IQR: interquartile range; NA: not applicable; $PM_{2.5}$: particulate matter <2.5 μm; $PM_{10}$: particulate matter <10 μm; SBP: systolic blood pressure; SD: standard deviation

**Supplementary Table 13.** Association between confounders and urban environmental exposures in each cohort

| Confounder | Humidity | | Temperature | | $PM_{2.5}$ | | $PM_{10}$ | |
|---|---|---|---|---|---|---|---|---|
| | β (95% CI) | p-value | β (95% CI) | p-value | β (95% CI) | p-value | β (95% CI) | p-value |
| **ALSPAC** | | | | | | | | |
| Maternal education (ref high) | | | | | | | | |
| Medium | 0.07 (-0.01, 0.16) | 0.089 | -0.03 (-0.11, 0.06) | 0.536 | 0.11 (0.06, 0.17) | <0.001 | -0.73 (-0.93, -0.53) | <0.001 |
| Low | 0.00 (-0.11, 0.11) | 0.987 | 0.05 (-0.06, 0.16) | 0.375 | 0.12 (0.05, 0.19) | 0.001 | -0.81 (-1.07, -0.55) | <0.001 |
| Ethnicity (Non-White) | 0.00 (-0.20, 0.21) | 0.966 | -0.13 (-0.34, 0.08) | 0.230 | 0.29 (0.15, 0.42) | <0.001 | 1.54 (1.05, 2.03) | <0.001 |
| Area socioeconomic status (ref 1st quintile) | | | | | | | | |
| 2nd quintile | -0.05 (-0.15, 0.04) | 0.284 | -0.01 (-0.11, 0.09) | 0.868 | -0.09 (-0.16, -0.02) | 0.010 | -0.05 (-0.26, 0.17) | 0.686 |
| 3rd quintile | -0.08 (-0.17, 0.01) | 0.086 | -0.01 (-0.11, 0.08) | 0.774 | 0.08 (0.02, 0.14) | 0.013 | 0.86 (0.66, 1.07) | <0.001 |
| 4th quintile | -0.04 (-0.13, 0.06) | 0.426 | -0.01 (-0.10, 0.09) | 0.875 | 0.22 (0.16, 0.28) | <0.001 | 1.46 (1.27, 1.66) | <0.001 |
| 5th quintile (more deprived) | -0.13 (-0.23, -0.04) | 0.006 | 0.04 (-0.05, 0.14) | 0.381 | 0.30 (0.24, 0.36) | <0.001 | 0.95 (0.73, 1.16) | <0.001 |
| Age of mother at birth (years) | 0.00 (0.00, 0.01) | 0.159 | 0.00 (-0.01, 0.01) | 0.636 | -0.01 (-0.02, -0.01) | <0.001 | 0.01 (-0.01, 0.02) | 0.246 |
| **GenR** | | | | | | | | |
| Maternal education (ref high) | | | | | | | | |
| Medium | -0.06 (-0.07, -0.04) | <0.001 | 0.01 (-0.00, 0.01) | 0.078 | 0.16 (0.14, 0.17) | <0.001 | 0.04 (0.01, 0.06) | 0.005 |
| Low | 0.06 (0.04, 0.08) | <0.001 | -0.01 (-0.03, -0.00) | 0.022 | -0.19 (-0.22, -0.17) | <0.001 | -0.61 (-0.66, -0.57) | <0.001 |
| Ethnicity (Western) | | | | | | | | |
| Non-Western | 0.12 (0.10, 0.14) | <0.001 | -0.08 (-0.09, -0.08) | <0.001 | -0.19 (-0.21, -0.17) | <0.001 | -0.51 (-0.54, -0.49) | <0.001 |
| Mixed | 0.09 (0.07, 0.10) | <0.001 | -0.08 (-0.09, -0.07) | <0.001 | -0.21 (-0.24, -0.19) | <0.001 | -0.33 (-0.37, -0.29) | <0.001 |
| Area socioeconomic status (ref 1st quintile) | | | | | | | | |
| 2nd quintile | -0.21 (-0.24, -0.19) | <0.001 | 0.18 (0.16, 0.19) | <0.001 | 0.62 (0.59, 0.65) | <0.001 | 1.26 (1.21, 1.31) | <0.001 |
| 3rd quintile | -0.08 (-0.11, -0.06) | <0.001 | 0.14 (0.12, 0.15) | <0.001 | -0.15 (-0.18, -0.12) | <0.001 | -0.63 (-0.68, -0.58) | <0.001 |
| 4th quintile | -0.13 (-0.15 -0.11) | <0.001 | 0.09 (0.08, 0.11) | <0.001 | 0.61 (0.58, 0.64) | <0.001 | 1.20 (1.15, 1.25) | <0.001 |
| 5th quintile (more deprived) | -0.12 (-0.14, -0.11) | <0.001 | 0.12 (0.11, 0.13) | <0.001 | 0.35 (0.33, 0.38) | <0.001 | 0.47 (0.43, 0.51) | <0.001 |
| Age of mother at birth (years) | 0.01 (0.00, 0.01) | <0.001 | 0.00 (0.00, 0.00) | <0.001 | -0.01 (-0.01, -0.01) | <0.001 | 0.00 (0.00, 0.00) | 0.108 |
| **EDEN (Nancy)** | | | | | | | | |
| Maternal education (ref high) | | | | | | | | |
| Medium | 0.26 (-0.03, 0.56) | 0.082 | -0.06 (-0.31, 0.20) | 0.666 | -0.15 (-0.39, 0.09) | 0.222 | -0.20 (-0.57, 0.18) | 0.311 |

| | | | | | | | | |
|---|---|---|---|---|---|---|---|---|
| Low | -0.43 (-1.21, 0.35) | 0.277 | 0.14 (-0.53, 0.81) | 0.677 | 0.42 (-0.21, 1.04) | 0.189 | 0.23 (-0.68, 1.14) | 0.626 |
| Area socioeconomic status (ref 1st quintile) | | | | | | | | |
| 2nd quintile | 0.23 (-0.19, 0.66) | 0.276 | -0.08 (-0.45, 0.28) | 0.648 | 0.02 (-0.32, 0.35) | 0.914 | 1.91 (1.43, 2.38) | <0.001 |
| 3rd quintile | 0.42 (-0.01, 0.86) | 0.058 | -0.22 (-0.60, 0.15) | 0.245 | 0.72 (0.37, 1.07) | <0.001 | 0.94 (0.49, 1.39) | <0.001 |
| 4th quintile | 0.01 (-0.39, 0.42) | 0.944 | 0.09 (-0.26, 0.44) | 0.632 | 0.55 (0.23, 0.88) | 0.001 | 2.49 (2.07, 2.92) | <0.001 |
| 5th quintile (more deprived) | 0.38 (-0.01, 0.78) | 0.057 | -0.15 (-0.49, 0.19) | 0.381 | 0.34 (0.03, 0.66) | 0.034 | 3.00 (2.58, 3.43) | <0.001 |
| Age of mother at birth (years) | 0.06 (0.03, 0.08) | <0.001 | -0.03 (-0.06, -0.01) | 0.010 | -0.02 (-0.05, 0.00) | 0.038 | -0.02 (-0.05, 0.02) | 0.313 |
| **EDEN (Poitiers)** | | | | | | | | |
| Maternal education (ref high) | | | | | | | | |
| Medium | -0.05 (-0.43, 0.32) | 0.783 | -0.10 (-0.32, 0.12) | 0.366 | 0.05 (-0.20, 0.31) | 0.692 | 0.10 (-0.12, 0.32) | 0.371 |
| Low | -0.04 (-0.88, 0.79) | 0.919 | -0.39 (-0.87, 0.10) | 0.118 | 0.74 (0.19, 1.30) | 0.009 | -0.20 (-0.72, 0.32) | 0.449 |
| Area socioeconomic status (ref 1st quintile) | | | | | | | | |
| 2nd quintile | -0.32 (-0.80, 0.16) | 0.186 | 0.37 (0.10, 0.65) | 0.008 | -0.10 (-0.42, 0.22) | 0.543 | 0.17 (-0.14, 0.47) | 0.281 |
| 3rd quintile | 0.37 (-0.18, 0.91) | 0.189 | -0.15 (-0.46, 0.17) | 0.363 | 0.25 (-0.11, 0.62) | 0.173 | 0.48 (0.13, 0.82) | 0.007 |
| 4th quintile | 0.32 (-0.25, 0.88) | 0.273 | -0.19 (-0.51, 0.14) | 0.261 | 0.80 (0.42, 1.17) | <0.001 | 0.95 (0.64, 1.25) | <0.001 |
| 5th quintile (more deprived) | 0.38 (-0.30, 1.07) | 0.270 | -0.07 (-0.46, 0.32) | 0.727 | 0.34 (-0.12, 0.80) | 0.142 | 0.29 (-0.03, 0.61) | 0.078 |
| Age of mother at birth (years) | 0.04 (0.00, 0.08) | 0.040 | -0.02 (-0.04, 0.00) | 0.058 | 0.01 (-0.02, 0.03) | 0.648 | -0.01 (-0.03, 0.01) | 0.244 |
| **PANIC** | | | | | | | | |
| Maternal education (ref high) | | | | | | | | |
| Medium | 0.99 (0.39, 1.59) | 0.001 | -0.26 (-0.60, 0.07) | 0.124 | - | - | 0.01 (-0.17, 0.19) | 0.916 |
| Low | 0.87 (0.19,1.55) | 0.011 | 0.099 (-0.28, 0.48) | 0.611 | - | - | -0.13 (-0.33, 0.08) | 0.229 |
| Ethnicity (Non-White) | -0.90 (-2.7, 0.90) | 0.328 | 0.98 (-0.03, 2.00) | 0.057 | - | - | 0.36 (-0.19, 0.91) | 0.195 |
| Age of mother at birth (years) | 0.07 (0.02, 0.12) | 0.003 | 0.00 (-0.02, 0.03) | 0.846 | - | - | 0.01 (-0.01, 0.02) | 0.410 |
| **NFBC1986** | | | | | | | | |
| Maternal education (ref high) | | | | | | | | |
| Medium | 1.14 (0.14, 2.14) | 0.026 | -0.98 (-2.11, 0.15) | 0.089 | - | - | - | - |
| Low | 0.23 (-0.75, 1.22) | 0.641 | -0.98 (-2.10, 0.13) | 0.084 | - | - | - | - |
| Age of mother at birth (years) | -0.06 (-0.13, 0.01) | 0.105 | -0.02 (-0.06, 0.10) | 0.610 | - | - | - | - |

**Supplementary Table 14.** Associations of humidity and temperature, mutually adjusted, with changes in systolic (SBP) and diastolic blood pressure (DBP) in ALSPAC

| Exposure | Parameter | N | N obs | beta (95% CI) | p-value |
|---|---|---|---|---|---|
| **Systolic blood pressure** | | | | | |
| Humidity | mean difference in SBP at age 3y (mmHg) | 7,402 | 40,925 | -2.16 (-2.74, -1.57) | 7.56E-13 |
| | change in SBP in childhood (mmHg/y): 3 to 10y | 7,402 | 40,925 | 0.30 (0.21, 0.39) | 3.95E-10 |
| | change in SBP in adolescence (mmHg/y): 10 to 18y | 7,402 | 40,925 | 0.06 (0.00, 0.12) | 0.062 |
| | change in SBP in adulthood (mmHg/y): 18 to 26y | 7,402 | 40,925 | -0.11 (-0.20, -0.03) | 0.010 |
| | change in SBP from childhood to early adulthood | | | | 1.28E-13 |
| Mean temperature | mean difference in SBP at age 3y (mmHg) | 7,413 | 40,988 | 0.74 (0.09, 1.38) | 0.024 |
| | change in SBP in childhood (mmHg/y): 3 to 10y | 7,413 | 40,988 | -0.17 (-0.28, -0.07) | 0.001 |
| | change in SBP in adolescence (mmHg/y): 10 to 18y | 7,413 | 40,988 | 0.02 (-0.06, 0.09) | 0.638 |
| | change in SBP in adulthood (mmHg/y): 18 to 26y | 7,413 | 40,988 | 0.02 (-0.08, 0.12) | 0.649 |
| | change in SBP from childhood to early adulthood | | | | 0.008 |
| **Diastolic blood pressure** | | | | | |
| Humidity | mean difference in DBP at age 3y (mmHg) | 7,402 | 40,925 | -1.70 (-2.17, -1.24) | 7.06E-13 |
| | change in DBP in childhood (mmHg/y): 3 to 10y | 7,402 | 40,925 | 0.34 (0.27, 0.42) | 9.20E-20 |
| | change in DBP in adolescence (mmHg/y): 10 to 18y | 7,402 | 40,925 | -0.16 (-0.20, -0.12) | 4.47E-13 |
| | change in DBP in adulthood (mmHg/y): 18 to 26y | 7,402 | 40,925 | 0.06 (-0.01, 0.13) | 0.114 |
| | change in DBP from childhood to early adulthood | | | | 5.82E-22 |
| Mean temperature | mean difference in DBP at age 3y (mmHg) | 7,413 | 40,988 | 0.19 (-0.31, 0.69) | 0.464 |
| | change in DBP in childhood (mmHg/y): 3 to 10y | 7,413 | 40,988 | -0.09 (-0.17, -0.01) | 0.032 |
| | change in DBP in adolescence (mmHg/y): 10 to 18y | 7,413 | 40,988 | 0.08 (0.03, 0.13) | 0.002 |
| | change in DBP in adulthood (mmHg/y): 18 to 26y | 7,413 | 40,988 | 0.04 (-0.04, 0.12) | 0.342 |
| | change in DBP from childhood to early adulthood | | | | 0.001 |

DBP: diastolic blood pressure; SBP: systolic blood pressure

**Supplementary Table 15.** Association between being a complete case and trajectory of systolic and diastolic blood pressure from childhood to early adulthood in ALSPAC (n=9,234)

| | **Mean (95%CI)** |
|---|---|
| **Systolic blood pressure (SBP)** | |
| SBP at 3y, mmHg | 94.6 (93.7, 95.5) |
| Mean difference in SBP at 3y in complete cases, mmHg | -1.73 (-2.69, -0.77) |
| Change in SBP in childhood (3 to 10y), mmHg/y | 1.31 (1.16, 1.47) |
| Change in SBP in childhood in complete cases, mmHg/y | 0.25 (0.08, 0.41) |
| Change in SBP in adolescence (10 to 18y), mmHg/y | 2.55 (2.46, 2.65) |
| Change in SBP in adolescence in complete cases, mmHg/y | 0.01 (-0.09, 0.12) |
| Change in SBP in early adulthood (18 to 24y), mmHg/y | -0.88 (-1.00, -0.76) |
| Change in SBP in adulthood in complete cases, mmHg/y | -0.14 (-0.28, -0.01) |
| **Diastolic blood pressure (DBP)** | |
| DBP at 3y, mmHg | 56.5 (55.8, 57.2) |
| Change in DBP in childhood (3 to 10y), mmHg/y | 0.10 (-0.01, 0.22) |
| Change in DBP in childhood in complete cases, mmHg/y | 0.06 (-0.06, 0.19) |
| Change in DBP in adolescence (10 to 18y), mmHg/y | 1.07 (1.00, 1.14) |
| Change in DBP in adolescence in complete cases, mmHg/y | -0.02, (-0.09, 0.05) |
| Change in DBP in early adulthood (18 to 24y), mmHg/y | 0.15 (0.05, 0.25) |
| Change in DBP in adulthood in complete cases, mmHg/y | -0.01 (-0.12, 0.09) |

Complete case corresponds to having complete data in all confounders used in the analyses i.e., maternal education, age at delivery, ethnicity, and area deprivation). Analysis was adjusted for sex.
DBP: diastolic blood pressure; SBP: systolic blood pressure

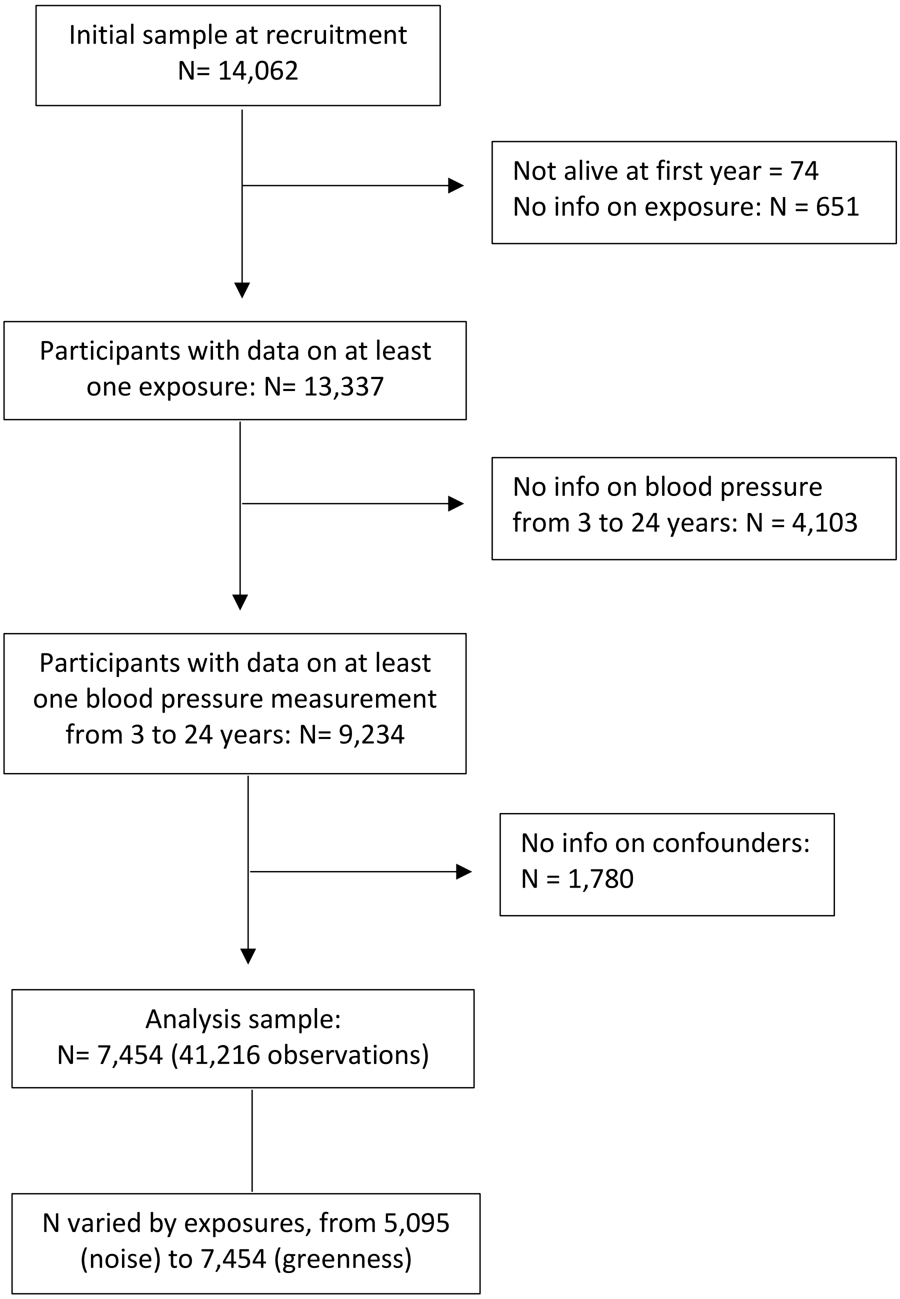

**Supplementary Figure 1.** Flow of participants in ALSPAC

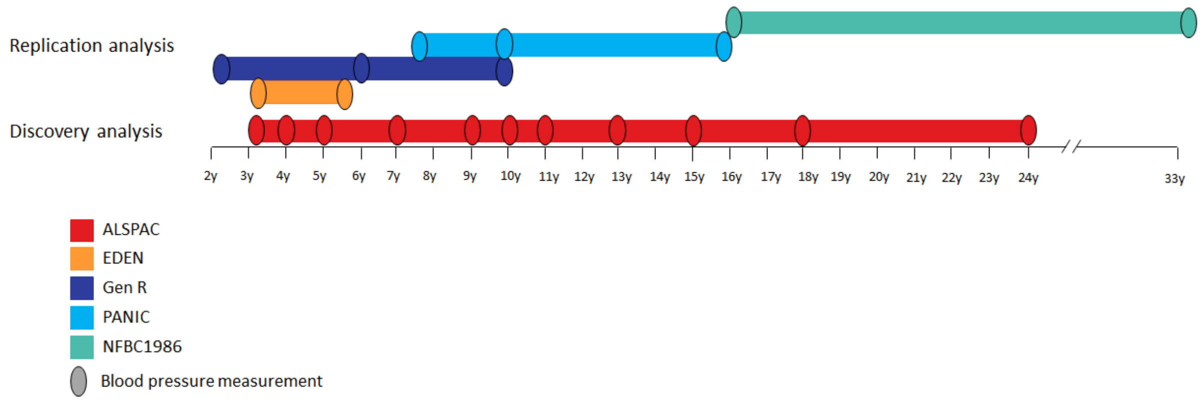

**Supplementary Figure 2.** Average age at the time of blood pressure measurements available in each study

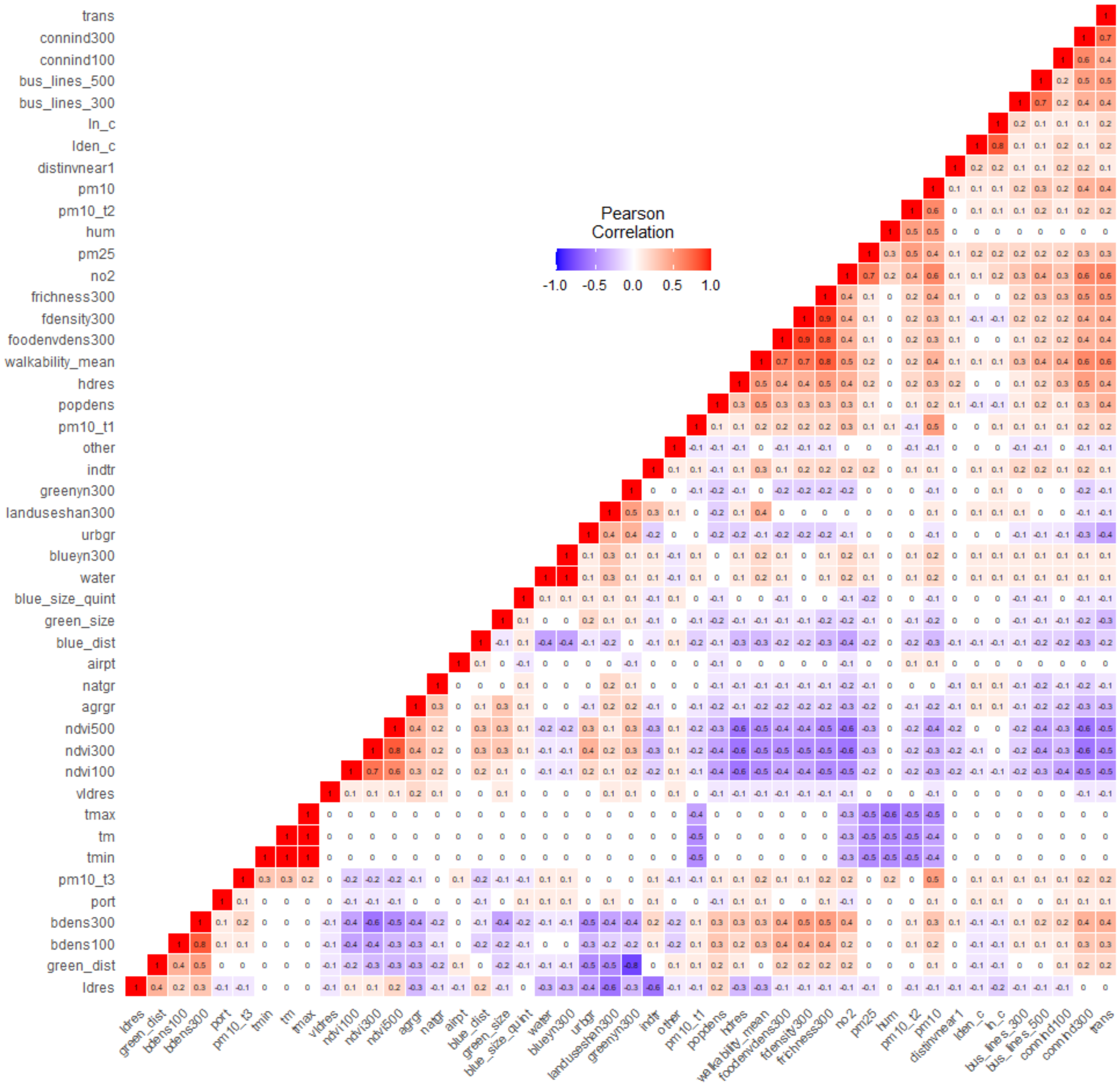

**Supplementary Figure 3.** Correlation matrix of the urban environmental exposures in ALSPAC.

Four exposures were excluded from the analyses due to very high correlation: minimum and maximum temperature (r=0.99 with mean temperature), % water land use within 300m buffer (r=0.99 with existence of blue area within 300m buffer), and facility richness within 300m buffer (r=0.91 with facility density within 300m buffer).

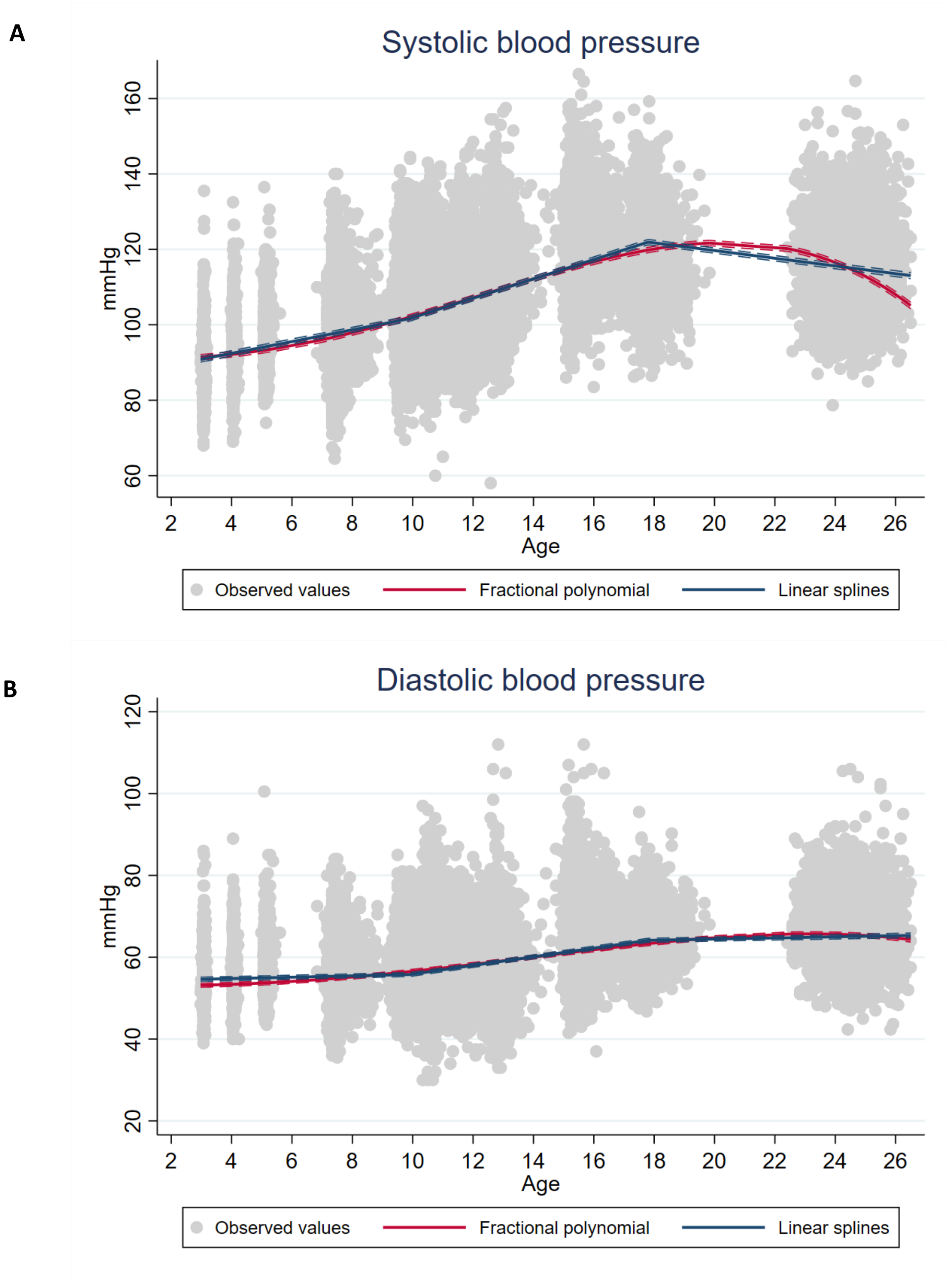

**Supplementary Figure 4.** Observed and average predicted (A) systolic blood pressure and (B) diastolic blood pressure trajectories in the ALSPAC.

Grey points are the observed blood pressure values, the red line is the average predicted trajectory using fractional polynomials and the blue line is the average predicted trajectory using linear splines. All covariates used in the adjustment were set to the mean value or the reference category: maternal education (high), age at delivery (28.9 years), ethnicity (White), area deprivation (least deprived), and sex (male).

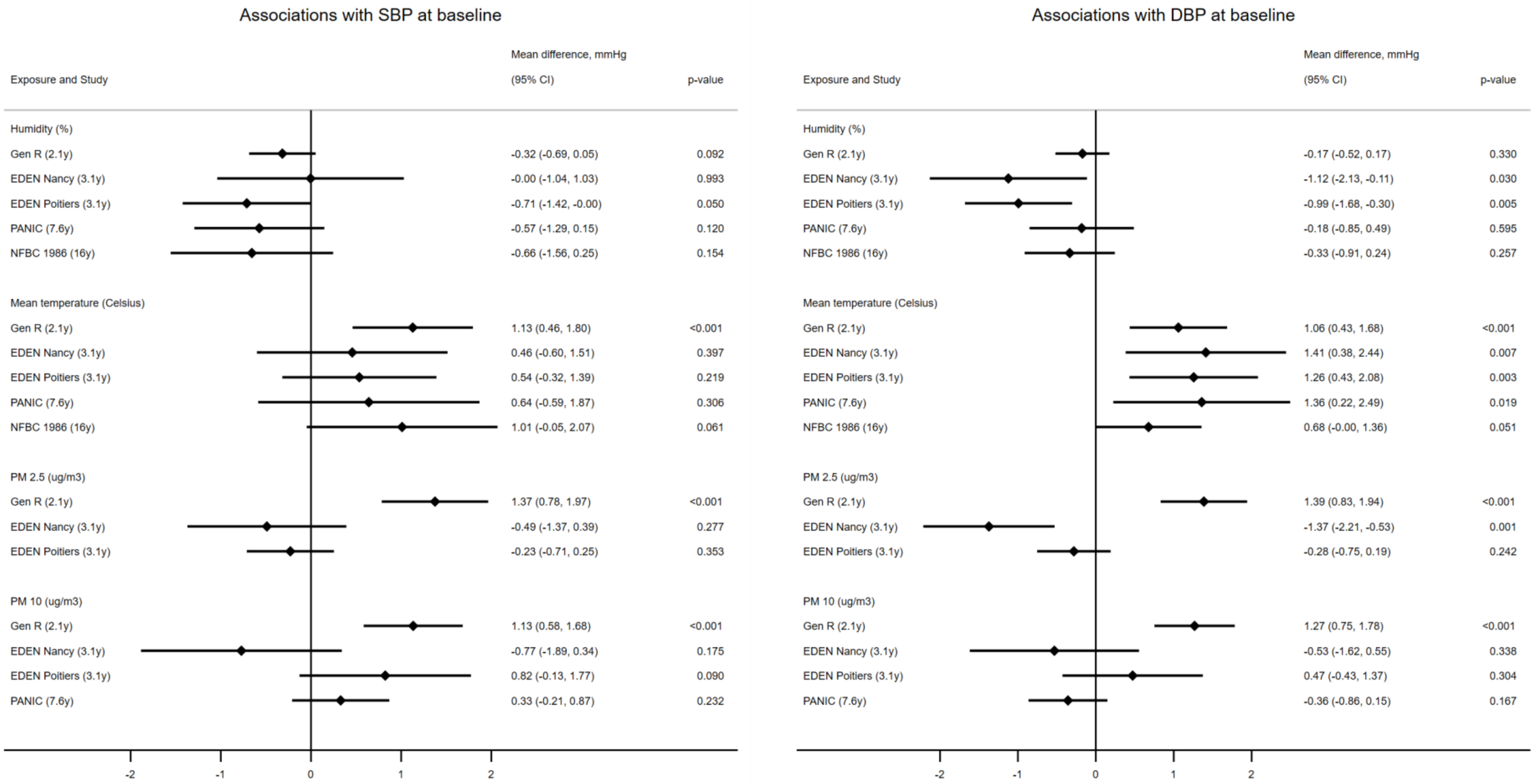

**Supplementary Figure 5.** Associations of humidity, mean temperature, $PM_{2.5}$ and $PM_{10}$ with SBP and DBP at baseline in GenR, EDEN, PANIC and NFBC1986.
DBP: diastolic blood pressure; $PM_{2.5}$: particulate matter <2.5 µm; $PM_{10}$: particulate matter <10 µm; SBP: systolic blood pressure

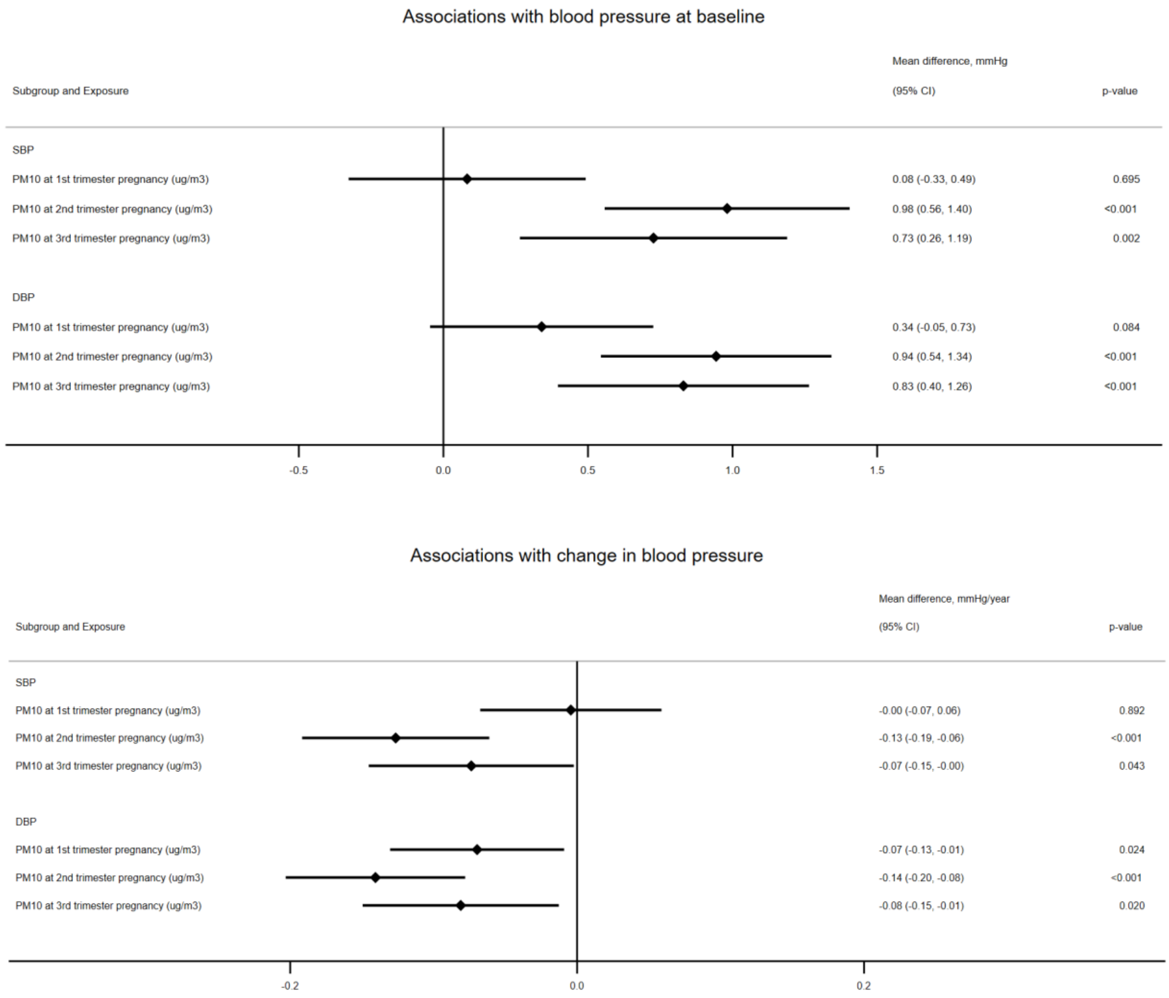

**Supplementary Figure 6.** Associations of $PM_{10}$ in different trimesters of pregnancy with blood pressure in GenR.

DBP: diastolic blood pressure; $PM_{2.5}$: particulate matter <2.5 µm; $PM_{10}$: particulate matter <10 µm; SBP: systolic blood pressure
